# Comparison of Population Characteristics in Real-World Clinical Oncology Databases in the US: Flatiron Health-Foundation Medicine Clinico-Genomic Databases, Flatiron Health Research Databases, and the National Cancer Institute SEER Population-Based Cancer Registry

**DOI:** 10.1101/2023.01.03.22283682

**Authors:** Tamara Snow, Jeremy Snider, Leah Comment, Stella Stergiopoulos, Virginia Fisher, Margaret McCusker, Cheryl Cho-Phan

**Affiliations:** Flatiron Health, Inc., New York, NY; Foundation Medicine Inc, Cambridge, MA

**Keywords:** cancer, real-world data, cancer registries, clinico-genomic database

## Abstract

**Background:** The Flatiron Health-Foundation Medicine Clinico-Genomic Databases (CGDBs) are de-identified, real-world data sources that link comprehensive genomic profiling (CGP) data with clinical data derived from electronic health records (EHRs) for patients with cancer. Comparing the CGDBs to the US population of patients with cancer allows researchers to understand the representativeness of a cohort when designing, conducting, and interpreting their analyses. The objective of this study was to compare the demographic and clinical characteristics of patients in the CGDBs with the Flatiron Health Research Databases (FHRDs) and The National Cancer Institute’s Surveillance, Epidemiology, and End Results (SEER) population-based cancer registry.

**Methods:** We compared disease-specific CGDBs that had corresponding disease-specific FHRDs with relevant SEER patients using demographic and clinical characteristics of patients with cancer who had documented care from January 1, 2011 to March 31, 2021. For CGDBs where a corresponding disease-specific FHRD does not exist, comparisons were only done against SEER. The SEER Incidence Data 1975-2018 Research Database was used for this analysis, of which patients with a relevant cancer diagnosis from January 1, 2011 to December 31, 2018 were included. Subgroup analyses were performed to address potential biases related to temporal drifts and allow for a more direct comparison of the datasets as well as to examine biases that may be due to data missingness. The impact of the determination to reimburse for next generation sequencing (NGS) testing was not feasible to analyze given the most recent SEER data was available only through the end of 2018 at the time this study was conducted.

**Results:** The overall distribution of cancer types was similar between the 22 CGDB databases and SEER. The overall distributions of gender and diagnosis year were similar across all databases. The CGDB has a lower proportion of patients who were aged 80 years or older at initial diagnosis compared to FHRD and SEER cohorts. However, narrower differences were observed in diseases where targeted therapies are approved and comprehensive genomic profiling is indicated (e.g., Melanoma, NSCLC). The proportion of incomplete records for race in the CGDB and FHRD was greater than in SEER. Completeness of stage varied by disease across all 3 cohorts, but was generally lower in CGDB and FHRD for clinical and data model design reasons. Overall the stage distributions for solid tumor cohorts were similar across CGDB and FHRD with SEER tending to have more earlier stage patients, which is expected given differences in data collection methods for the sources.

**Conclusion:** This comparative analysis of real-world, US-based oncology databases provides crucial insights into the similarities and differences in patient characteristics across these three types of data sources. Observed variances could be due to several factors, including differences in CGP testing dynamics and data collection approaches used to create each of the databases. Ongoing monitoring and evaluation of the representativeness of these databases will be critical to help researchers and regulators contextualize evidence from the CGDBs, particularly as the CGDBs are expected to change over time due to increased adoption of CGP as part of routine clinical practice for a growing number of cancers.

## Introduction

In recent years, high-quality, patient-level, real-world clinico-genomic data has demonstrated the potential to accelerate research and support clinical practice. These data have been used for a variety of applications, from informing therapeutic target identification and optimizing study design^(1,2)^ to strengthening regulatory submissions and delivering real-time insights that influence clinical practice.^(3)^ Real-world clinico-genomic data can also be used to train and validate machine learning algorithms, identifying new complex molecular signatures that may inform clinical decision making.^(4–6)^

Whether the data are reflective of the target population defined in a specific application is termed the representativeness of a dataset (i.e., the closeness with which sampled patients from a setting of interest align with the patient population at large in terms of relevant demographic and clinical characteristics).^(7)^ In cases of imperfect representativeness, individual analytic conclusions can only be appraised if the sources and degree of non-representativeness are understood, documented, and clearly communicated. Algorithms trained on datasets known to be highly representative of broad target populations are more likely to be robust to clinic-to-clinic variability in patient characteristics, whereas algorithms trained in datasets of unknown representativeness (e.g., a single academic center) require substantially more external validation. Furthermore, the potential for a dataset to generate a specific clinically-relevant insight to a target population depends on whether the relationship between the dataset’s source population and the target population is generalizable (or “causally transportable”).^(8)^

The Flatiron Health-Foundation Medicine Clinico-Genomic Databases (CGDBs) are de-identified, real-world data sources that link Foundation Medicine’s (FMI) comprehensive genomic profiling (CGP) data with Flatiron Health’s Research Network, a de-identified clinical database derived from electronic health records (EHRs) for patients with cancer. There are 22 individual CGDBs, 21 of which are disease-specific and one which is disease-agnostic. The representativeness of the CGDBs impacts the generalizability of the results researchers draw from analyses based on these data. Since the CGDBs are composed of a subset of the broader population of patients with cancer (i.e., only FMI CGP-tested patients from the Flatiron Health Research Network), it is important for researchers who use the CGDBs to understand how representative they are compared to the US population of patients with cancer when designing and conducting their analyses. We expect the representativeness of the CGDBs to this target population to differ depending on the extent to which next-generation sequencing (NGS) has become part of the standard of care in a given patient population.

The objective of this analysis was to compare the demographic and clinical characteristics of patients included in the CGDBs with patients in the disease-specific Flatiron Health Research Databases (FHRDs) and The National Cancer Institute’s Surveillance, Epidemiology, and End Results (SEER) population-based cancer registry. A related analysis previously compared the FHRDs with SEER and the Centers for Disease Control and Prevention’s National Program of Cancer Registries (NPCR) data.^(9)^ It showed that these databases have similar demographic characteristics and geographic distribution; however, there were overarching differences across the populations they respectively cover. Differences in data collection methods, such as sampling approaches, cadence of information ingestion, and rules of data acquisition, may explain some of the observed divergences. For this analysis, we used a similar methodological approach, with the addition of disease-agnostic comparisons and mapping of relevant American Joint Committee on Cancer (AJCC) tumor classifications^(10)^, to improve utility for researchers.

## Methods

This study is a retrospective observational data analysis comparing information on patients from three types of data sources: the FHRDs, the CGDBs, and the SEER cancer registry. Refer to **Table 1** for a summary of features of these databases. For patients in NPCR, case listing is not publicly available, therefore the current analysis was limited to SEER as a registry data source. The FHRDs and CGDBs are retrospective longitudinal databases, comprised of de-identified patient-level structured and unstructured data derived from EHRs within the Flatiron Health Research Network which at the time of study data cut-off (March 31, 2021), was composed of approximately 280 US cancer clinics (∼800 sites of care). Unlike the FHRDs, the CGDBs draw patients from the Flatiron Health Research Network who are linked by deterministic matching to de-identified genomic data derived from FMI CGP tests.^(11)^

**Table 1.** Comparison of General Features of SEER, FHRDs, and CGDBs as Types of Data Sources.

| Feature | SEER | FHRD<sup>29</sup> | CGDBs |
| --- | --- | --- | --- |
| **Source**<sup>20, 30</sup> | The SEER Program of the NCI is an authoritative source of information on cancer incidence and survival in the United States. SEER currently collects and publishes cancer incidence and survival data from population-based cancer registries. SEER 18 consists of CORE registries (Greater California; Greater Bay, comprised of San Francisco-Oakland and San Jose-Monterey; Los Angeles; Connecticut; Georgia; Hawaii; Illinois; Iowa; Kentucky; Louisiana; New Jersey; New Mexico, including a subcontract with Alaska Natives; Seattle-Puget Sound; Texas; and Utah) and ten Research Support registries (Arkansas, California Department of Public Health, Colorado, Michigan, Detroit, Missouri, New Hampshire, Oregon, Tennessee, and Wisconsin) representing approximately 27.8% of the U.S. population (based on 2010 census). | Clinical data are from EHRs used in community and academic oncology sites of care participating in the Flatiron Health Research Databases; clinical data consist of routinely collected information (i.e., without intentional or pre-specified collection). Information from non-oncology care settings or sites outside of participating clinics can be captured via scanned in documentation. | Clinical data source is the same as the FHRDs; genomic data are sourced from Foundation Medicine's CGP database.<br>Genomic data are sourced from hybrid capture-based NGS assays performed on patient samples in a CLIA-certified, CAP-accredited laboratory (Foundation Medicine, Cambridge, MA).<br>Tissue-based testing and the FoundationOne Liquid CDx assess >300 cancer-related genes, microsatellite instability, and tumor mutational burden. Liquid biopsy CGP performed prior to the FoundationOne Liquid CDx FDA approval were done with the 62-gene FoundationACT or the 70-gene FoundationOne Liquid assay. Both tissue- and liquid-biopsy based tests assess base substitutions, short insertions/deletions, rearrangements/fusions, and copy number variations. |
| **Collection approach** | Data collection is based on SEER and NAACCR data standards, and is intended to meet pre-specified registry program standards for completeness and quality.<br>All previous and current in situ/malignant reportable primaries<sup>1</sup> which occur(red) over the lifetime of the patient, regardless of where he/she lived at diagnosis are captured. | Originating information is collected as routinely documented or entered by participating practitioners.<br>For patients with multiple qualifying primaries within the same disease-specific build, the primary with the worse prognosis (i.e., higher stage) is captured.<br>For patients with multiple qualifying primaries in different disease-specific builds, the primary with the worst prognosis is captured for each disease. | Clinical data collection is the same as the FHRDs.<br>Genomic data are collected based on routine physician ordering of an FMI test to inform patient care.<br>For patients with multiple qualifying primaries within the disease-agnostic build, all primaries are captured. |
| **Curation** | Data are abstracted by tumor registrars, who enter text manually into data collection software. | Structured data are harmonized and mapped to common units and terminology and combined with unstructured data processed by technology-enabled manual abstraction.<br>Abstractors (clinical oncology nurses and tumor registrars) are trained to identify and extract relevant information by following optimized policies and procedures, and oversight is provided by oncology-trained clinicians. | Clinical data are curated in the same manner as the FHRDs.<br>Genomic data are curated according to a bioinformatics quality control protocol for variant calling and clinical reporting. CGP results that fail quality control are excluded from the database.<br>Patient's demographic information as available at Flatiron Health and FMI is uniquely and deterministically matched by a third-party linking vendor to create the linked CGDBs. |
| **Recency** | Releases data with a 2-year delay | Data are available with a 1-month delay | Data are available with a 3-month delay |
| **Key data model elements** |  |  |  |
| **Demographics** | Age, sex, race, ethnicity | Age, sex, race, ethnicity, geographic location | Age, sex, race<br>Ethnicity and geographic location are |

<sup>1</sup> A 'reportable' primary refers to the site/histology/behavior of the tumor and the years when reporting was required.

|  |  |  |  |
| --- | --- | --- | --- |
|  |  |  | not available for de-identification reasons |
| **Tumor details** | Date of initial diagnosis, primary tumor site, limited biomarker information, a morphology/histology, behavior, laterality and stage at diagnosis | Date of initial diagnosis, primary tumor site, morphology/histology, stage at diagnosis, standard biomarker information<sup>a</sup>, date of advanced/metastatic diagnosis as relevant for the patient's cancer type, sites of metastatic disease<sup>b</sup>. | Same as the FHRDs. |
| Abbreviations: CAP, College of American Pathologists; CGDBs, Flatiron Health-Foundation Medicine Clinico-Genomic Databases; CGP, comprehensive genomic profiling; CLIA, Clinical Laboratory Improvement Amendments; EHR, electronic health record; FHRDs, Flatiron Health Research Databases; NCI, National Cancer Institute; FMI, Foundation Medicine Inc; NAACCR, North American Association of Central Cancer Registries; NGS, next-generation sequencing; SEER, The Surveillance, Epidemiology, and End Results research database.<br><sup>a</sup>The biomarker portfolio offered by each database is different, with SEER providing information about a limited set of specific biomarkers that might predict outcomes or response to specific therapies and FHRDs and CGDBs offering a more extensive portfolio of biomarkers for which testing is considered standard of care.<br><sup>b</sup>Flatiron Health databases provide information on specific sites of metastatic disease for a selected set of tumor types. |  |  |  |

For the disease-specific FHRDs and CGDBs, there is a core clinical data model for each disease, which consists of a common set of clinically-relevant data variables derived from the EHR (e.g., initial diagnosis date, stage at initial diagnosis, advanced/metastatic diagnosis date, labs, medications ordered/received, date of death). These data are curated from either structured data or technology-enabled abstraction of unstructured data as relevant for the patient’s cancer type. These disease-specific core data models differ across diseases based upon the clinical relevance of capturing advanced and/or metastatic disease status as key events (e.g., patients with NSCLC are typically considered non-operable with locally-advanced/metastatic disease; whereas non-operable colorectal cancer is more closely associated with presence of distant/metastatic disease, and advanced/metastatic status are not relevant for leukemias since they are considered disseminated at diagnosis); the CGDB disease-agnostic database provides date of initial diagnosis and date of metastatic disease (when applicable) as key events for patients with solid tumors (see further details in **Supplementary Table I: Comparison of Key Cohort Selection Criteria for Disease-specific Builds**).

Additional disease-specific abstracted, derived, and composite variables are added to the core data model for each disease to form more comprehensive disease-specific databases that represent the FHRDs and the clinical data within the CGDBs. For both the FHRDs and the CGDBs, the data used in this study are approved for research use via Institutional review board (IRB) approval of a master study protocol with waiver of informed consent obtained prior to study conduct. The de-identified SEER data are available by request for research use via a data use agreement.^(12)^

### Flatiron Health Research Databases

Of the approximately 2.7 million patients with cancer in the Flatiron Health Research Network as of March 31, 2021, approximately 283,000 were probabilistically sampled into 20 disease-specific FHRDs, each of which corresponds to a unique chart-confirmed tumor type. The majority of solid tumor disease-specific FHRDs are further focused upon requiring patients to have advanced or metastatic disease (either at initial diagnosis or upon recurrence/progression) with the exception of the hepatocellular carcinoma, ovarian cancer, and small cell lung cancer FHRDs which do not have a requirement for advanced/metastatic disease, and the early-stage breast cancer FHRD. The 20 disease-specific FHRDs are: advanced endometrial cancer, acute myeloid leukemia (AML), advanced gastric/esophageal/gastroesophageal junction (GEJ) cancer, advanced melanoma, advanced head and neck squamous cell carcinoma, advanced non-small cell lung cancer (NSCLC), advanced urothelial carcinoma, chronic lymphocytic leukemia (CLL), diffuse large B-cell lymphoma (DLBCL), breast cancer (2 datasets: one for early-stage and another for metastatic breast cancer patients), follicular lymphoma (FL), hepatocellular carcinoma, mantle cell lymphoma, metastatic colorectal cancer, metastatic prostate cancer, metastatic renal cell carcinoma (RCC), metastatic pancreatic adenocarcinoma, multiple myeloma, ovarian cancer, and small cell lung cancer (SCLC).

All patients who met disease-specific inclusion criteria for each FHRD were eligible for inclusion in this study, as previously described.^(9)^ Patients were excluded from this analysis if they had a missing initial diagnosis date and/or birth year. This analysis also included patients whose data appear within multiple disease-specific databases since all analyses were done at the disease level.

### Flatiron Health-Foundation Medicine CGDBs

Of the approximately 2.7 million patients with cancer in the Flatiron Health Research Network as of March 31, 2021, approximately 69,000 received at least one FMI CGP test and thus were included in at least one CGDB. There is a subset of patients who meet criteria for inclusion into both a disease-specific CGDB and the respective disease-specific FHRD. These patients are probabilistically sampled into the FHRD, and therefore are included within both the CGDB and FHRD for that disease (**Figure 1**). All FMI genomic data tables are the same across all databases. Patients may qualify for inclusion in one of two types of CGDBs — within at least one of 21 disease-specific CGDBs or within the disease-agnostic CGDB:

1. Disease-specific CGDBs: Structured and unstructured clinical data derived from the EHR for patients with abstractor confirmation of a specific cancer type, linked with CGP data from FMI for patients whose sequenced specimen is consistent with the Flatiron Health abstracted cancer type. For solid tumor CGDBs, patients of all stages are eligible for inclusion with the exception of the bladder cancer, endometrial cancer, and prostate cancer CGDBs which require patients to have advanced or metastatic disease (either at initial diagnosis or upon recurrence/progression). There are 21 distinct disease-specific CGDBs: AML, breast cancer, CLL, colorectal cancer, DLBCL, advanced endometrial cancer, FL, gastric/esophageal/GEJ cancer, hepatocellular carcinoma, head and neck cancer squamous cell carcinoma, malignant pleural mesothelioma, mantle cell lymphoma, melanoma, multiple myeloma, NSCLC, ovarian cancer, pancreatic adenocarcinoma, metastatic prostate cancer, RCC, SCLC, and advanced bladder cancer. Patients can be included in more than one disease-specific database if they meet the cohort criteria for more than one cancer type.
2. Disease-agnostic CGDB: Structured and unstructured clinical data from the EHR including abstractor confirmation of each specific cancer type, for patients who do not meet the inclusion criteria for one of the 21 disease-specific CGDBs, linked with CGP data from FMI. This database is exclusively built to serve as a universal or core clinical data model for which key clinical variables applicable across diseases, including rare tumors, would provide key information to serve as a basis to conduct outcomes analysis. Patients who are included in one or more disease-specific CGDBs are not included in the disease-agnostic CGDB.

**Figure 1.**
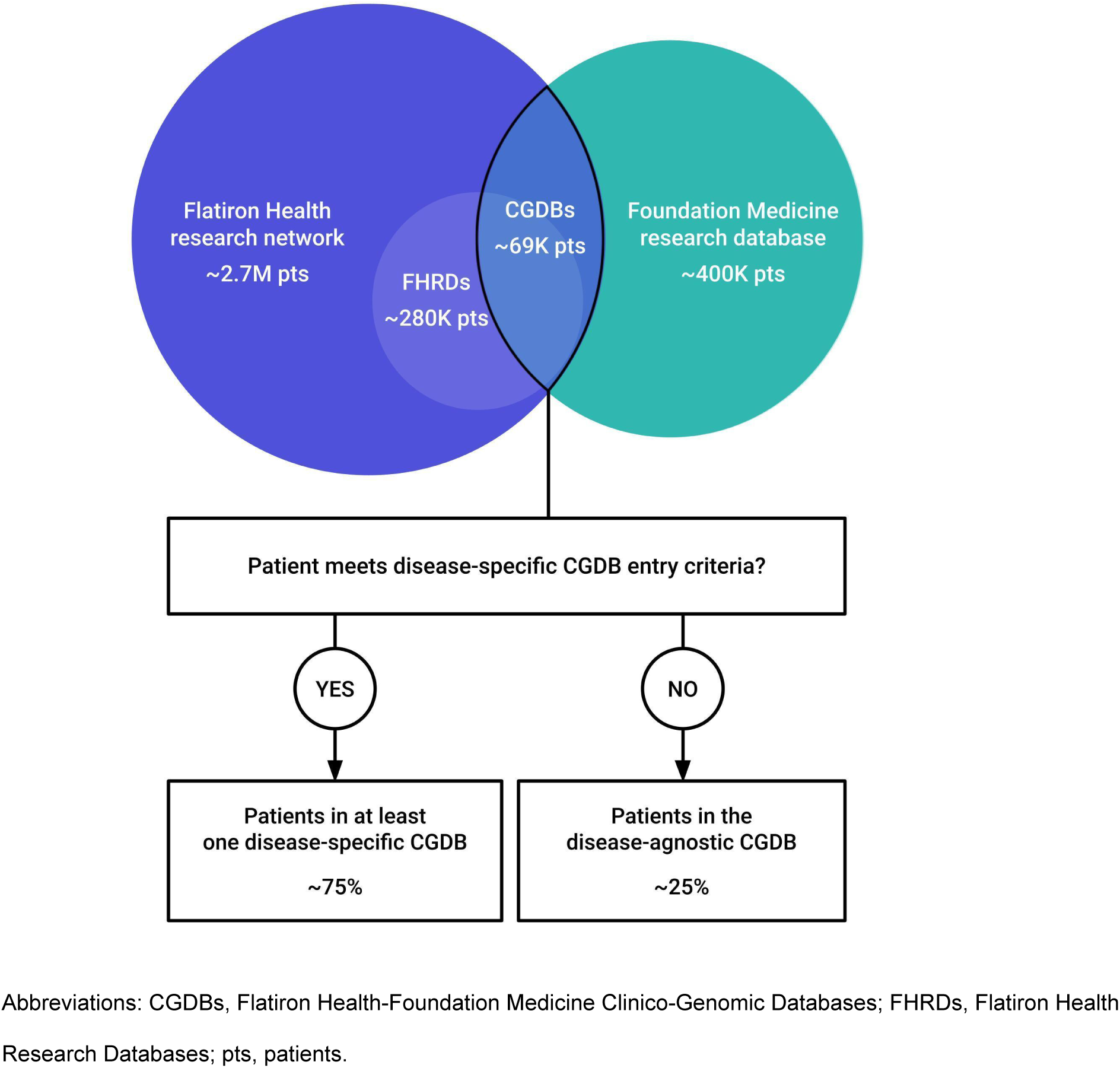
Relationship Between the FHRDs and CGDBs, and How CGDB Patients are Included in Either Disease-specific or Disease-agnostic CGDBs. Flatiron Health Research Network: A de-identified, real-world data source comprised of patient-level structured and unstructured data derived from electronic health records (EHRs) for patients with cancer which at the time of study data cut-off (March 31, 2021), was composed of approximately 280 US cancer clinics (∼800 sites of care). Foundation Medicine Research Database: A de-identified, real-world data source comprised of patient-level Foundation Medicine comprehensive genomic profiling data for patients with cancer. CGDBs: Flatiron Health-Foundation Medicine Clinico-Genomic Databases are retrospective longitudinal databases of de-identified, real-world clinical data sourced from the Flatiron Health Research Network that is uniquely and deterministically matched to Foundation Medicine’s comprehensive genomic profiling data; the CGDBs are comprised of 22 individual databases, 21 of which are disease-specific CGDBs and 1 which is disease-agnostic. FHRDs: Flatiron Health Research Databases are 20 disease-specific retrospective longitudinal databases of de-identified, EHR-derived real-world data sourced from the Flatiron Health Research Network via probabilistically sampling.

Data included in this study were collected through March 31, 2021. The following exclusion criteria were applied to the CGDBs to align with how the FHRDs are generated: patients were excluded if they had a missing diagnosis date and/or birth year, or if they had an initial diagnosis date prior to January 1, 2011.

### National Cancer Institute’s SEER Program

The National Cancer Institute’s SEER program collects and publishes cancer incidence and survival data from population-based cancer registries that covered approximately 34.6% of the US population at the time this study was conducted.^(13)^ SEER cancer registries routinely collect data on patient demographics, primary tumor site, tumor morphology, extent of disease, first course of treatment, and active follow-up for vital status; however, SEER does not capture disease recurrence, progression, or the development of distant metastasis in patients initially diagnosed with earlier stage disease. The SEER Incidence Data 1975-2018 Research Database was used for this analysis,^(13)^ of which patients with a relevant cancer diagnosis from January 1, 2011 to December 31, 2018 were included, which represents 27.8% of the US population.

### Data Source Considerations

Differences in the inclusion criteria between the disease-specific FHRDs, CGDBs, and SEER are summarized as follows: patients are included in CGDBs regardless of extent of disease with the exception of the advanced bladder cancer, advanced endometrial cancer, and metastatic prostate cancer CGDBs, which follow the same staging criteria as the respective FHRDs. Additional FHRDs include only patients with advanced or metastatic disease (i.e., colorectal cancer, gastric/esophageal/GEJ cancer, head and neck squamous cell carcinoma, melanoma, NSCLC, pancreatic adenocarcinoma, and RCC) (see **Supplementary Table 1)**. All SEER cohorts include patients regardless of extent of disease. The hepatocellular carcinoma, ovarian cancer, and small cell lung cancer FHRDs include patients regardless of extent of disease aligned with the respective CGDB and SEER cohorts. Second, the urothelial carcinoma FHRD includes only patients with urothelial carcinoma tumor histology originating in the urothelial tract or bladder, while the bladder cancer CGDB and SEER bladder cancer cohort include tumors with any carcinoma histology from the same sites of origin. Third, the following FHRDs have different diagnosis date windows which are not applied in the corresponding CGDB and SEER cohorts: AML (≥January 1, 2014), metastatic colorectal cancer (≥January 1, 2013), advanced endometrial cancer (≥January 1, 2013), metastatic pancreatic adenocarcinoma (≥January 1, 2014), metastatic prostate cancer (≥January 1, 2013), and SCLC (≥January 1, 2013) (see **Supplementary Table 1)**. Finally, at the time of data cut-off, all CGDBs (disease-specific and disease-agnostic) included patients from more sites of care within the Flatiron Health Research Network than the disease-specific FHRDs due to sampling rules applied in the FHRDs.

### Comparative Analysis

#### Mapping Variables Across Databases

For each cancer type, demographic and clinical characteristics including race, age, year of diagnosis, and stage at diagnosis were compared between the CGDBs, FHRDs, and SEER databases. Refer to **Supplementary Table 2** for variable definitions. In the FHRDs and CGDBs, cancer staging information was collected as entered into the EHR by the treating physician or as otherwise assessed by Flatiron Health abstractors based on standard rules. The applicable staging criteria for solid tumors were those of the American Joint Committee on Cancer (AJCC) 7th edition and 8th edition manuals,^(10,14)^ including the adoption of the International Federation of Gynecology and Obstetrics (FIGO) staging system by the AJCC for endometrial cancer.^(15)^ For hematologic malignancies, where TNM-type staging systems are not of practical value, alternative classification systems were collected: World Health Organization (WHO) Classification for AML^(16)^, Rai staging for CLL^(17)^, Ann Arbor Staging System for DLBCL, FL, and mantle cell lymphoma^(18)^, and the International Staging System (ISS) for multiple myeloma^(19)^ as entered into the EHR by the treating physician or otherwise as assessed by Flatiron Health abstractors based on standard rules.

For SEER, diagnosis and staging information were abstracted from various sources including medical records and pathology reports, with staging information derived according to the relevant approaches defined for each time period.^(20)^ For patients diagnosed before 2016, SEER’s derived AJCC Stage Group v7 variable was used. For patients diagnosed on or after 2016, SEER’s derived combined stage group was used. The one exception was for the early breast cancer comparison where patients were included if one of the following variables contained a value of 1, 2, or 3 from: derived_ajcc_stage_group_6th_ed_2004_2015, derived_seer_cmb_stg_grp_2016_2017, derived_eod_2018_stage_group_2018. Since SEER has different classification systems for cancer type compared to that of the FHRDs and CGDBs, cancer type was mapped by the study authors between the databases using ICD-9 and ICD-10 codes, AJCC site classification and either SEER Site recode ICD-O-3/WHO 2008 variables,^(21)^ or ICD-O-3 site and histology codes.

#### Comparative Analyses

Of the 22 CGDBs, 20 disease-specific CGDBs have a corresponding FHRD disease-specific database, which were compared with relevant SEER patients. Because the breast cancer CGDB combines Flatiron Health’s early and metastatic breast cancer clinical data models and source populations, three tables were generated for this disease: 1) patients with metastatic breast cancer at any time in the breast cancer FHRD vs patients with metastatic breast cancer at any time in the breast cancer CGDB vs patients with metastatic breast cancer at the time of initial diagnosis in SEER (patients with breast cancer in SEER were included if SEER’s Localized/Regional/Distant (LRD) Summary stage (2004+) variable = “Distant”); 2) early breast cancer FHRD (patients with stage I-III explicitly documented or patients with documented TNM stage equivalent to stage I-III disease without stage explicitly documented) vs patients with stage I-III breast cancer explicitly documented only in the breast cancer CGDB vs patients with stage I-III breast cancer in SEER (patients with breast cancer in SEER were included if the following variables contained 1, 2, or 3: derived_ajcc_stage_group_6th_ed_2004_2015, derived_seer_cmb_stg_grp_2016_2017, derived_eod_2018_stage_group_2018); 3) the full breast cancer CGDB vs all patients with breast cancer in SEER.

For CGDBs where a corresponding disease-specific database from the FHRD does not exist, comparisons were only done against SEER. These include all CGDB patients (patients within any disease-specific CGDB or the disease-agnostic CGDB) vs all SEER patients, all CGDB patients with a hematologic malignancy vs all SEER patients with a hematologic malignancy (includes patients in hematologic disease-specific CGDBs and with hematologic tumor types included in the disease-agnostic CGDB), all CGDB patients with a malignant solid tumor (includes patients in solid tumor disease-specific CGDBs and with solid tumor types within the disease-agnostic CGDB) vs all SEER patients with a malignant solid tumor, the malignant pleural mesothelioma CGDB vs SEER patients with malignant pleural mesothelioma, and two highly represented disease types in the disease-agnostic CGDB (patients with cholangiocarcinoma in the CGDB vs patients with cholangiocarcinoma in SEER and patients with glioma in the CGDB vs patients with glioma in SEER).

For the purposes of this study, we limited analyses to patients with a diagnosis date ≥2011 across the CGDBs and SEER in order to compare relatively recent patient populations. In order to compare common cohorts across all three databases, descriptive analyses were performed not only for all patients available for analysis across the entire time frame of January 1, 2011 to March 31, 2021 in the FHRDs and CGDBs, but also in the subset of patients available for analysis from January 1, 2011 to December 31, 2018 to align with SEER data availability. To compare different data sources, differences in proportions for selected variables were examined, using a denominator of all patients in the given disease.

#### Subgroup Analyses

As was done in Ma *et al* 2020,^(9)^ we conducted a subgroup analysis to allow for a more direct comparison of the datasets and to understand potential biases related to temporal drifts (detailed below). To assess this dynamic, a subgroup analysis was performed on patients with stage IV disease at diagnosis in each disease where this designation is relevant. This subgroup was chosen since these patients are expected to have shorter survival times, which means their date of diagnosis would be expected to be closer to the database entry point. The potential biases to address were threefold:

1. The FHRDs and CGDBs include both incident and prevalent cancer cases, whereas SEER only collects incident cancer cases. Therefore, the FHRDs and CGDBs may include patients at the time of diagnosis as well as patients whose initial diagnosis dates were in the past. It is possible that these cases have long intervening periods between the initial diagnosis date and the date of entry into the FHRDs or CGDBs, which may introduce a potential bias for patient characteristics that are associated with longer survival times when compared with strictly incident cases in SEER.
2. Additionally, the patient characteristics included in this analysis may be associated with a cancer diagnosis during a specific time period, which can influence the distributions within a given year.
3. Lastly, use of FMI CGP tests has changed over time and to varying degrees across diseases; differences could be due to variability in utility of broad genomic panel sequencing to inform treatment for a given disease and relatedly the fact that Medicare’s National Coverage Determination to reimburse for NGS testing did not become effective until March 18, 2018^(22)^ and the most recent SEER data used in this analysis are available only through the end of 2018.

Finally, in order to examine biases that may be due to data missingness, we conducted a sensitivity analysis that included only patients with complete data across all study variables.

## Results

The FHRD and CGDB disease-specific databases vary in size depending on the incidence and prevalence of the disease. The FHRDs also vary in size depending on the probabilistic sampling approach used to create each disease-specific database and the length of time the database has been active. The CGDBs also vary in size depending on how commonly CGP is utilized in clinical practice for managing each specific disease; CGDBs are not subject to probabilistic sampling. The overall distribution of cancer types is similar between the 22 CGDBs and SEER, with small variances likely due to differences in CGP testing dynamics across diseases and in the source of respective data (**Table 2**). In the sensitivity analysis only including patients with complete data, no substantial differences in clinical or demographic characteristics were noted between those patients and the overall patient populations (data not published).

**Table 2.**
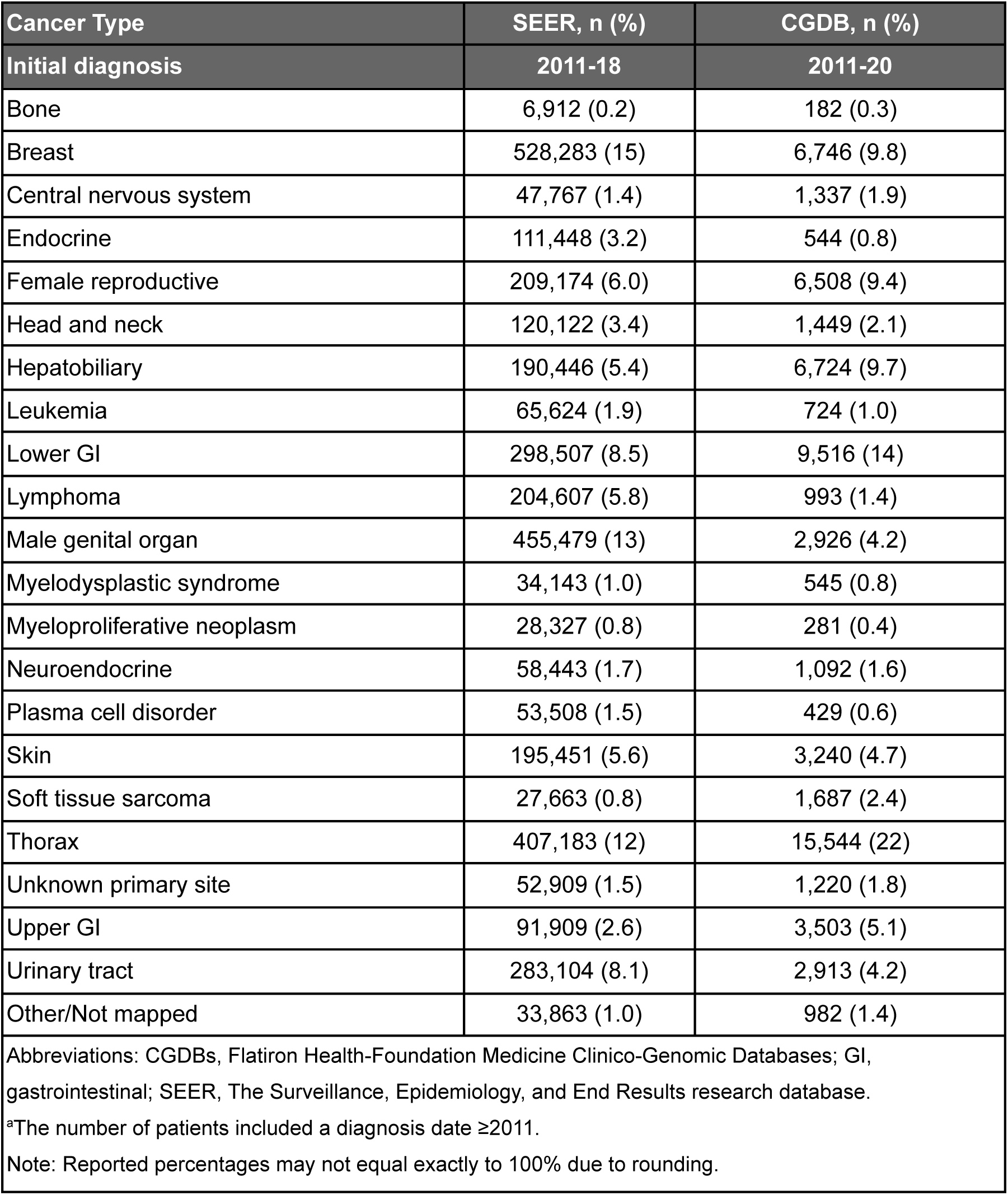
Distribution of Cancer Types across CGDBs (N=69,085)^a^ and SEER (N=3,504,872)

| Cancer Type | SEER, n (%) | CGDB, n (%) |
| --- | --- | --- |
| Initial diagnosis | 2011-18 | 2011-20 |
| Bone | 6,912 (0.2) | 182 (0.3) |
| Breast | 528,283 (15) | 6,746 (9.8) |
| Central nervous system | 47,767 (1.4) | 1,337 (1.9) |
| Endocrine | 111,448 (3.2) | 544 (0.8) |
| Female reproductive | 209,174 (6.0) | 6,508 (9.4) |
| Head and neck | 120,122 (3.4) | 1,449 (2.1) |
| Hepatobiliary | 190,446 (5.4) | 6,724 (9.7) |
| Leukemia | 65,624 (1.9) | 724 (1.0) |
| Lower GI | 298,507 (8.5) | 9,516 (14) |
| Lymphoma | 204,607 (5.8) | 993 (1.4) |
| Male genital organ | 455,479 (13) | 2,926 (4.2) |
| Myelodysplastic syndrome | 34,143 (1.0) | 545 (0.8) |
| Myeloproliferative neoplasm | 28,327 (0.8) | 281 (0.4) |
| Neuroendocrine | 58,443 (1.7) | 1,092 (1.6) |
| Plasma cell disorder | 53,508 (1.5) | 429 (0.6) |
| Skin | 195,451 (5.6) | 3,240 (4.7) |
| Soft tissue sarcoma | 27,663 (0.8) | 1,687 (2.4) |
| Thorax | 407,183 (12) | 15,544 (22) |
| Unknown primary site | 52,909 (1.5) | 1,220 (1.8) |
| Upper GI | 91,909 (2.6) | 3,503 (5.1) |
| Urinary tract | 283,104 (8.1) | 2,913 (4.2) |
| Other/Not mapped | 33,863 (1.0) | 982 (1.4) |
| Abbreviations: CGDBs, Flatiron Health-Foundation Medicine Clinico-Genomic Databases; GI, gastrointestinal; SEER, The Surveillance, Epidemiology, and End Results research database.<br><sup>a</sup>The number of patients included a diagnosis date ≥2011.<br>Note: Reported percentages may not equal exactly to 100% due to rounding. |  |  |

**Tables 3–23** present CGDB vs FHRD vs SEER comparisons for the following cancer types: AML (**Table 3**), advanced urothelial (or bladder) cancer (**Table 4**), early breast cancer (**Table 5**), metastatic breast cancer (**Table 6**), CLL (**Table 7**), colorectal cancer (**Table 8**), DLBCL (**Table 9**), advanced endometrial cancer (**Table 10**), FL (**Table 11**), gastric/esophageal/GEJ cancer (**Table 12**), squamous cell carcinoma of the head and neck (**Table 13**), hepatocellular carcinoma (**Table 14**), mantle cell lymphoma (**Table 15**), melanoma (**Table 16**), multiple myeloma (**Table 17**), NSCLC (**Table 18**), ovarian cancer (**Table 19**), pancreatic adenocarcinoma (**Table 20**), metastatic prostate cancer (**Table 21**), renal cell carcinoma (**Table 22**), and SCLC (**Table 23**).

**Table 3.** Acute Myeloid Leukemia – Comparison of Demographic and Clinical Characteristics Between CGDB vs FHRD vs SEER^a^.

| Characteristic | All eligible, n (%) |  |  |  |  |
| --- | --- | --- | --- | --- | --- |
|  | SEER | FHRD |  | CGDB |  |
| Initial diagnosis | 2011–18 | 2011–18 | 2011–20 | 2011–18 | 2011–20 |
|  | N=33,392 | N=4,510 | N=6,489 | N=223 | N=307 |
| **Age at initial diagnosis (years)** |  |  |  |  |  |
| 0–19 | 1,609 (4.8) | 47 (1.0) | 59 (0.9) | 5 (2.2) | 6 (2.0) |
| 20–34 | 1,919 (5.7) | 249 (5.5) | 311 (4.8) | 20 (9.0) | 21 (6.8) |
| 35–49 | 2,987 (8.9) | 433 (9.6) | 565 (8.7) | 15 (6.7) | 18 (5.9) |
| 50–64 | 7,419 (22) | 964 (21) | 1,360 (21) | 54 (24) | 62 (20) |
| 65–79 | 12,398 (37) | 1,930 (43) | 2,834 (44) | 102 (46) | 144 (47) |
| 80+ | 7,060 (21) | 887 (20) | 1,360 (21) | 27 (12) | 56 (18) |
| **Sex** |  |  |  |  |  |
| Female | 15,089 (45) | 1,932 (43) | 2,755 (42) | 98 (44) | 137 (45) |
| Male | 18,303 (55) | 2,578 (57) | 3,734 (58) | 125 (56) | 170 (55) |
| **Race** |  |  |  |  |  |
| Asian | 2,777 (8.3) | 111 (2.5) | 146 (2.2) | 7 (3.1) | 10 (3.3) |
| Black | 3,098 (9.3) | 300 (6.7) | 424 (6.5) | 10 (4.5) | 19 (6.2) |
| Other | 221 (0.7) | 508 (11) | 860 (13) | 27 (12) | 43 (14) |
| Unknown | 168 (0.5) | 411 (9.1) | 667 (10) | 8 (3.6) | 11 (3.6) |
| White | 27,128 (81) | 3,180 (71) | 4,392 (68) | 171 (77) | 224 (73) |
| **WHO classification at diagnosis** |  |  |  |  |  |
| AML and myelodysplastic syndromes - therapy related | 1,591 (4.8) | 315 (7.0) | 486 (7.5) | 20 (9.0) | 28 (9.1) |
| AML with myelodysplasia related changes | 3,424 (10) | 1,396 (31) | 2,020 (31) | 78 (35) | 108 (35) |
| AML with recurrent genetic abnormalities | 3,996 (12) | 732 (16) | 1,069 (16) | 24 (11) | 32 (10) |
| AML, not otherwise categorized | 24,285 (73) | 775 (17) | 1,091 (17) | 36 (16) | 53 (17) |
| Myeloid sarcoma | <5 (<0.1) | 25 (0.6) | 37 (0.6) | <5 (<2.2) | 5 (1.6) |
| Unknown | 96 (0.3) | 1,267 (28) | 1,786 (28) | 62 (28) | 81 (26) |
| **Year of initial diagnosis** |  |  |  |  |  |
| 2011 | 3,919 (12) | NA | NA | 6 (2.7) | 6 (2.0) |
| 2012 | 4,083 (12) | NA | NA | 10 (4.5) | 10 (3.3) |
| 2013 | 4,139 (12) | NA | NA | 16 (7.2) | 16 (5.2) |
| 2014 | 4,265 (13) | 820 (18) | 820 (13) | 32 (14) | 32 (10) |
| 2015 | 4,179 (13) | 926 (21) | 926 (14) | 30 (13) | 30 (9.8) |
| 2016 | 4,267 (13) | 889 (20) | 889 (14) | 43 (19) | 43 (14) |
| 2017 | 4,225 (13) | 931 (21) | 931 (14) | 43 (19) | 43 (14) |
| 2018 | 4,315 (13) | 944 (21) | 944 (15) | 43 (19) | 43 (14) |
| 2019 | NA | NA | 1,056 (16) | NA | 47 (15) |
| 2020 | NA | NA | 839 (13) | NA | 34 (11) |
| 2021 | NA | NA | 84 (1.3) | NA | <5 (<2.2) |

Abbreviations: AML, acute myeloid leukemia; CGDB, Flatiron Health-Foundation Medicine Clinico-Genomic Database; FHRD, Flatiron Health Research Database; NA, not applicable; SEER, The Surveillance, Epidemiology, and End Results research database; WHO, World Health Organization.
<sup>a</sup>Proportion of FHRD patients reported by Year of initial diagnosis is not directly comparable to the SEER and CGDB cohorts given that the initial diagnosis year for inclusion began in 2014.
Note: Reported percentages may not equal exactly to 100% due to rounding.

**Table 4.**
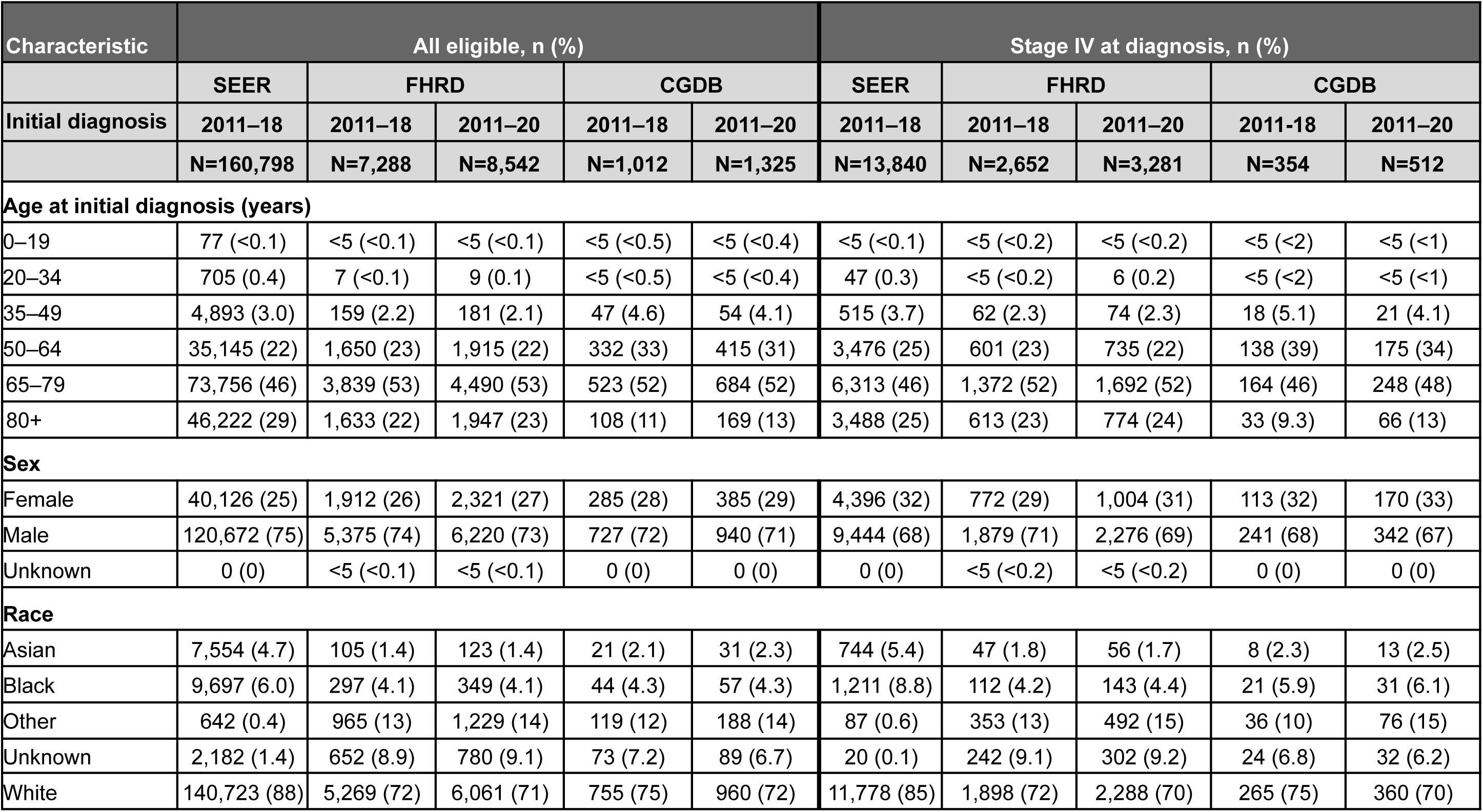
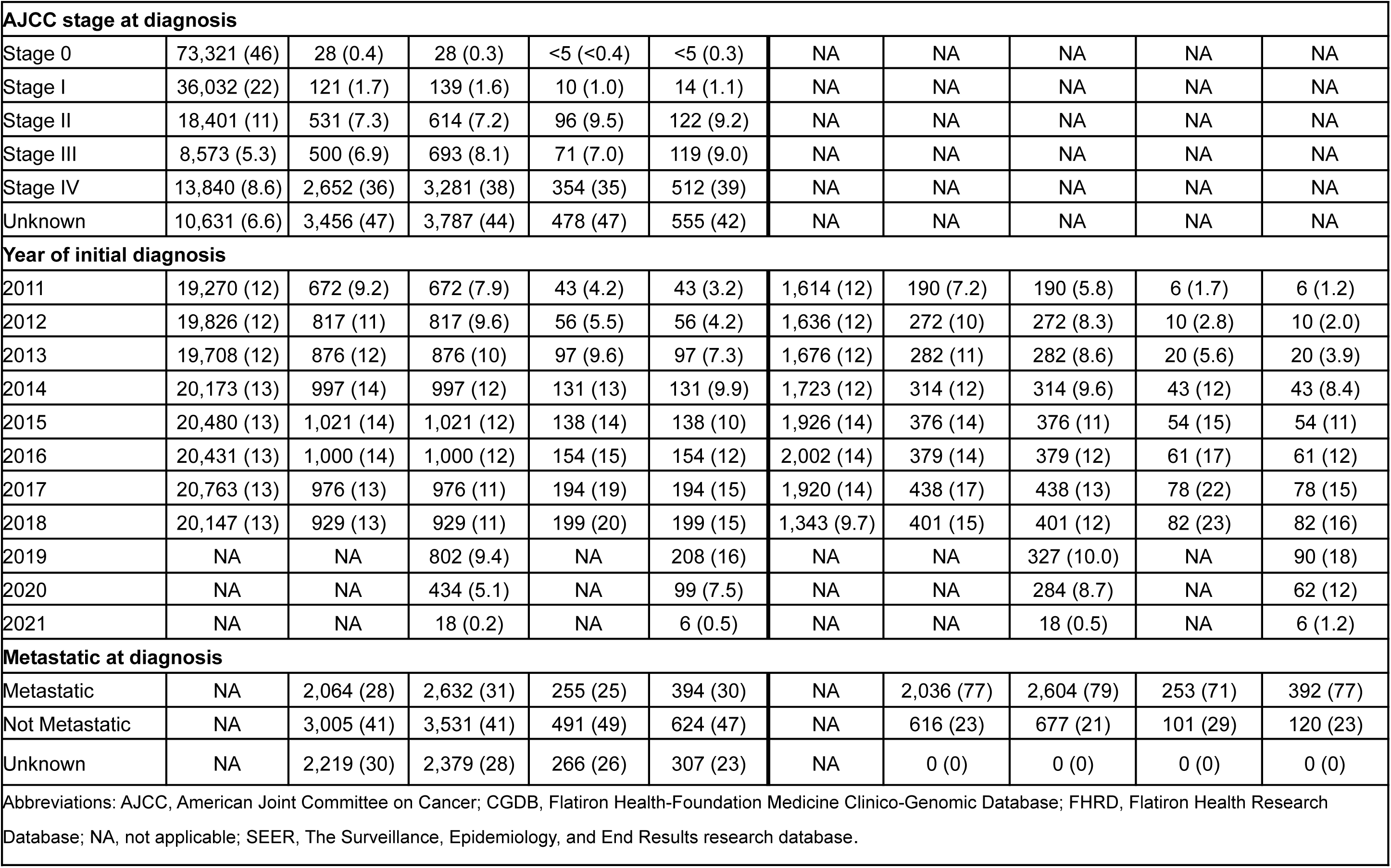
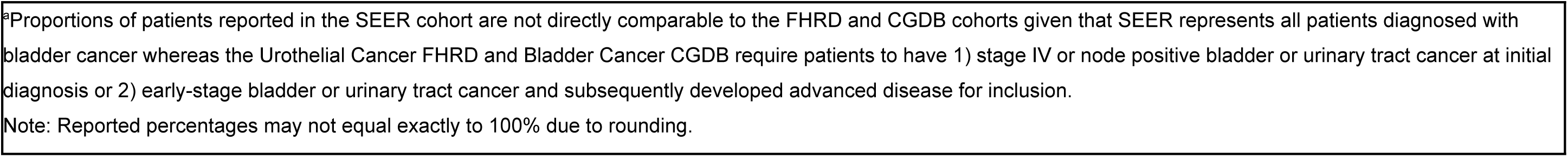
Bladder Cancer – Comparison of Demographic and Clinical Characteristics Between SEER (advanced and non-advanced at diagnosis) vs FHRD (advanced at any time) vs CGDB (advanced at any time)^a^.

| Characteristic | All eligible, n (%) |  |  |  |  | Stage IV at diagnosis, n (%) |  |  |  |  |
| --- | --- | --- | --- | --- | --- | --- | --- | --- | --- | --- |
|  | SEER | FHRD |  | CGDB |  | SEER | FHRD |  | CGDB |  |
| Initial diagnosis | 2011–18 | 2011–18 | 2011–20 | 2011–18 | 2011–20 | 2011–18 | 2011–18 | 2011–20 | 2011–18 | 2011–20 |
|  | N=160,798 | N=7,288 | N=8,542 | N=1,012 | N=1,325 | N=13,840 | N=2,652 | N=3,281 | N=354 | N=512 |
| **Age at initial diagnosis (years)** |  |  |  |  |  |  |  |  |  |  |
| 0–19 | 77 (<0.1) | <5 (<0.1) | <5 (<0.1) | <5 (<0.5) | <5 (<0.4) | <5 (<0.1) | <5 (<0.2) | <5 (<0.2) | <5 (<2) | <5 (<1) |
| 20–34 | 705 (0.4) | 7 (<0.1) | 9 (0.1) | <5 (<0.5) | <5 (<0.4) | 47 (0.3) | <5 (<0.2) | 6 (0.2) | <5 (<2) | <5 (<1) |
| 35–49 | 4,893 (3.0) | 159 (2.2) | 181 (2.1) | 47 (4.6) | 54 (4.1) | 515 (3.7) | 62 (2.3) | 74 (2.3) | 18 (5.1) | 21 (4.1) |
| 50–64 | 35,145 (22) | 1,650 (23) | 1,915 (22) | 332 (33) | 415 (31) | 3,476 (25) | 601 (23) | 735 (22) | 138 (39) | 175 (34) |
| 65–79 | 73,756 (46) | 3,839 (53) | 4,490 (53) | 523 (52) | 684 (52) | 6,313 (46) | 1,372 (52) | 1,692 (52) | 164 (46) | 248 (48) |
| 80+ | 46,222 (29) | 1,633 (22) | 1,947 (23) | 108 (11) | 169 (13) | 3,488 (25) | 613 (23) | 774 (24) | 33 (9.3) | 66 (13) |
| **Sex** |  |  |  |  |  |  |  |  |  |  |
| Female | 40,126 (25) | 1,912 (26) | 2,321 (27) | 285 (28) | 385 (29) | 4,396 (32) | 772 (29) | 1,004 (31) | 113 (32) | 170 (33) |
| Male | 120,672 (75) | 5,375 (74) | 6,220 (73) | 727 (72) | 940 (71) | 9,444 (68) | 1,879 (71) | 2,276 (69) | 241 (68) | 342 (67) |
| Unknown | 0 (0) | <5 (<0.1) | <5 (<0.1) | 0 (0) | 0 (0) | 0 (0) | <5 (<0.2) | <5 (<0.2) | 0 (0) | 0 (0) |
| **Race** |  |  |  |  |  |  |  |  |  |  |
| Asian | 7,554 (4.7) | 105 (1.4) | 123 (1.4) | 21 (2.1) | 31 (2.3) | 744 (5.4) | 47 (1.8) | 56 (1.7) | 8 (2.3) | 13 (2.5) |
| Black | 9,697 (6.0) | 297 (4.1) | 349 (4.1) | 44 (4.3) | 57 (4.3) | 1,211 (8.8) | 112 (4.2) | 143 (4.4) | 21 (5.9) | 31 (6.1) |
| Other | 642 (0.4) | 965 (13) | 1,229 (14) | 119 (12) | 188 (14) | 87 (0.6) | 353 (13) | 492 (15) | 36 (10) | 76 (15) |
| Unknown | 2,182 (1.4) | 652 (8.9) | 780 (9.1) | 73 (7.2) | 89 (6.7) | 20 (0.1) | 242 (9.1) | 302 (9.2) | 24 (6.8) | 32 (6.2) |
| White | 140,723 (88) | 5,269 (72) | 6,061 (71) | 755 (75) | 960 (72) | 11,778 (85) | 1,898 (72) | 2,288 (70) | 265 (75) | 360 (70) |

| AJCC stage at diagnosis |  |  |  |  |  |  |  |  |  |  |
| --- | --- | --- | --- | --- | --- | --- | --- | --- | --- | --- |
| Stage 0 | 73,321 (46) | 28 (0.4) | 28 (0.3) | <5 (<0.4) | <5 (0.3) | NA | NA | NA | NA | NA |
| Stage I | 36,032 (22) | 121 (1.7) | 139 (1.6) | 10 (1.0) | 14 (1.1) | NA | NA | NA | NA | NA |
| Stage II | 18,401 (11) | 531 (7.3) | 614 (7.2) | 96 (9.5) | 122 (9.2) | NA | NA | NA | NA | NA |
| Stage III | 8,573 (5.3) | 500 (6.9) | 693 (8.1) | 71 (7.0) | 119 (9.0) | NA | NA | NA | NA | NA |
| Stage IV | 13,840 (8.6) | 2,652 (36) | 3,281 (38) | 354 (35) | 512 (39) | NA | NA | NA | NA | NA |
| Unknown | 10,631 (6.6) | 3,456 (47) | 3,787 (44) | 478 (47) | 555 (42) | NA | NA | NA | NA | NA |
| Year of initial diagnosis |  |  |  |  |  |  |  |  |  |  |
| 2011 | 19,270 (12) | 672 (9.2) | 672 (7.9) | 43 (4.2) | 43 (3.2) | 1,614 (12) | 190 (7.2) | 190 (5.8) | 6 (1.7) | 6 (1.2) |
| 2012 | 19,826 (12) | 817 (11) | 817 (9.6) | 56 (5.5) | 56 (4.2) | 1,636 (12) | 272 (10) | 272 (8.3) | 10 (2.8) | 10 (2.0) |
| 2013 | 19,708 (12) | 876 (12) | 876 (10) | 97 (9.6) | 97 (7.3) | 1,676 (12) | 282 (11) | 282 (8.6) | 20 (5.6) | 20 (3.9) |
| 2014 | 20,173 (13) | 997 (14) | 997 (12) | 131 (13) | 131 (9.9) | 1,723 (12) | 314 (12) | 314 (9.6) | 43 (12) | 43 (8.4) |
| 2015 | 20,480 (13) | 1,021 (14) | 1,021 (12) | 138 (14) | 138 (10) | 1,926 (14) | 376 (14) | 376 (11) | 54 (15) | 54 (11) |
| 2016 | 20,431 (13) | 1,000 (14) | 1,000 (12) | 154 (15) | 154 (12) | 2,002 (14) | 379 (14) | 379 (12) | 61 (17) | 61 (12) |
| 2017 | 20,763 (13) | 976 (13) | 976 (11) | 194 (19) | 194 (15) | 1,920 (14) | 438 (17) | 438 (13) | 78 (22) | 78 (15) |
| 2018 | 20,147 (13) | 929 (13) | 929 (11) | 199 (20) | 199 (15) | 1,343 (9.7) | 401 (15) | 401 (12) | 82 (23) | 82 (16) |
| 2019 | NA | NA | 802 (9.4) | NA | 208 (16) | NA | NA | 327 (10.0) | NA | 90 (18) |
| 2020 | NA | NA | 434 (5.1) | NA | 99 (7.5) | NA | NA | 284 (8.7) | NA | 62 (12) |
| 2021 | NA | NA | 18 (0.2) | NA | 6 (0.5) | NA | NA | 18 (0.5) | NA | 6 (1.2) |
| Metastatic at diagnosis |  |  |  |  |  |  |  |  |  |  |
| Metastatic | NA | 2,064 (28) | 2,632 (31) | 255 (25) | 394 (30) | NA | 2,036 (77) | 2,604 (79) | 253 (71) | 392 (77) |
| Not Metastatic | NA | 3,005 (41) | 3,531 (41) | 491 (49) | 624 (47) | NA | 616 (23) | 677 (21) | 101 (29) | 120 (23) |
| Unknown | NA | 2,219 (30) | 2,379 (28) | 266 (26) | 307 (23) | NA | 0 (0) | 0 (0) | 0 (0) | 0 (0) |
| Abbreviations: AJCC, American Joint Committee on Cancer; CGDB, Flatiron Health-Foundation Medicine Clinico-Genomic Database; FHRD, Flatiron Health Research Database; NA, not applicable; SEER, The Surveillance, Epidemiology, and End Results research database. |  |  |  |  |  |  |  |  |  |  |

<sup>a</sup>Proportions of patients reported in the SEER cohort are not directly comparable to the FHRD and CGDB cohorts given that SEER represents all patients diagnosed with bladder cancer whereas the Urothelial Cancer FHRD and Bladder Cancer CGDB require patients to have 1) stage IV or node positive bladder or urinary tract cancer at initial diagnosis or 2) early-stage bladder or urinary tract cancer and subsequently developed advanced disease for inclusion.
Note: Reported percentages may not equal exactly to 100% due to rounding.

**Table 5.** Early Breast Cancer – Comparison of Demographic and Clinical Characteristics Between SEER vs FHRD vs CGDB^a^.

| Characteristic | All eligible, n (%) |  |  |  |  |
| --- | --- | --- | --- | --- | --- |
|  | SEER | FHRD |  | CGDB |  |
| Initial diagnosis | 2011–18 | 2011–18 | 2011–20 | 2011–18 | 2011–20 |
|  | N=468,001 | N=8,797 | N=10,541 | N=3,691 | N=4,026 |
| **Age at initial diagnosis (years)** |  |  |  |  |  |
| 0–19 | 28 (<0.1) | <5 (<0.1) | <5 (<0.1) | <5 (<0.2) | <5 (<0.2) |
| 20–34 | 8,367 (1.8) | 141 (1.6) | 162 (1.5) | 201 (5.4) | 215 (5.3) |
| 35–49 | 81,765 (17) | 1,475 (17) | 1,739 (16) | 1,107 (30) | 1,204 (30) |
| 50–64 | 173,112 (37) | 3,228 (37) | 3,824 (36) | 1,532 (42) | 1,647 (41) |
| 65–79 | 157,540 (34) | 3,159 (36) | 3,860 (37) | 756 (20) | 850 (21) |
| 80+ | 47,189 (10) | 793 (9.0) | 955 (9.1) | 95 (2.6) | 110 (2.7) |
| **Sex** |  |  |  |  |  |
| Female | 464,502 (99) | 8,722 (99) | 10,454 (99) | 3,648 (99) | 3,977 (99) |
| Male | 3,499 (0.7) | 74 (0.8) | 86 (0.8) | 43 (1.2) | 49 (1.2) |
| Unknown | 0 (0) | <5 (<0.1) | <5 (<0.1) | 0 (0) | 0 (0) |
| **Race** |  |  |  |  |  |
| Asian | 42,225 (9.0) | 244 (2.8) | 294 (2.8) | 85 (2.3) | 92 (2.3) |
| Black | 50,898 (11) | 789 (9.0) | 972 (9.2) | 397 (11) | 444 (11) |
| Other | 2,821 (0.6) | 1,158 (13) | 1,428 (14) | 549 (15) | 604 (15) |
| Unknown | 3,130 (0.7) | 716 (8.1) | 924 (8.8) | 283 (7.7) | 326 (8.1) |
| White | 368,927 (79) | 5,890 (67) | 6,923 (66) | 2,377 (64) | 2,560 (64) |
| **AJCC stage at diagnosis** |  |  |  |  |  |
| Stage 0 | NA | NA | NA | <5 (<0.2) | <5 (<0.2) |
| Stage I | 259,391 (55) | 4,360 (50) | 5,496 (52) | 680 (18) | 748 (19) |
| Stage II | 158,583 (34) | 2,693 (31) | 2,988 (28) | 1,687 (46) | 1,794 (45) |
| Stage III | 50,027 (11) | 888 (10) | 1,043 (9.9) | 1,323 (36) | 1,483 (37) |
| Unknown | NA | 856 (9.7) | 1,014 (9.6) | NA | NA |
| **Year of initial diagnosis** |  |  |  |  |  |
| 2011 | 55,007 (12) | 1,026 (12) | 1,026 (9.7) | 423 (11) | 423 (11) |
| 2012 | 56,280 (12) | 1,011 (11) | 1,011 (9.6) | 441 (12) | 441 (11) |
| 2013 | 57,511 (12) | 1,110 (13) | 1,110 (11) | 483 (13) | 483 (12) |
| 2014 | 58,572 (13) | 1,162 (13) | 1,162 (11) | 502 (14) | 502 (12) |
| 2015 | 60,624 (13) | 1,143 (13) | 1,143 (11) | 571 (15) | 571 (14) |
| 2016 | 60,952 (13) | 1,172 (13) | 1,172 (11) | 525 (14) | 525 (13) |
| 2017 | 61,916 (13) | 1,079 (12) | 1,079 (10) | 418 (11) | 418 (10) |
| 2018 | 57,139 (12) | 1,094 (12) | 1,094 (10) | 328 (8.9) | 328 (8.1) |
| 2019 | NA | NA | 1,013 (9.6) | NA | 222 (5.5) |
| 2020 | NA | NA | 700 (6.6) | NA | 109 (2.7) |
| 2021 | NA | NA | 31 (0.3) | NA | <5 (<0.1) |

Abbreviations: AJCC, American Joint Committee on Cancer; CGDB, Flatiron Health-Foundation Medicine Clinico-Genomic Database; FHRD, Flatiron Health Research Database; NA, not applicable; SEER, The Surveillance, Epidemiology, and End Results research database.
<sup>a</sup>Proportion of FHRD patients reported by AJCC stage at diagnosis is not directly comparable to the SEER and CGDB cohorts given that patients with stage I-III explicitly documented and patients with documented TNM stage equivalent to stage I-III disease without stage explicitly documented are included in the FHRD cohort whereas the SEER cohort includes patients with Stage I-III disease only, and the CGDB cohort includes patients with stage I-III disease explicitly documented only for inclusion.
Note: Reported percentages may not equal exactly to 100% due to rounding.

**Table 6.** Metastatic Breast Cancer – Comparison of Demographic and Clinical Characteristics Between SEER (distant at diagnosis) vs FHRD (metastatic at any time) vs CGDB (metastatic at any time)^a^.

| Characteristic | All eligible, n (%) |  |  |  |  | Stage IV at diagnosis, n (%) |  |  |  |  |
| --- | --- | --- | --- | --- | --- | --- | --- | --- | --- | --- |
|  | SEER | FHRD |  | CGDB |  | SEER | FHRD |  | CGDB |  |
| Initial diagnosis | 2011–18 | 2011–18 | 2011–20 | 2011–18 | 2011–20 | 2011–18 | 2011–18 | 2011–20 | 2011–18 | 2011–20 |
|  | N=31,201 | N=14,816 | N=16,674 | N=4,361 | N=4,973 | N=29,979 | N=6,235 | N=7,797 | N=1,384 | N=1,820 |
| **Age at initial diagnosis (years)** |  |  |  |  |  |  |  |  |  |  |
| 0–19 | 5 (<0.1) | <5 (<0.1) | <5 (<0.1) | <5 (<0.2) | <5 (<0.1) | <5 (<0.1) | <5 (<0.1) | <5 (<0.1) | <5 (<0.4) | <5 (<0.3) |
| 20–34 | 872 (2.8) | 492 (3.3) | 558 (3.3) | 241 (5.5) | 267 (5.4) | 824 (2.7) | 171 (2.7) | 218 (2.8) | 78 (5.6) | 93 (5.1) |
| 35–49 | 4,815 (15) | 2,823 (19) | 3,112 (19) | 1,220 (28) | 1,349 (27) | 4,550 (15) | 908 (15) | 1,126 (14) | 314 (23) | 386 (21) |
| 50–64 | 11,407 (37) | 5,507 (37) | 6,130 (37) | 1,865 (43) | 2,100 (42) | 10,937 (36) | 2,209 (35) | 2,731 (35) | 591 (43) | 769 (42) |
| 65–79 | 9,739 (31) | 5,402 (36) | 6,043 (36) | 912 (21) | 1,099 (22) | 9,407 (31) | 2,587 (41) | 3,149 (40) | 346 (25) | 490 (27) |
| 80+ | 4,363 (14) | 592 (4.0) | 831 (5.0) | 123 (2.8) | 158 (3.2) | 4,258 (14) | 360 (5.8) | 573 (7.3) | 55 (4.0) | 82 (4.5) |
| **Sex** |  |  |  |  |  |  |  |  |  |  |
| Female | 30,825 (99) | 14,615 (99) | 16,450 (99) | 4,305 (99) | 4,901 (99) | 29,610 (99) | 6,158 (99) | 7,699 (99) | 1,367 (99) | 1,791 (98) |
| Male | 376 (1.2) | 200 (1.3) | 223 (1.3) | 55 (1.3) | 71 (1.4) | 369 (1.2) | 76 (1.2) | 97 (1.2) | 16 (1.2) | 28 (1.5) |
| Unknown | 0 (0) | <5 (<0.1) | <5 (<0.1) | <5 (<0.1) | <5 (<0.1) | 0 (0) | <5 (<0.1) | <5 (<0.1) | <5 (<0.1) | <5 (<0.1) |
| **Race** |  |  |  |  |  |  |  |  |  |  |
| Asian | 2,349 (7.5) | 368 (2.5) | 412 (2.5) | 84 (1.9) | 95 (1.9) | 2,247 (7.5) | 144 (2.3) | 184 (2.4) | 18 (1.3) | 25 (1.4) |
| Black | 5,145 (16) | 1,904 (13) | 2,151 (13) | 476 (11) | 565 (11) | 4,926 (16) | 738 (12) | 944 (12) | 154 (11) | 218 (12) |
| Other | 212 (0.7) | 1,696 (11) | 1,934 (12) | 633 (15) | 755 (15) | 204 (0.7) | 703 (11) | 906 (12) | 200 (14) | 299 (16) |
| Unknown | 156 (0.5) | 1,418 (9.6) | 1,712 (10) | 320 (7.3) | 391 (7.9) | 143 (0.5) | 676 (11) | 921 (12) | 107 (7.7) | 149 (8.2) |
| White | 23,339 (75) | 9,430 (64) | 10,465 (63) | 2,848 (65) | 3,167 (64) | 22,459 (75) | 3,974 (64) | 4,842 (62) | 905 (65) | 1,129 (62) |

| AJCC stage at diagnosis |  |  |  |  |  |  |  |  |  |  |
| --- | --- | --- | --- | --- | --- | --- | --- | --- | --- | --- |
| Stage 0 | <5 (<0.1) | <5 (<0.1) | <5 (<0.1) | <5 (<0.1) | <5 (<0.1) | NA | NA | NA | NA | NA |
| Stage I | 17 (<0.1) | 1,197 (8.1) | 1,229 (7.4) | 438 (10) | 457 (9.2) | NA | NA | NA | NA | NA |
| Stage II | 43 (0.1) | 3,323 (22) | 3,411 (20) | 1,241 (28) | 1,294 (26) | NA | NA | NA | NA | NA |
| Stage III | 907 (2.9) | 3,191 (22) | 3,317 (20) | 1,041 (24) | 1,131 (23) | NA | NA | NA | NA | NA |
| Stage IV | 29,979 (96) | 6,235 (42) | 7,797 (47) | 1,384 (32) | 1,820 (37) | NA | NA | NA | NA | NA |
| Unknown | 254 (0.8) | 869 (5.9) | 919 (5.5) | 256 (5.9) | 270 (5.4) | NA | NA | NA | NA | NA |
| Year of initial diagnosis |  |  |  |  |  |  |  |  |  |  |
| 2011 | 3,601 (12) | 1,968 (13) | 1,968 (12) | 426 (9.8) | 426 (8.6) | 3,474 (12) | 603 (9.7) | 603 (7.7) | 62 (4.5) | 62 (3.4) |
| 2012 | 3,640 (12) | 2,058 (14) | 2,058 (12) | 513 (12) | 513 (10) | 3,519 (12) | 690 (11) | 690 (8.8) | 117 (8.5) | 117 (6.4) |
| 2013 | 3,869 (12) | 2,024 (14) | 2,024 (12) | 531 (12) | 531 (11) | 3,754 (13) | 766 (12) | 766 (9.8) | 125 (9.0) | 125 (6.9) |
| 2014 | 3,914 (13) | 2,053 (14) | 2,053 (12) | 642 (15) | 642 (13) | 3,792 (13) | 819 (13) | 819 (11) | 203 (15) | 203 (11) |
| 2015 | 3,860 (12) | 1,953 (13) | 1,953 (12) | 632 (14) | 632 (13) | 3,735 (12) | 823 (13) | 823 (11) | 199 (14) | 199 (11) |
| 2016 | 4,022 (13) | 1,826 (12) | 1,826 (11) | 623 (14) | 623 (13) | 3,842 (13) | 871 (14) | 871 (11) | 227 (16) | 227 (12) |
| 2017 | 4,090 (13) | 1,620 (11) | 1,620 (9.7) | 524 (12) | 524 (11) | 3,827 (13) | 830 (13) | 830 (11) | 207 (15) | 207 (11) |
| 2018 | 4,205 (13) | 1,314 (8.9) | 1,314 (7.9) | 470 (11) | 470 (9.5) | 4,036 (13) | 833 (13) | 833 (11) | 244 (18) | 244 (13) |
| 2019 | NA | NA | 1,050 (6.3) | NA | 332 (6.7) | NA | NA | 808 (10) | NA | 196 (11) |
| 2020 | NA | NA | 685 (4.1) | NA | 259 (5.2) | NA | NA | 632 (8.1) | NA | 219 (12) |
| 2021 | NA | NA | 123 (0.7) | NA | 21 (0.4) | NA | NA | 122 (1.6) | NA | 21 (1.2) |
| Abbreviations: CGDB, Flatiron Health-Foundation Medicine Clinico-Genomic Database; FHRD, Flatiron Health Research Database; NA, not applicable; SEER, The Surveillance, Epidemiology, and End Results research database. |  |  |  |  |  |  |  |  |  |  |
| <sup>a</sup>Proportions of patients reported in the SEER cohort are not directly comparable to the FHRD and CGDB cohorts given that the SEER cohort represents all patients with distant breast cancer at initial diagnosis whereas the breast cancer FHRD and CGDB represent patients diagnosed with Stage IV disease at initial diagnosis and patients diagnosed with earlier stage disease who subsequently developed metastasis. Distant disease in SEER is defined as patients with Localized/Regional/Distant (LRD) Summary stage (2004+) variable = "Distant" at the time of diagnosis only. |  |  |  |  |  |  |  |  |  |  |
| Note: Reported percentages may not equal exactly to 100% due to rounding. |  |  |  |  |  |  |  |  |  |  |

**Table 7.** Chronic Lymphocytic Leukemia – Comparison of Demographic and Clinical Characteristics Between SEER vs FHRD vs CGDB.

| Characteristic | All eligible, n (%) |  |  |  |  |
| --- | --- | --- | --- | --- | --- |
|  | SEER | FHRD |  | CGDB |  |
| Initial diagnosis | 2011–18 | 2011–18 | 2011–20 | 2011–18 | 2011–20 |
|  | N=37,782 | N=6,367 | N=7,052 | N=81 | N=91 |
| **Age at initial diagnosis (years)** |  |  |  |  |  |
| 0–19 | 19 (<0.1) | <5 (<0.1) | <5 (<0.1) | <5 (<6.2) | <5 (<5.5) |
| 20–34 | 98 (0.3) | 13 (0.2) | 16 (0.2) | <5 (<6.2) | <5 (<5.5) |
| 35–49 | 1,608 (4.3) | 309 (4.9) | 335 (4.8) | <5 (<6.2) | <5 (<5.5) |
| 50–64 | 10,744 (28) | 1,987 (31) | 2,175 (31) | 40 (49) | 45 (49) |
| 65–79 | 16,434 (43) | 2,952 (46) | 3,284 (47) | 27 (33) | 29 (32) |
| 80+ | 8,879 (24) | 1,106 (17) | 1,242 (18) | 9 (11) | 11 (12) |
| **Sex** |  |  |  |  |  |
| Female | 14,845 (39) | 2,253 (35) | 2,489 (35) | 28 (35) | 33 (36) |
| Male | 22,937 (61) | 4,114 (65) | 4,563 (65) | 53 (65) | 58 (64) |
| **Race** |  |  |  |  |  |
| Asian | 874 (2.3) | 58 (0.9) | 66 (0.9) | <5 (<6.2) | <5 (<5.5) |
| Black | 2,773 (7.3) | 520 (8.2) | 578 (8.2) | 9 (11) | 9 (9.9) |
| Other | 117 (0.3) | 514 (8.1) | 597 (8.5) | 23 (28) | 26 (29) |
| Unknown | 1,225 (3.2) | 520 (8.2) | 620 (8.8) | <5 (<6.2) | <5 (<5.5) |
| White | 32,793 (87) | 4,755 (75) | 5,191 (74) | 45 (56) | 51 (56) |
| **Rai stage at diagnosis** |  |  |  |  |  |
| Stage 0 | NA | 1,273 (20) | 1,325 (19) | 6 (7.4) | 7 (7.7) |
| Stage I | NA | 862 (14) | 934 (13) | 17 (21) | 19 (21) |
| Stage II | NA | 419 (6.6) | 479 (6.8) | 10 (12) | 10 (11) |
| Stage III | NA | 386 (6.1) | 453 (6.4) | <5 (<6.2) | 6 (6.6) |
| Stage IV | NA | 680 (11) | 798 (11) | 14 (17) | 15 (16) |
| Unknown | NA | 2,747 (43) | 3,063 (43) | 31 (38) | 34 (37) |
| **Year of initial diagnosis** |  |  |  |  |  |
| 2011 | 4,625 (12) | 780 (12) | 780 (11) | 11 (14) | 11 (12) |
| 2012 | 4,698 (12) | 800 (13) | 800 (11) | 10 (12) | 10 (11) |
| 2013 | 4,776 (13) | 884 (14) | 884 (13) | 9 (11) | 9 (9.9) |
| 2014 | 4,864 (13) | 876 (14) | 876 (12) | 15 (19) | 15 (16) |
| 2015 | 4,889 (13) | 855 (13) | 855 (12) | 10 (12) | 10 (11) |
| 2016 | 4,690 (12) | 837 (13) | 837 (12) | 9 (11) | 9 (9.9) |
| 2017 | 4,859 (13) | 713 (11) | 713 (10) | 11 (14) | 11 (12) |
| 2018 | 4,381 (12) | 622 (9.8) | 622 (8.8) | 6 (7.4) | 6 (6.6) |
| 2019 | NA | NA | 458 (6.5) | NA | <5 (<5.5) |
| 2020 | NA | NA | 218 (3.1) | NA | 5 (5.5) |
| 2021 | NA | NA | 9 (0.1) | NA | <5 (<5.5) |

Abbreviations: CGDB, Flatiron Health-Foundation Medicine Clinico-Genomic Database; FHRD, Flatiron Health Research Database; NA, not applicable; SEER, The Surveillance, Epidemiology, and End Results research database.
Note: Reported percentages may not equal exactly to 100% due to rounding.

**Table 8.** Colorectal Cancer – Comparison of Demographic and Clinical Characteristics Between SEER vs FHRD (metastatic-only) vs CGDB^a,b^.

| Characteristic | All eligible, n (%) |  |  |  |  | Stage IV at diagnosis, n (%) |  |  |  |  |
| --- | --- | --- | --- | --- | --- | --- | --- | --- | --- | --- |
|  | SEER | FHRD |  | CGDB |  | SEER | FHRD |  | CGDB |  |
| Initial diagnosis | 2011–18 | 2011–18 | 2011–20 | 2011–18 | 2011–20 | 2011–18 | 2011–18 | 2011–20 | 2011–18 | 2011–20 |
|  | N=291,724 | N=22,047 | N=26,529 | N=6,663 | N=8,367 | N=58,019 | N=12,196 | N=15,974 | N=3,338 | N=4,534 |
| **Age at initial diagnosis (years)** |  |  |  |  |  |  |  |  |  |  |
| 0–19 | 113 (<0.1) | 9 (<0.1) | 9 (<0.1) | <5 (<0.1) | <5 (<0.1) | 28 (<0.1) | 6 (<0.1) | 6 (<0.1) | <5 (<0.2) | <5 (<0.2) |
| 20–34 | 3,872 (1.3) | 376 (1.7) | 456 (1.7) | 184 (2.8) | 221 (2.6) | 1,052 (1.8) | 228 (1.9) | 295 (1.8) | 109 (3.3) | 131 (2.9) |
| 35–49 | 27,528 (9.4) | 2,920 (13) | 3,508 (13) | 1,294 (19) | 1,584 (19) | 7,051 (12) | 1,721 (14) | 2,241 (14) | 703 (21) | 923 (20) |
| 50–64 | 95,102 (33) | 7,919 (36) | 9,461 (36) | 2,921 (44) | 3,551 (42) | 19,771 (34) | 4,442 (36) | 5,765 (36) | 1,496 (45) | 1,944 (43) |
| 65–79 | 103,658 (36) | 8,145 (37) | 9,830 (37) | 1,942 (29) | 2,531 (30) | 19,395 (33) | 4,333 (36) | 5,746 (36) | 897 (27) | 1,305 (29) |
| 80+ | 61,451 (21) | 2,678 (12) | 3,265 (12) | 321 (4.8) | 479 (5.7) | 10,722 (18) | 1,466 (12) | 1,921 (12) | 133 (4.0) | 231 (5.1) |
| **Sex** |  |  |  |  |  |  |  |  |  |  |
| Female | 138,215 (47) | 9,984 (45) | 11,987 (45) | 3,063 (46) | 3,854 (46) | 26,424 (46) | 5,576 (46) | 7,273 (46) | 1,569 (47) | 2,118 (47) |
| Male | 153,509 (53) | 12,060 (55) | 14,539 (55) | 3,600 (54) | 4,513 (54) | 31,595 (54) | 6,618 (54) | 8,699 (54) | 1,769 (53) | 2,416 (53) |
| Unknown | 0 (0) | <5 (<0.1) | <5 (<0.1) | 0 (0) | 0 (0) | 0 (0) | <5 (<0.1) | <5 (<0.1) | 0 (0) | 0 (0) |
| **Race** |  |  |  |  |  |  |  |  |  |  |
| Asian | 25,265 (8.7) | 648 (2.9) | 796 (3.0) | 177 (2.7) | 233 (2.8) | 4,572 (7.9) | 324 (2.7) | 453 (2.8) | 79 (2.4) | 116 (2.6) |
| Black | 36,107 (12) | 2,337 (11) | 2,821 (11) | 531 (8.0) | 690 (8.2) | 8,648 (15) | 1,312 (11) | 1,719 (11) | 269 (8.1) | 384 (8.5) |
| Other | 2,449 (0.8) | 2,612 (12) | 3,205 (12) | 961 (14) | 1,276 (15) | 519 (0.9) | 1,386 (11) | 1,900 (12) | 475 (14) | 701 (15) |
| Unknown | 3,158 (1.1) | 2,089 (9.5) | 2,782 (10) | 511 (7.7) | 686 (8.2) | 179 (0.3) | 1,240 (10) | 1,851 (12) | 249 (7.5) | 374 (8.2) |
| White | 224,745 (77) | 14,361 (65) | 16,925 (64) | 4,483 (67) | 5,482 (66) | 44,101 (76) | 7,934 (65) | 10,051 (63) | 2,266 (68) | 2,959 (65) |

| AJCC stage at diagnosis |  |  |  |  |  |  |  |  |  |  |
| --- | --- | --- | --- | --- | --- | --- | --- | --- | --- | --- |
| Stage 0 | 8,055 (2.8) | <5 (<0.1) | <5 (<0.1) | <5 (<0.1) | <5 (<0.1) | NA | NA | NA | NA | NA |
| Stage I | 60,878 (21) | 577 (2.6) | 594 (2.2) | 204 (3.1) | 223 (2.7) | NA | NA | NA | NA | NA |
| Stage II | 62,781 (22) | 2,585 (12) | 2,778 (10) | 820 (12) | 934 (11) | NA | NA | NA | NA | NA |
| Stage III | 70,130 (24) | 5,904 (27) | 6,349 (24) | 2,043 (31) | 2,373 (28) | NA | NA | NA | NA | NA |
| Stage IV | 58,019 (20) | 12,196 (55) | 15,974 (60) | 3,338 (50) | 4,534 (54) | NA | NA | NA | NA | NA |
| Unknown | 31,861 (11) | 785 (3.6) | 834 (3.1) | 257 (3.9) | 302 (3.6) | NA | NA | NA | NA | NA |
| Year of initial diagnosis |  |  |  |  |  |  |  |  |  |  |
| 2011 | 36,242 (12) | 750 (3.4) | 750 (2.8) | 327 (4.9) | 327 (3.9) | 6,996 (12) | NA | NA | 110 (3.3) | 110 (2.4) |
| 2012 | 36,286 (12) | 1,295 (5.9) | 1,295 (4.9) | 499 (7.5) | 499 (6.0) | 6,962 (12) | NA | NA | 178 (5.3) | 178 (3.9) |
| 2013 | 35,956 (12) | 3,314 (15) | 3,314 (12) | 584 (8.8) | 584 (7.0) | 7,031 (12) | 1,851 (15) | 1,851 (12) | 265 (7.9) | 265 (5.8) |
| 2014 | 37,210 (13) | 3,479 (16) | 3,479 (13) | 785 (12) | 785 (9.4) | 7,346 (13) | 2,037 (17) | 2,037 (13) | 360 (11) | 360 (7.9) |
| 2015 | 36,691 (13) | 3,550 (16) | 3,550 (13) | 990 (15) | 990 (12) | 7,232 (12) | 2,109 (17) | 2,109 (13) | 474 (14) | 474 (10) |
| 2016 | 36,870 (13) | 3,412 (15) | 3,412 (13) | 1,117 (17) | 1,117 (13) | 7,476 (13) | 2,097 (17) | 2,097 (13) | 577 (17) | 577 (13) |
| 2017 | 36,239 (12) | 3,228 (15) | 3,228 (12) | 1,160 (17) | 1,160 (14) | 7,444 (13) | 2,034 (17) | 2,034 (13) | 652 (20) | 652 (14) |
| 2018 | 36,230 (12) | 3,019 (14) | 3,019 (11) | 1,201 (18) | 1,201 (14) | 7,532 (13) | 2,068 (17) | 2,068 (13) | 722 (22) | 722 (16) |
| 2019 | NA | NA | 2,622 (9.9) | NA | 979 (12) | NA | NA | 2,029 (13) | NA | 642 (14) |
| 2020 | NA | NA | 1,734 (6.5) | NA | 665 (7.9) | NA | NA | 1,623 (10) | NA | 505 (11) |
| 2021 | NA | NA | 126 (0.5) | NA | 60 (0.7) | NA | NA | 126 (0.8) | NA | 49 (1.1) |

Abbreviations: CGDB, Flatiron Health-Foundation Medicine Clinico-Genomic Database; FHRD, Flatiron Health Research Database; NA, not applicable; SEER, The Surveillance, Epidemiology, and End Results research database.
\*Proportions of patients reported in the FHRD cohort are not directly comparable to the SEER and CGDB cohorts given that patients with metastatic disease (at initial diagnosis or through the subsequent development of metastatic disease) are included in the Colorectal Cancer FHRD whereas SEER and the Colorectal Cancer CGDB include patients of all stages regardless of metastatic disease status.
\*Proportion of FHRD patients reported by Year of initial diagnosis is not directly comparable to the SEER and CGDB cohorts given that the diagnosis year for inclusion began in 2013 for patients with stage IV disease, and metastatic diagnosis year for inclusion began in 2013 for patients who presented with earlier stage CRC and subsequently developed metastatic disease.
Note: Reported percentages may not equal exactly to 100% due to rounding.

**Table 9.** Diffuse Large B–Cell Lymphoma – Comparison of Demographic and Clinical Characteristics Between SEER vs FHRD vs CGDB.

| Characteristic | All eligible, n (%) |  |  |  |  | Stage IV at diagnosis, n (%) |  |  |  |  |
| --- | --- | --- | --- | --- | --- | --- | --- | --- | --- | --- |
|  | SEER | FHRD |  | CGDB |  | SEER | FHRD |  | CGDB |  |
| Initial diagnosis | 2011–18 | 2011–18 | 2011–20 | 2011–18 | 2011–20 | 2011–18 | 2011–18 | 2011–20 | 2011–18 | 2011–20 |
|  | N=53,690 | N=6,130 | N=7,689 | N=155 | N=185 | N=18,396 | N=1,735 | N=2,157 | N=58 | N=63 |
| **Age at initial diagnosis (years)** |  |  |  |  |  |  |  |  |  |  |
| 0–19 | 489 (0.9) | 14 (0.2) | 19 (0.2) | <5 (<3.2) | <5 (<2.7) | 128 (0.7) | <5 (<0.1) | <5 (<0.1) | <5 (<8.6) | <5 (<7.9) |
| 20–34 | 1,937 (3.6) | 240 (3.9) | 279 (3.6) | 13 (8.4) | 13 (7.0) | 630 (3.4) | 63 (3.6) | 72 (3.3) | <5 (<8.6) | <5 (<7.9) |
| 35–49 | 4,855 (9.0) | 518 (8.5) | 617 (8.0) | 22 (14) | 24 (13) | 1,632 (8.9) | 145 (8.4) | 176 (8.2) | 8 (14) | 8 (13) |
| 50–64 | 14,493 (27) | 1,686 (28) | 2,064 (27) | 41 (26) | 50 (27) | 5,132 (28) | 491 (28) | 603 (28) | 18 (31) | 22 (35) |
| 65–79 | 20,705 (39) | 2,524 (41) | 3,248 (42) | 65 (42) | 79 (43) | 7,283 (40) | 916 (53) | 1,121 (52) | 25 (43) | 26 (41) |
| 80+ | 11,211 (21) | 1,148 (19) | 1,462 (19) | 14 (9.0) | 19 (10) | 3,591 (20) | 118 (6.8) | 182 (8.4) | <5 (<8.6) | <5 (<7.9) |
| **Sex** |  |  |  |  |  |  |  |  |  |  |
| Female | 23,789 (44) | 2,785 (45) | 3,462 (45) | 52 (34) | 64 (35) | 7,943 (43) | 757 (44) | 937 (43) | 19 (33) | 20 (32) |
| Male | 29,901 (56) | 3,344 (55) | 4,226 (55) | 103 (66) | 121 (65) | 10,453 (57) | 978 (56) | 1,220 (57) | 39 (67) | 43 (68) |
| Unknown | 0 (0) | <5 (<0.1) | <5 (<0.1) | 0 (0) | 0 (0) | 0 (0) | 0 (0) | 0 (0) | 0 (0) | 0 (0) |
| **Race** |  |  |  |  |  |  |  |  |  |  |
| Asian | 4,804 (8.9) | 137 (2.2) | 176 (2.3) | <5 (<3.2) | 5 (2.7) | 1,532 (8.3) | 32 (1.8) | 43 (2.0) | <5 (<8.6) | <5 (<7.9) |
| Black | 3,841 (7.2) | 391 (6.4) | 484 (6.3) | 5 (3.2) | 6 (3.2) | 1,509 (8.2) | 142 (8.2) | 174 (8.1) | <5 (<8.6) | <5 (<7.9) |
| Other | 311 (0.6) | 732 (12) | 988 (13) | 24 (15) | 31 (17) | 110 (0.6) | 196 (11) | 269 (12) | 9 (16) | 9 (14) |
| Unknown | 414 (0.8) | 590 (9.6) | 767 (10.0) | 19 (12) | 20 (11) | 84 (0.5) | 149 (8.6) | 200 (9.3) | 6 (10) | 6 (9.5) |
| White | 44,320 (83) | 4,280 (70) | 5,274 (69) | 103 (66) | 123 (66) | 15,161 (82) | 1,216 (70) | 1,471 (68) | 41 (71) | 44 (70) |

| AJCC stage at diagnosis |  |  |  |  |  |  |  |  |  |  |
| --- | --- | --- | --- | --- | --- | --- | --- | --- | --- | --- |
| Stage I | 12,666 (24) | 736 (12) | 887 (12) | 15 (9.7) | 17 (9.2) | NA | NA | NA | NA | NA |
| Stage II | 9,639 (18) | 957 (16) | 1,186 (15) | 21 (14) | 26 (14) | NA | NA | NA | NA | NA |
| Stage III | 9,162 (17) | 1,117 (18) | 1,403 (18) | 16 (10) | 20 (11) | NA | NA | NA | NA | NA |
| Stage IV | 18,396 (34) | 1,735 (28) | 2,157 (28) | 58 (37) | 63 (34) | NA | NA | NA | NA | NA |
| Unknown | 3,827 (7.1) | 1,585 (26) | 2,056 (27) | 45 (29) | 59 (32) | NA | NA | NA | NA | NA |
| Year of initial diagnosis |  |  |  |  |  |  |  |  |  |  |
| 2011 | 6,212 (12) | 591 (9.6) | 591 (7.7) | 8 (5.2) | 8 (4.3) | 2,101 (11) | 163 (9.4) | 163 (7.6) | <5 (<8.6) | <5 (<7.9) |
| 2012 | 6,515 (12) | 693 (11) | 693 (9.0) | 5 (3.2) | 5 (2.7) | 2,070 (11) | 207 (12) | 207 (9.6) | <5 (<8.6) | <5 (<7.9) |
| 2013 | 6,492 (12) | 749 (12) | 749 (9.7) | 18 (12) | 18 (9.7) | 2,208 (12) | 210 (12) | 210 (9.7) | 7 (12) | 7 (11) |
| 2014 | 6,709 (12) | 738 (12) | 738 (9.6) | 13 (8.4) | 13 (7.0) | 2,277 (12) | 189 (11) | 189 (8.8) | 6 (10) | 6 (9.5) |
| 2015 | 6,817 (13) | 829 (14) | 829 (11) | 23 (15) | 23 (12) | 2,352 (13) | 240 (14) | 240 (11) | 9 (16) | 9 (14) |
| 2016 | 6,890 (13) | 849 (14) | 849 (11) | 26 (17) | 26 (14) | 2,358 (13) | 241 (14) | 241 (11) | 10 (17) | 10 (16) |
| 2017 | 6,989 (13) | 864 (14) | 864 (11) | 35 (23) | 35 (19) | 2,392 (13) | 252 (15) | 252 (12) | 16 (28) | 16 (25) |
| 2018 | 7,066 (13) | 817 (13) | 817 (11) | 27 (17) | 27 (15) | 2,638 (14) | 233 (13) | 233 (11) | 7 (12) | 7 (11) |
| 2019 | NA | NA | 791 (10) | NA | 14 (7.6) | NA | NA | 223 (10) | NA | <5 (<7.9) |
| 2020 | NA | NA | 717 (9.3) | NA | 16 (8.6) | NA | NA | 190 (8.8) | NA | <5 (<7.9) |
| 2021 | NA | NA | 51 (0.7) | NA | <5 (<2.7) | NA | NA | 9 (0.4) | NA | <5 (<7.9) |
| Abbreviations: AJCC, American Joint Committee on Cancer; CGDB, Flatiron Health-Foundation Medicine Clinico-Genomic Database; FHRD, Flatiron Health Research Database; NA, not applicable; SEER, The Surveillance, Epidemiology, and End Results research database. |  |  |  |  |  |  |  |  |  |  |
| Note: Reported percentages may not equal exactly to 100% due to rounding. |  |  |  |  |  |  |  |  |  |  |

**Table 10.**
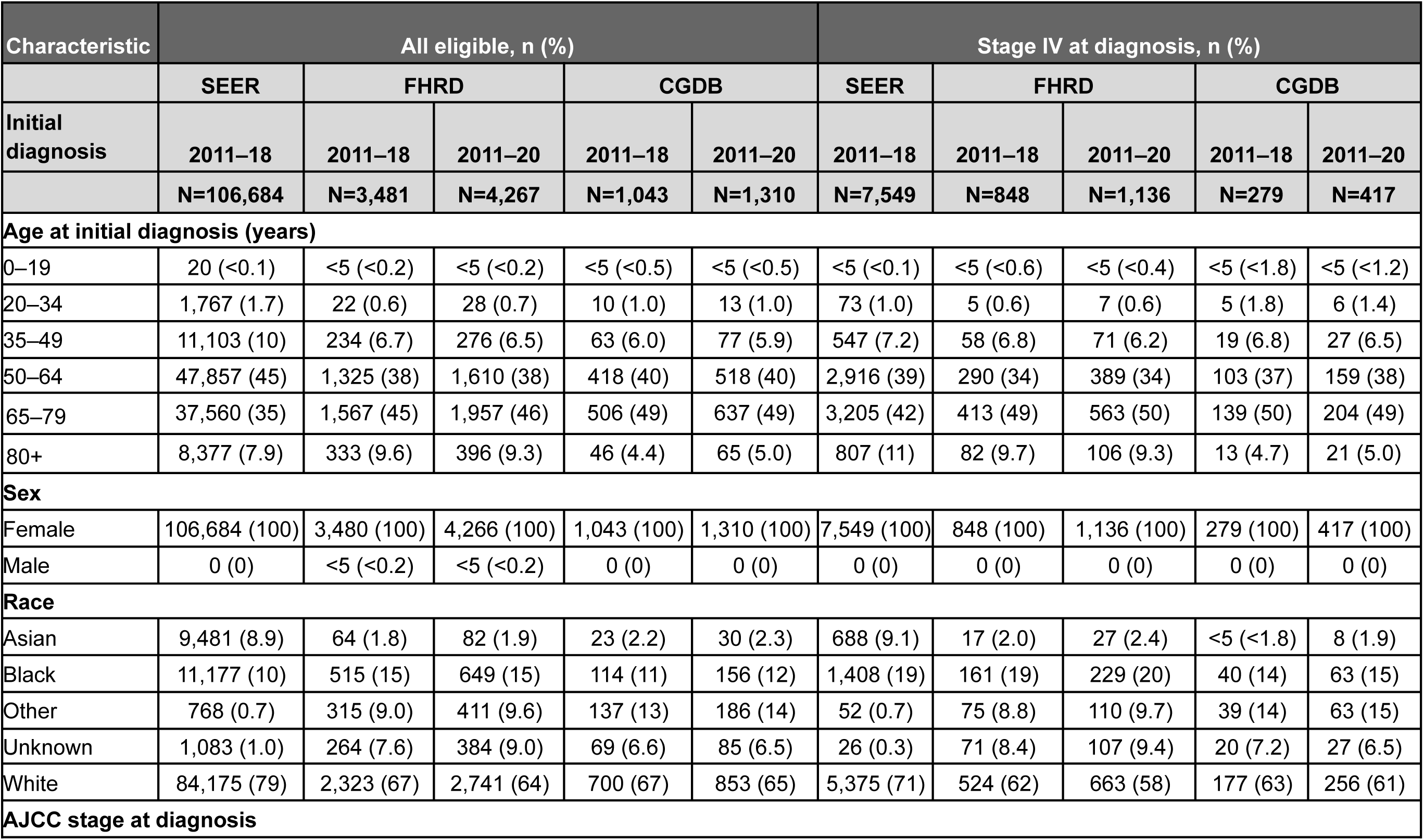
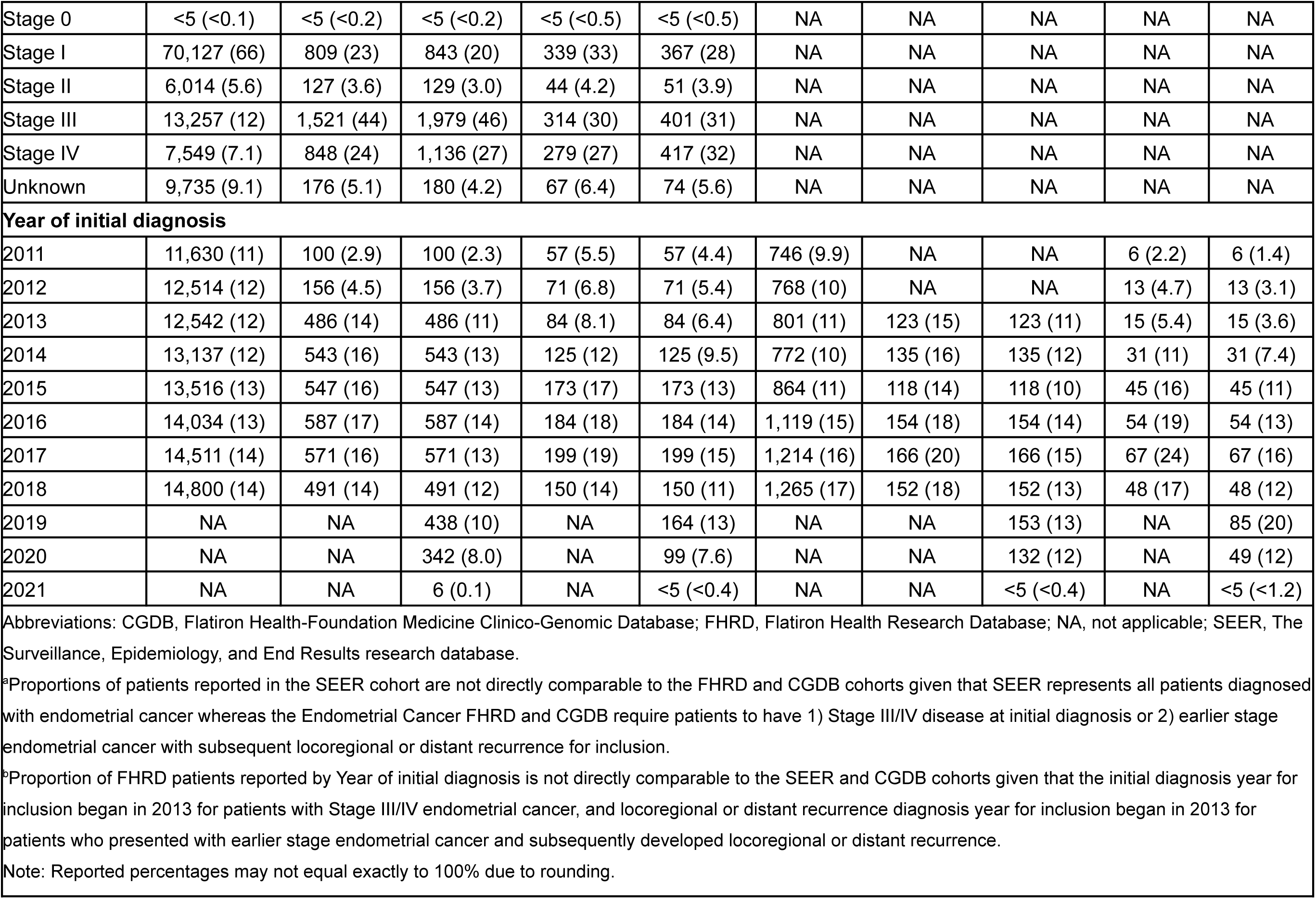
Endometrial Cancer – Comparison of Demographic and Clinical Characteristics Between SEER (advanced and non-advanced at diagnosis) vs FHRD (advanced at any time) vs CGDB (advanced at any time)^a,b^.

(advanced and non-advanced at diagnosis) vs FHRD (advanced at any time) vs CGDB (advanced at any time)<sup>a,b</sup>
| Characteristic | All eligible, n (%) |  |  |  |  | Stage IV at diagnosis, n (%) |  |  |  |  |
| --- | --- | --- | --- | --- | --- | --- | --- | --- | --- | --- |
|  | SEER | FHRD |  | CGDB |  | SEER | FHRD |  | CGDB |  |
| Initial diagnosis | 2011–18 | 2011–18 | 2011–20 | 2011–18 | 2011–20 | 2011–18 | 2011–18 | 2011–20 | 2011–18 | 2011–20 |
|  | N=106,684 | N=3,481 | N=4,267 | N=1,043 | N=1,310 | N=7,549 | N=848 | N=1,136 | N=279 | N=417 |
| Age at initial diagnosis (years) |  |  |  |  |  |  |  |  |  |  |
| 0–19 | 20 (<0.1) | <5 (<0.2) | <5 (<0.2) | <5 (<0.5) | <5 (<0.5) | <5 (<0.1) | <5 (<0.6) | <5 (<0.4) | <5 (<1.8) | <5 (<1.2) |
| 20–34 | 1,767 (1.7) | 22 (0.6) | 28 (0.7) | 10 (1.0) | 13 (1.0) | 73 (1.0) | 5 (0.6) | 7 (0.6) | 5 (1.8) | 6 (1.4) |
| 35–49 | 11,103 (10) | 234 (6.7) | 276 (6.5) | 63 (6.0) | 77 (5.9) | 547 (7.2) | 58 (6.8) | 71 (6.2) | 19 (6.8) | 27 (6.5) |
| 50–64 | 47,857 (45) | 1,325 (38) | 1,610 (38) | 418 (40) | 518 (40) | 2,916 (39) | 290 (34) | 389 (34) | 103 (37) | 159 (38) |
| 65–79 | 37,560 (35) | 1,567 (45) | 1,957 (46) | 506 (49) | 637 (49) | 3,205 (42) | 413 (49) | 563 (50) | 139 (50) | 204 (49) |
| 80+ | 8,377 (7.9) | 333 (9.6) | 396 (9.3) | 46 (4.4) | 65 (5.0) | 807 (11) | 82 (9.7) | 106 (9.3) | 13 (4.7) | 21 (5.0) |
| Sex |  |  |  |  |  |  |  |  |  |  |
| Female | 106,684 (100) | 3,480 (100) | 4,266 (100) | 1,043 (100) | 1,310 (100) | 7,549 (100) | 848 (100) | 1,136 (100) | 279 (100) | 417 (100) |
| Male | 0 (0) | <5 (<0.2) | <5 (<0.2) | 0 (0) | 0 (0) | 0 (0) | 0 (0) | 0 (0) | 0 (0) | 0 (0) |
| Race |  |  |  |  |  |  |  |  |  |  |
| Asian | 9,481 (8.9) | 64 (1.8) | 82 (1.9) | 23 (2.2) | 30 (2.3) | 688 (9.1) | 17 (2.0) | 27 (2.4) | <5 (<1.8) | 8 (1.9) |
| Black | 11,177 (10) | 515 (15) | 649 (15) | 114 (11) | 156 (12) | 1,408 (19) | 161 (19) | 229 (20) | 40 (14) | 63 (15) |
| Other | 768 (0.7) | 315 (9.0) | 411 (9.6) | 137 (13) | 186 (14) | 52 (0.7) | 75 (8.8) | 110 (9.7) | 39 (14) | 63 (15) |
| Unknown | 1,083 (1.0) | 264 (7.6) | 384 (9.0) | 69 (6.6) | 85 (6.5) | 26 (0.3) | 71 (8.4) | 107 (9.4) | 20 (7.2) | 27 (6.5) |
| White | 84,175 (79) | 2,323 (67) | 2,741 (64) | 700 (67) | 853 (65) | 5,375 (71) | 524 (62) | 663 (58) | 177 (63) | 256 (61) |
| AJCC stage at diagnosis |  |  |  |  |  |  |  |  |  |  |
| Stage 0 | <5 (<0.1) | <5 (<0.2) | <5 (<0.2) | <5 (<0.5) | <5 (<0.5) | NA | NA | NA | NA | NA |
| Stage I | 70,127 (66) | 809 (23) | 843 (20) | 339 (33) | 367 (28) | NA | NA | NA | NA | NA |
| Stage II | 6,014 (5.6) | 127 (3.6) | 129 (3.0) | 44 (4.2) | 51 (3.9) | NA | NA | NA | NA | NA |
| Stage III | 13,257 (12) | 1,521 (44) | 1,979 (46) | 314 (30) | 401 (31) | NA | NA | NA | NA | NA |
| Stage IV | 7,549 (7.1) | 848 (24) | 1,136 (27) | 279 (27) | 417 (32) | NA | NA | NA | NA | NA |
| Unknown | 9,735 (9.1) | 176 (5.1) | 180 (4.2) | 67 (6.4) | 74 (5.6) | NA | NA | NA | NA | NA |
| Year of initial diagnosis |  |  |  |  |  |  |  |  |  |  |
| 2011 | 11,630 (11) | 100 (2.9) | 100 (2.3) | 57 (5.5) | 57 (4.4) | 746 (9.9) | NA | NA | 6 (2.2) | 6 (1.4) |
| 2012 | 12,514 (12) | 156 (4.5) | 156 (3.7) | 71 (6.8) | 71 (5.4) | 768 (10) | NA | NA | 13 (4.7) | 13 (3.1) |
| 2013 | 12,542 (12) | 486 (14) | 486 (11) | 84 (8.1) | 84 (6.4) | 801 (11) | 123 (15) | 123 (11) | 15 (5.4) | 15 (3.6) |
| 2014 | 13,137 (12) | 543 (16) | 543 (13) | 125 (12) | 125 (9.5) | 772 (10) | 135 (16) | 135 (12) | 31 (11) | 31 (7.4) |
| 2015 | 13,516 (13) | 547 (16) | 547 (13) | 173 (17) | 173 (13) | 864 (11) | 118 (14) | 118 (10) | 45 (16) | 45 (11) |
| 2016 | 14,034 (13) | 587 (17) | 587 (14) | 184 (18) | 184 (14) | 1,119 (15) | 154 (18) | 154 (14) | 54 (19) | 54 (13) |
| 2017 | 14,511 (14) | 571 (16) | 571 (13) | 199 (19) | 199 (15) | 1,214 (16) | 166 (20) | 166 (15) | 67 (24) | 67 (16) |
| 2018 | 14,800 (14) | 491 (14) | 491 (12) | 150 (14) | 150 (11) | 1,265 (17) | 152 (18) | 152 (13) | 48 (17) | 48 (12) |
| 2019 | NA | NA | 438 (10) | NA | 164 (13) | NA | NA | 153 (13) | NA | 85 (20) |
| 2020 | NA | NA | 342 (8.0) | NA | 99 (7.6) | NA | NA | 132 (12) | NA | 49 (12) |
| 2021 | NA | NA | 6 (0.1) | NA | <5 (<0.4) | NA | NA | <5 (<0.4) | NA | <5 (<1.2) |

Abbreviations: CGDB, Flatiron Health-Foundation Medicine Clinico-Genomic Database; FHRD, Flatiron Health Research Database; NA, not applicable; SEER, The Surveillance, Epidemiology, and End Results research database.
\*Proportions of patients reported in the SEER cohort are not directly comparable to the FHRD and CGDB cohorts given that SEER represents all patients diagnosed with endometrial cancer whereas the Endometrial Cancer FHRD and CGDB require patients to have 1) Stage III/IV disease at initial diagnosis or 2) earlier stage endometrial cancer with subsequent locoregional or distant recurrence for inclusion.
<sup>b</sup>Proportion of FHRD patients reported by Year of initial diagnosis is not directly comparable to the SEER and CGDB cohorts given that the initial diagnosis year for inclusion began in 2013 for patients with Stage III/IV endometrial cancer, and locoregional or distant recurrence diagnosis year for inclusion began in 2013 for patients who presented with earlier stage endometrial cancer and subsequently developed locoregional or distant recurrence.
Note: Reported percentages may not equal exactly to 100% due to rounding.

**Table 11.**
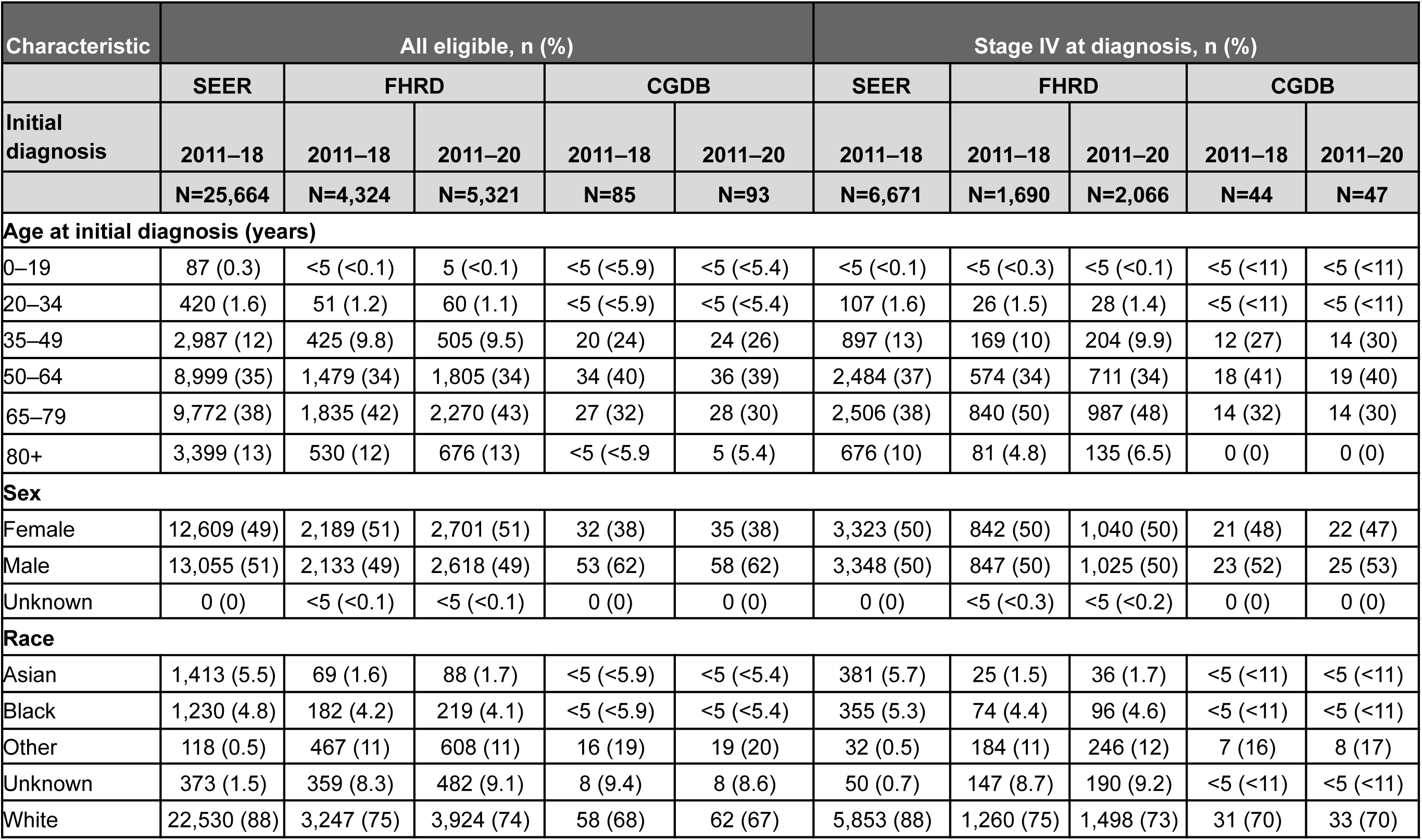
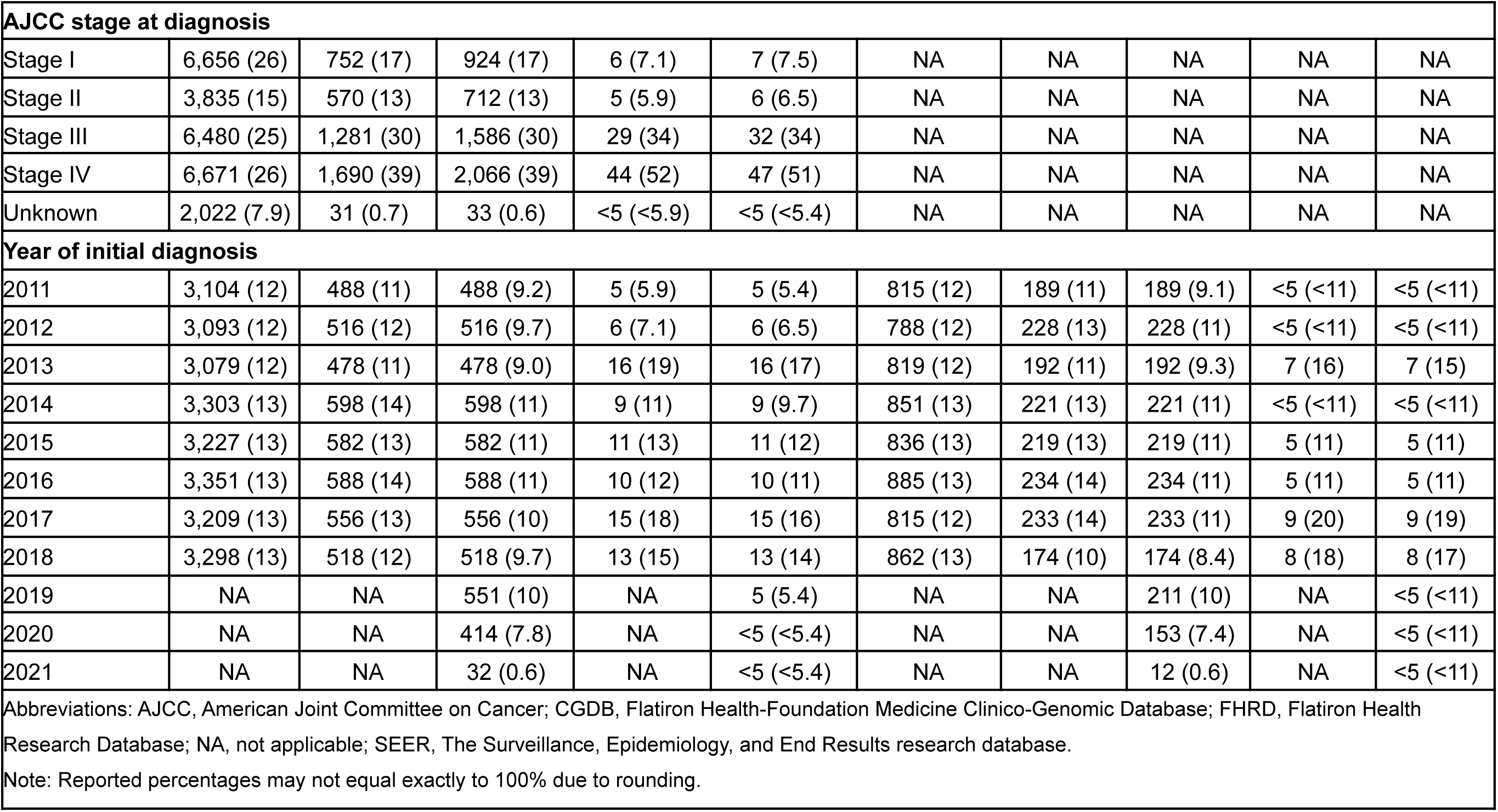
Follicular Lymphoma – Comparison of Demographic and Clinical Characteristics Between SEER vs FHRD vs CGDB.

| Characteristic | All eligible, n (%) |  |  |  |  | Stage IV at diagnosis, n (%) |  |  |  |  |
| --- | --- | --- | --- | --- | --- | --- | --- | --- | --- | --- |
|  | SEER | FHRD |  | CGDB |  | SEER | FHRD |  | CGDB |  |
| Initial diagnosis | 2011–18 | 2011–18 | 2011–20 | 2011–18 | 2011–20 | 2011–18 | 2011–18 | 2011–20 | 2011–18 | 2011–20 |
|  | N=25,664 | N=4,324 | N=5,321 | N=85 | N=93 | N=6,671 | N=1,690 | N=2,066 | N=44 | N=47 |
| **Age at initial diagnosis (years)** |  |  |  |  |  |  |  |  |  |  |
| 0–19 | 87 (0.3) | <5 (<0.1) | 5 (<0.1) | <5 (<5.9) | <5 (<5.4) | <5 (<0.1) | <5 (<0.3) | <5 (<0.1) | <5 (<11) | <5 (<11) |
| 20–34 | 420 (1.6) | 51 (1.2) | 60 (1.1) | <5 (<5.9) | <5 (<5.4) | 107 (1.6) | 26 (1.5) | 28 (1.4) | <5 (<11) | <5 (<11) |
| 35–49 | 2,987 (12) | 425 (9.8) | 505 (9.5) | 20 (24) | 24 (26) | 897 (13) | 169 (10) | 204 (9.9) | 12 (27) | 14 (30) |
| 50–64 | 8,999 (35) | 1,479 (34) | 1,805 (34) | 34 (40) | 36 (39) | 2,484 (37) | 574 (34) | 711 (34) | 18 (41) | 19 (40) |
| 65–79 | 9,772 (38) | 1,835 (42) | 2,270 (43) | 27 (32) | 28 (30) | 2,506 (38) | 840 (50) | 987 (48) | 14 (32) | 14 (30) |
| 80+ | 3,399 (13) | 530 (12) | 676 (13) | <5 (<5.9) | 5 (5.4) | 676 (10) | 81 (4.8) | 135 (6.5) | 0 (0) | 0 (0) |
| **Sex** |  |  |  |  |  |  |  |  |  |  |
| Female | 12,609 (49) | 2,189 (51) | 2,701 (51) | 32 (38) | 35 (38) | 3,323 (50) | 842 (50) | 1,040 (50) | 21 (48) | 22 (47) |
| Male | 13,055 (51) | 2,133 (49) | 2,618 (49) | 53 (62) | 58 (62) | 3,348 (50) | 847 (50) | 1,025 (50) | 23 (52) | 25 (53) |
| Unknown | 0 (0) | <5 (<0.1) | <5 (<0.1) | 0 (0) | 0 (0) | 0 (0) | <5 (<0.3) | <5 (<0.2) | 0 (0) | 0 (0) |
| **Race** |  |  |  |  |  |  |  |  |  |  |
| Asian | 1,413 (5.5) | 69 (1.6) | 88 (1.7) | <5 (<5.9) | <5 (<5.4) | 381 (5.7) | 25 (1.5) | 36 (1.7) | <5 (<11) | <5 (<11) |
| Black | 1,230 (4.8) | 182 (4.2) | 219 (4.1) | <5 (<5.9) | <5 (<5.4) | 355 (5.3) | 74 (4.4) | 96 (4.6) | <5 (<11) | <5 (<11) |
| Other | 118 (0.5) | 467 (11) | 608 (11) | 16 (19) | 19 (20) | 32 (0.5) | 184 (11) | 246 (12) | 7 (16) | 8 (17) |
| Unknown | 373 (1.5) | 359 (8.3) | 482 (9.1) | 8 (9.4) | 8 (8.6) | 50 (0.7) | 147 (8.7) | 190 (9.2) | <5 (<11) | <5 (<11) |
| White | 22,530 (88) | 3,247 (75) | 3,924 (74) | 58 (68) | 62 (67) | 5,853 (88) | 1,260 (75) | 1,498 (73) | 31 (70) | 33 (70) |

| AJCC stage at diagnosis |  |  |  |  |  |  |  |  |  |  |
| --- | --- | --- | --- | --- | --- | --- | --- | --- | --- | --- |
| Stage I | 6,656 (26) | 752 (17) | 924 (17) | 6 (7.1) | 7 (7.5) | NA | NA | NA | NA | NA |
| Stage II | 3,835 (15) | 570 (13) | 712 (13) | 5 (5.9) | 6 (6.5) | NA | NA | NA | NA | NA |
| Stage III | 6,480 (25) | 1,281 (30) | 1,586 (30) | 29 (34) | 32 (34) | NA | NA | NA | NA | NA |
| Stage IV | 6,671 (26) | 1,690 (39) | 2,066 (39) | 44 (52) | 47 (51) | NA | NA | NA | NA | NA |
| Unknown | 2,022 (7.9) | 31 (0.7) | 33 (0.6) | <5 (<5.9) | <5 (<5.4) | NA | NA | NA | NA | NA |
| Year of initial diagnosis |  |  |  |  |  |  |  |  |  |  |
| 2011 | 3,104 (12) | 488 (11) | 488 (9.2) | 5 (5.9) | 5 (5.4) | 815 (12) | 189 (11) | 189 (9.1) | <5 (<11) | <5 (<11) |
| 2012 | 3,093 (12) | 516 (12) | 516 (9.7) | 6 (7.1) | 6 (6.5) | 788 (12) | 228 (13) | 228 (11) | <5 (<11) | <5 (<11) |
| 2013 | 3,079 (12) | 478 (11) | 478 (9.0) | 16 (19) | 16 (17) | 819 (12) | 192 (11) | 192 (9.3) | 7 (16) | 7 (15) |
| 2014 | 3,303 (13) | 598 (14) | 598 (11) | 9 (11) | 9 (9.7) | 851 (13) | 221 (13) | 221 (11) | <5 (<11) | <5 (<11) |
| 2015 | 3,227 (13) | 582 (13) | 582 (11) | 11 (13) | 11 (12) | 836 (13) | 219 (13) | 219 (11) | 5 (11) | 5 (11) |
| 2016 | 3,351 (13) | 588 (14) | 588 (11) | 10 (12) | 10 (11) | 885 (13) | 234 (14) | 234 (11) | 5 (11) | 5 (11) |
| 2017 | 3,209 (13) | 556 (13) | 556 (10) | 15 (18) | 15 (16) | 815 (12) | 233 (14) | 233 (11) | 9 (20) | 9 (19) |
| 2018 | 3,298 (13) | 518 (12) | 518 (9.7) | 13 (15) | 13 (14) | 862 (13) | 174 (10) | 174 (8.4) | 8 (18) | 8 (17) |
| 2019 | NA | NA | 551 (10) | NA | 5 (5.4) | NA | NA | 211 (10) | NA | <5 (<11) |
| 2020 | NA | NA | 414 (7.8) | NA | <5 (<5.4) | NA | NA | 153 (7.4) | NA | <5 (<11) |
| 2021 | NA | NA | 32 (0.6) | NA | <5 (<5.4) | NA | NA | 12 (0.6) | NA | <5 (<11) |
| Abbreviations: AJCC, American Joint Committee on Cancer; CGDB, Flatiron Health-Foundation Medicine Clinico-Genomic Database; FHRD, Flatiron Health Research Database; NA, not applicable; SEER, The Surveillance, Epidemiology, and End Results research database. |  |  |  |  |  |  |  |  |  |  |
| Note: Reported percentages may not equal exactly to 100% due to rounding. |  |  |  |  |  |  |  |  |  |  |

**Table 12.**
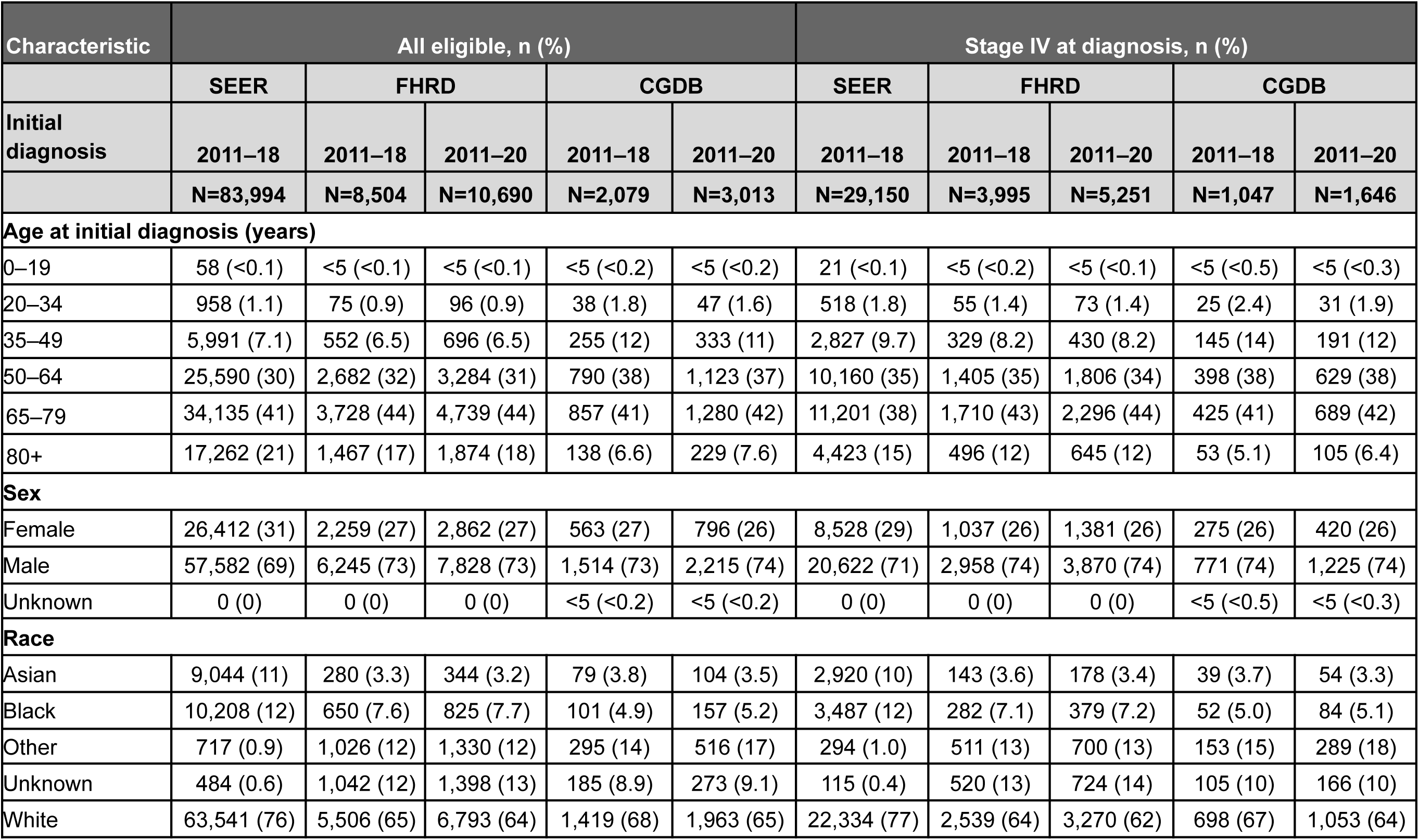
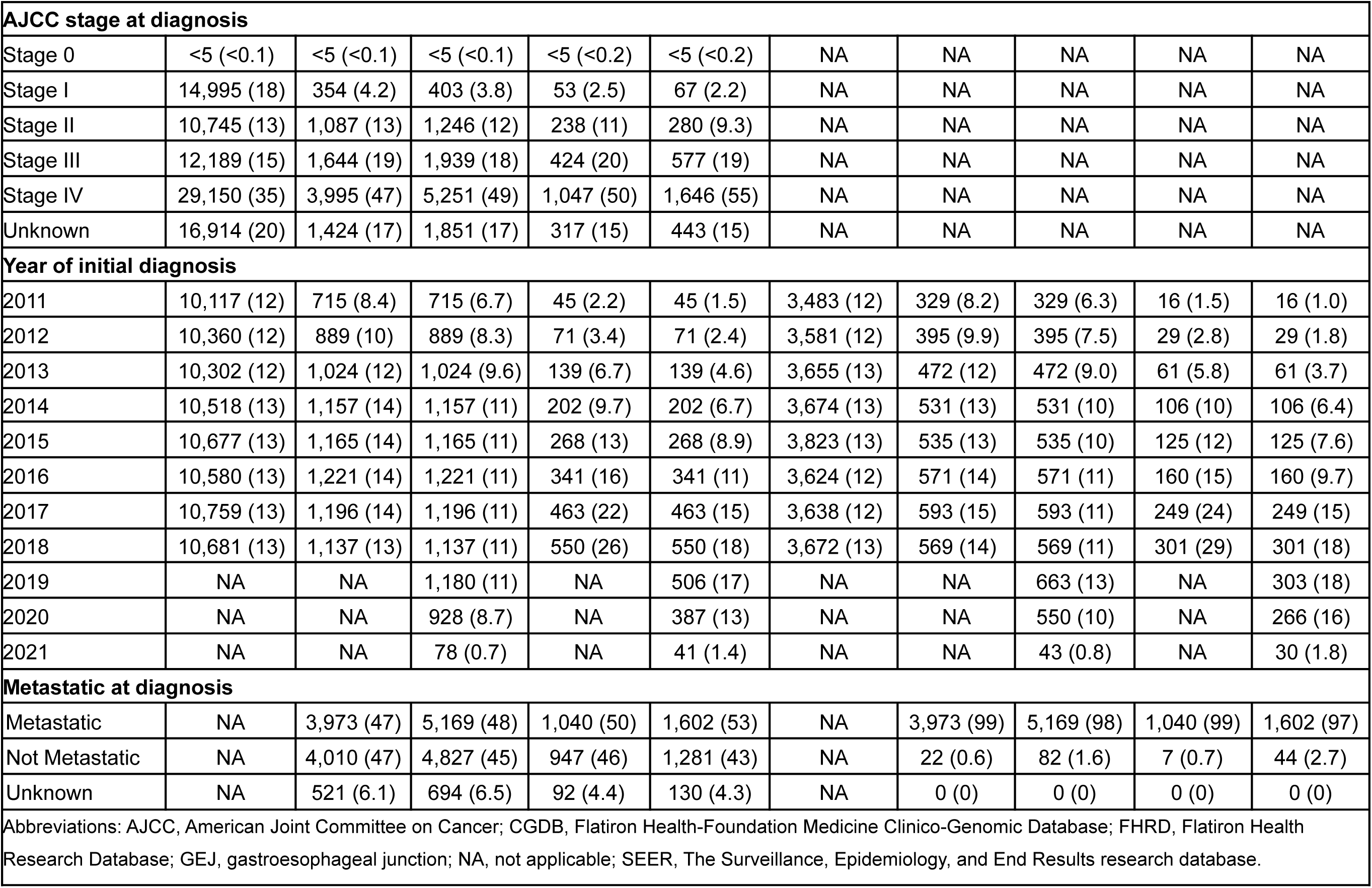
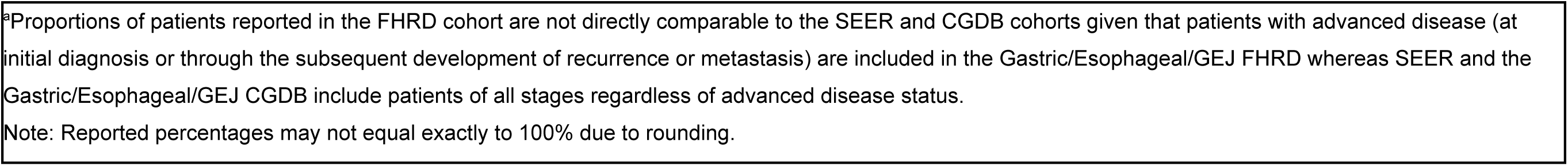
Gastric/Esophageal Cancer/GEJ – Comparison of Demographic and Clinical Characteristics Between SEER vs FHRD (advanced at any time) vs CGDB^a^.

| Characteristic | All eligible, n (%) |  |  |  |  | Stage IV at diagnosis, n (%) |  |  |  |  |
| --- | --- | --- | --- | --- | --- | --- | --- | --- | --- | --- |
|  | SEER | FHRD |  | CGDB |  | SEER | FHRD |  | CGDB |  |
| Initial diagnosis | 2011–18 | 2011–18 | 2011–20 | 2011–18 | 2011–20 | 2011–18 | 2011–18 | 2011–20 | 2011–18 | 2011–20 |
|  | N=83,994 | N=8,504 | N=10,690 | N=2,079 | N=3,013 | N=29,150 | N=3,995 | N=5,251 | N=1,047 | N=1,646 |
| **Age at initial diagnosis (years)** |  |  |  |  |  |  |  |  |  |  |
| 0–19 | 58 (<0.1) | <5 (<0.1) | <5 (<0.1) | <5 (<0.2) | <5 (<0.2) | 21 (<0.1) | <5 (<0.2) | <5 (<0.1) | <5 (<0.5) | <5 (<0.3) |
| 20–34 | 958 (1.1) | 75 (0.9) | 96 (0.9) | 38 (1.8) | 47 (1.6) | 518 (1.8) | 55 (1.4) | 73 (1.4) | 25 (2.4) | 31 (1.9) |
| 35–49 | 5,991 (7.1) | 552 (6.5) | 696 (6.5) | 255 (12) | 333 (11) | 2,827 (9.7) | 329 (8.2) | 430 (8.2) | 145 (14) | 191 (12) |
| 50–64 | 25,590 (30) | 2,682 (32) | 3,284 (31) | 790 (38) | 1,123 (37) | 10,160 (35) | 1,405 (35) | 1,806 (34) | 398 (38) | 629 (38) |
| 65–79 | 34,135 (41) | 3,728 (44) | 4,739 (44) | 857 (41) | 1,280 (42) | 11,201 (38) | 1,710 (43) | 2,296 (44) | 425 (41) | 689 (42) |
| 80+ | 17,262 (21) | 1,467 (17) | 1,874 (18) | 138 (6.6) | 229 (7.6) | 4,423 (15) | 496 (12) | 645 (12) | 53 (5.1) | 105 (6.4) |
| **Sex** |  |  |  |  |  |  |  |  |  |  |
| Female | 26,412 (31) | 2,259 (27) | 2,862 (27) | 563 (27) | 796 (26) | 8,528 (29) | 1,037 (26) | 1,381 (26) | 275 (26) | 420 (26) |
| Male | 57,582 (69) | 6,245 (73) | 7,828 (73) | 1,514 (73) | 2,215 (74) | 20,622 (71) | 2,958 (74) | 3,870 (74) | 771 (74) | 1,225 (74) |
| Unknown | 0 (0) | 0 (0) | 0 (0) | <5 (<0.2) | <5 (<0.2) | 0 (0) | 0 (0) | 0 (0) | <5 (<0.5) | <5 (<0.3) |
| **Race** |  |  |  |  |  |  |  |  |  |  |
| Asian | 9,044 (11) | 280 (3.3) | 344 (3.2) | 79 (3.8) | 104 (3.5) | 2,920 (10) | 143 (3.6) | 178 (3.4) | 39 (3.7) | 54 (3.3) |
| Black | 10,208 (12) | 650 (7.6) | 825 (7.7) | 101 (4.9) | 157 (5.2) | 3,487 (12) | 282 (7.1) | 379 (7.2) | 52 (5.0) | 84 (5.1) |
| Other | 717 (0.9) | 1,026 (12) | 1,330 (12) | 295 (14) | 516 (17) | 294 (1.0) | 511 (13) | 700 (13) | 153 (15) | 289 (18) |
| Unknown | 484 (0.6) | 1,042 (12) | 1,398 (13) | 185 (8.9) | 273 (9.1) | 115 (0.4) | 520 (13) | 724 (14) | 105 (10) | 166 (10) |
| White | 63,541 (76) | 5,506 (65) | 6,793 (64) | 1,419 (68) | 1,963 (65) | 22,334 (77) | 2,539 (64) | 3,270 (62) | 698 (67) | 1,053 (64) |

| AJCC stage at diagnosis |  |  |  |  |  |  |  |  |  |  |
| --- | --- | --- | --- | --- | --- | --- | --- | --- | --- | --- |
| Stage 0 | <5 (<0.1) | <5 (<0.1) | <5 (<0.1) | <5 (<0.2) | <5 (<0.2) | NA | NA | NA | NA | NA |
| Stage I | 14,995 (18) | 354 (4.2) | 403 (3.8) | 53 (2.5) | 67 (2.2) | NA | NA | NA | NA | NA |
| Stage II | 10,745 (13) | 1,087 (13) | 1,246 (12) | 238 (11) | 280 (9.3) | NA | NA | NA | NA | NA |
| Stage III | 12,189 (15) | 1,644 (19) | 1,939 (18) | 424 (20) | 577 (19) | NA | NA | NA | NA | NA |
| Stage IV | 29,150 (35) | 3,995 (47) | 5,251 (49) | 1,047 (50) | 1,646 (55) | NA | NA | NA | NA | NA |
| Unknown | 16,914 (20) | 1,424 (17) | 1,851 (17) | 317 (15) | 443 (15) | NA | NA | NA | NA | NA |
| Year of initial diagnosis |  |  |  |  |  |  |  |  |  |  |
| 2011 | 10,117 (12) | 715 (8.4) | 715 (6.7) | 45 (2.2) | 45 (1.5) | 3,483 (12) | 329 (8.2) | 329 (6.3) | 16 (1.5) | 16 (1.0) |
| 2012 | 10,360 (12) | 889 (10) | 889 (8.3) | 71 (3.4) | 71 (2.4) | 3,581 (12) | 395 (9.9) | 395 (7.5) | 29 (2.8) | 29 (1.8) |
| 2013 | 10,302 (12) | 1,024 (12) | 1,024 (9.6) | 139 (6.7) | 139 (4.6) | 3,655 (13) | 472 (12) | 472 (9.0) | 61 (5.8) | 61 (3.7) |
| 2014 | 10,518 (13) | 1,157 (14) | 1,157 (11) | 202 (9.7) | 202 (6.7) | 3,674 (13) | 531 (13) | 531 (10) | 106 (10) | 106 (6.4) |
| 2015 | 10,677 (13) | 1,165 (14) | 1,165 (11) | 268 (13) | 268 (8.9) | 3,823 (13) | 535 (13) | 535 (10) | 125 (12) | 125 (7.6) |
| 2016 | 10,580 (13) | 1,221 (14) | 1,221 (11) | 341 (16) | 341 (11) | 3,624 (12) | 571 (14) | 571 (11) | 160 (15) | 160 (9.7) |
| 2017 | 10,759 (13) | 1,196 (14) | 1,196 (11) | 463 (22) | 463 (15) | 3,638 (12) | 593 (15) | 593 (11) | 249 (24) | 249 (15) |
| 2018 | 10,681 (13) | 1,137 (13) | 1,137 (11) | 550 (26) | 550 (18) | 3,672 (13) | 569 (14) | 569 (11) | 301 (29) | 301 (18) |
| 2019 | NA | NA | 1,180 (11) | NA | 506 (17) | NA | NA | 663 (13) | NA | 303 (18) |
| 2020 | NA | NA | 928 (8.7) | NA | 387 (13) | NA | NA | 550 (10) | NA | 266 (16) |
| 2021 | NA | NA | 78 (0.7) | NA | 41 (1.4) | NA | NA | 43 (0.8) | NA | 30 (1.8) |
| Metastatic at diagnosis |  |  |  |  |  |  |  |  |  |  |
| Metastatic | NA | 3,973 (47) | 5,169 (48) | 1,040 (50) | 1,602 (53) | NA | 3,973 (99) | 5,169 (98) | 1,040 (99) | 1,602 (97) |
| Not Metastatic | NA | 4,010 (47) | 4,827 (45) | 947 (46) | 1,281 (43) | NA | 22 (0.6) | 82 (1.6) | 7 (0.7) | 44 (2.7) |
| Unknown | NA | 521 (6.1) | 694 (6.5) | 92 (4.4) | 130 (4.3) | NA | 0 (0) | 0 (0) | 0 (0) | 0 (0) |
| Abbreviations: AJCC, American Joint Committee on Cancer; CGDB, Flatiron Health-Foundation Medicine Clinico-Genomic Database; FHRD, Flatiron Health Research Database; GEJ, gastroesophageal junction; NA, not applicable; SEER, The Surveillance, Epidemiology, and End Results research database. |  |  |  |  |  |  |  |  |  |  |

<sup>a</sup>Proportions of patients reported in the FHRD cohort are not directly comparable to the SEER and CGDB cohorts given that patients with advanced disease (at initial diagnosis or through the subsequent development of recurrence or metastasis) are included in the Gastric/Esophageal/GEJ FHRD whereas SEER and the Gastric/Esophageal/GEJ CGDB include patients of all stages regardless of advanced disease status.
Note: Reported percentages may not equal exactly to 100% due to rounding.

**Table 13.**
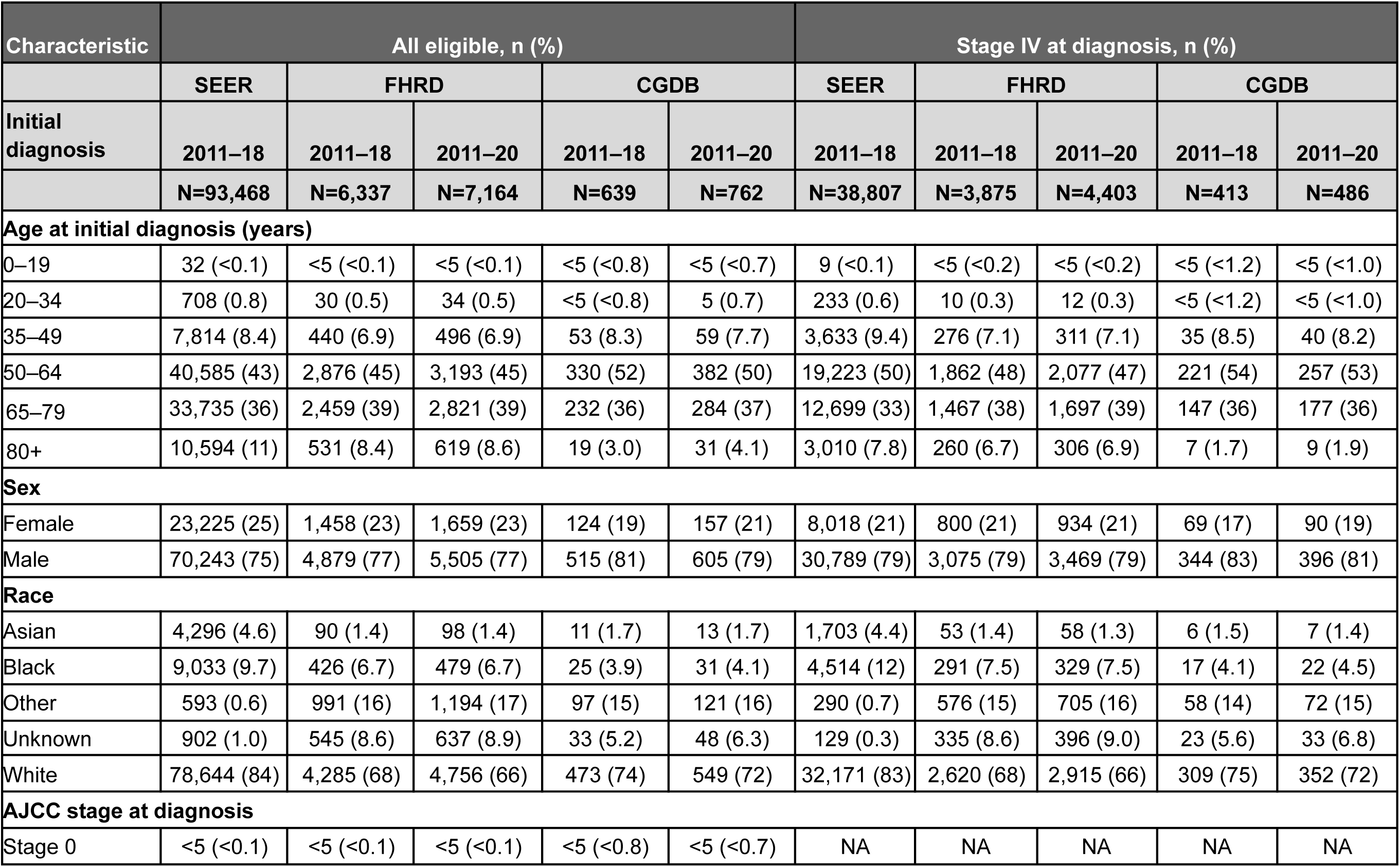
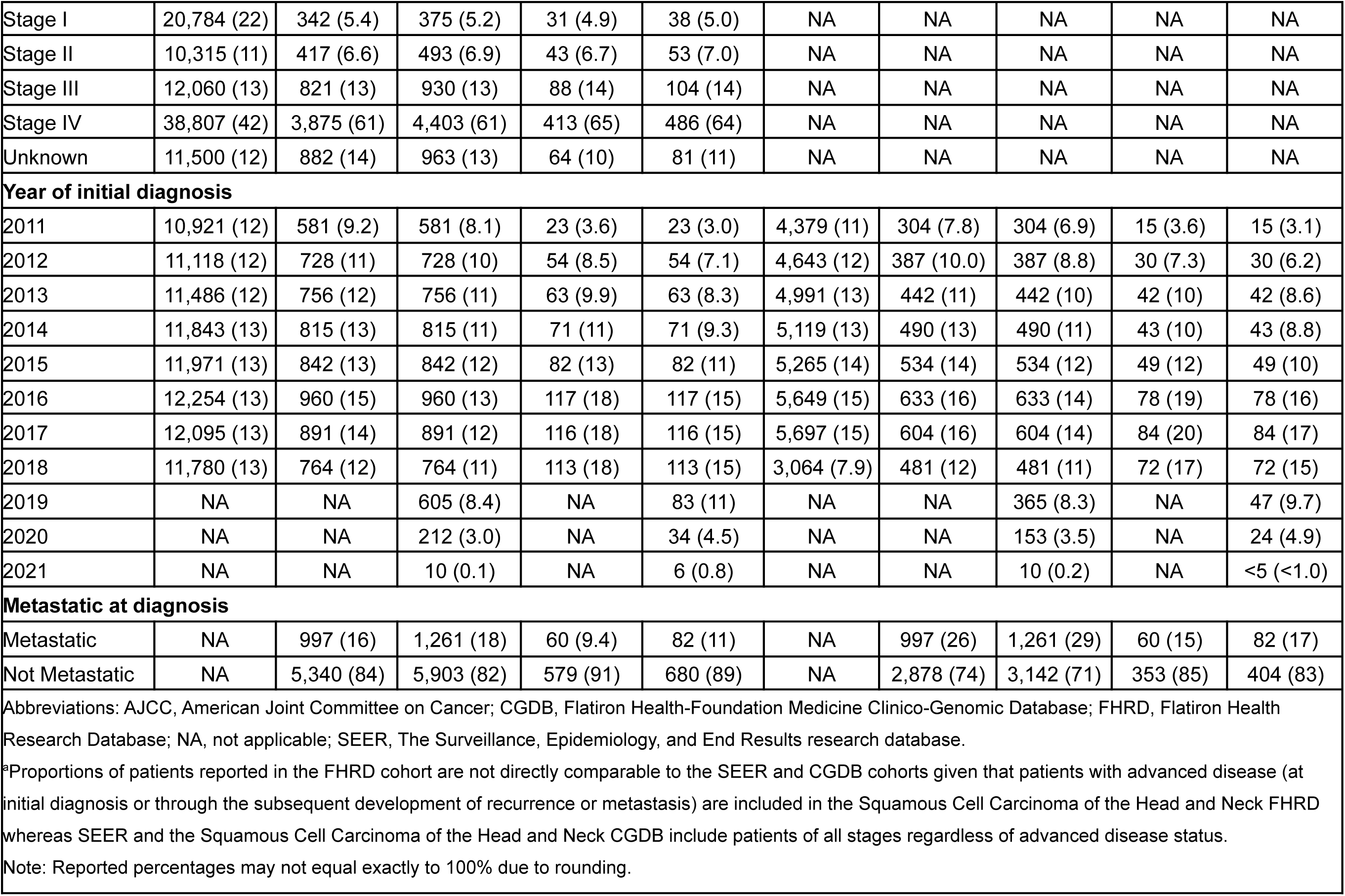
Squamous Cell Carcinoma of the Head & Neck – Comparison of Demographic and Clinical Characteristics Between SEER vs FHRD (advanced at any time) vs CGDB^a^.

**Table 14.**
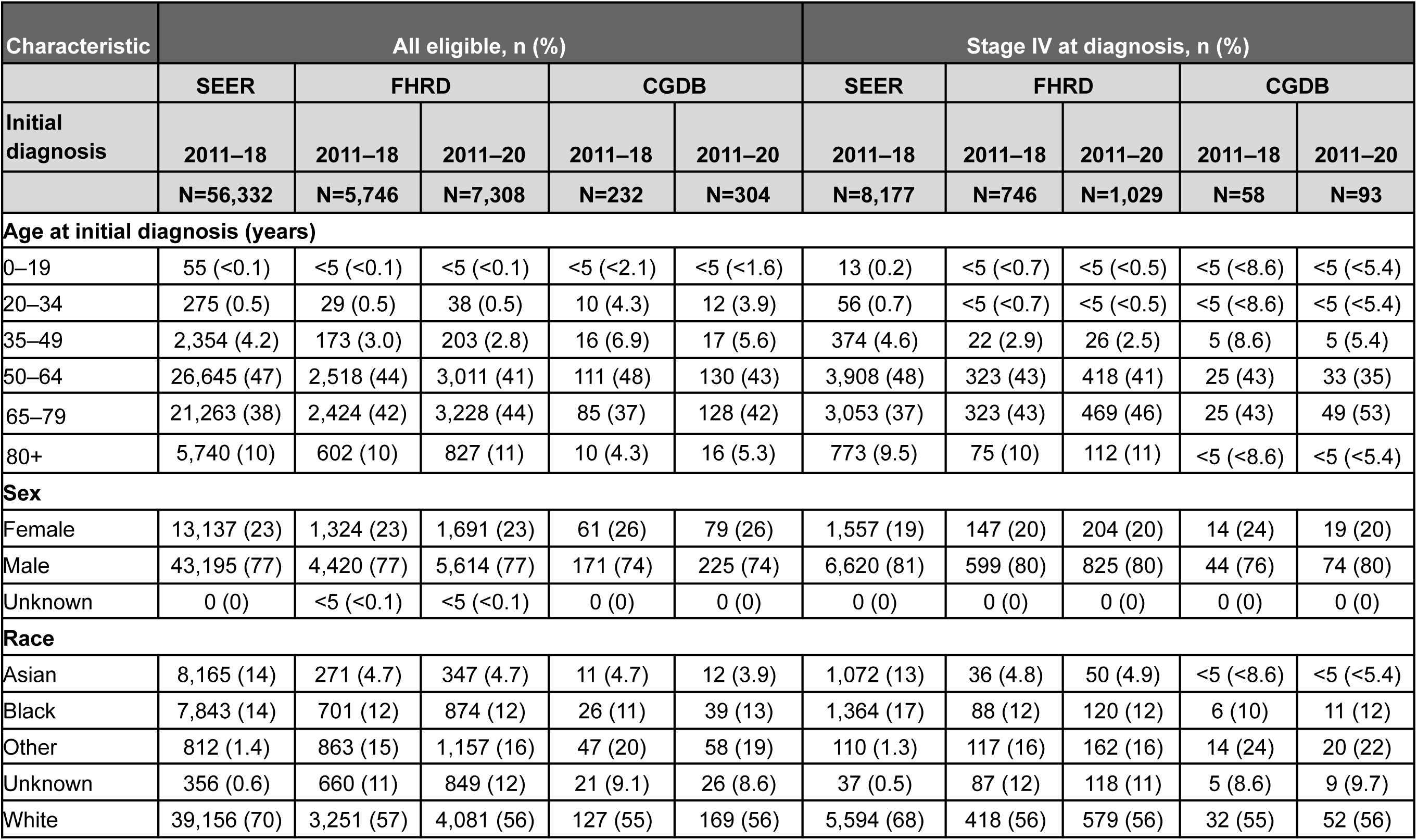
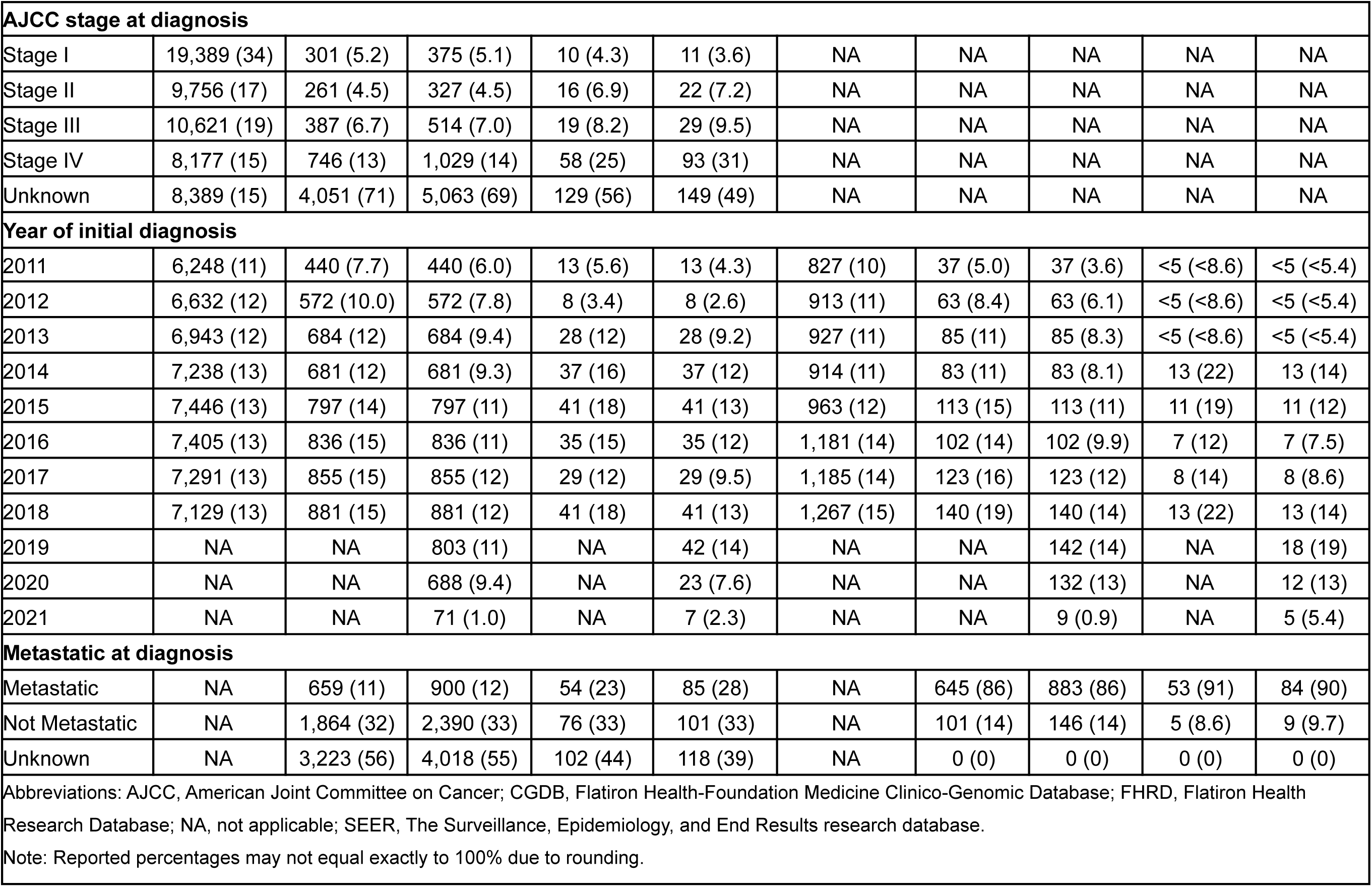
Hepatocellular Carcinoma – Comparison of Demographic and Clinical Characteristics Between SEER vs FHRD vs CGDB.

**Table 15.**
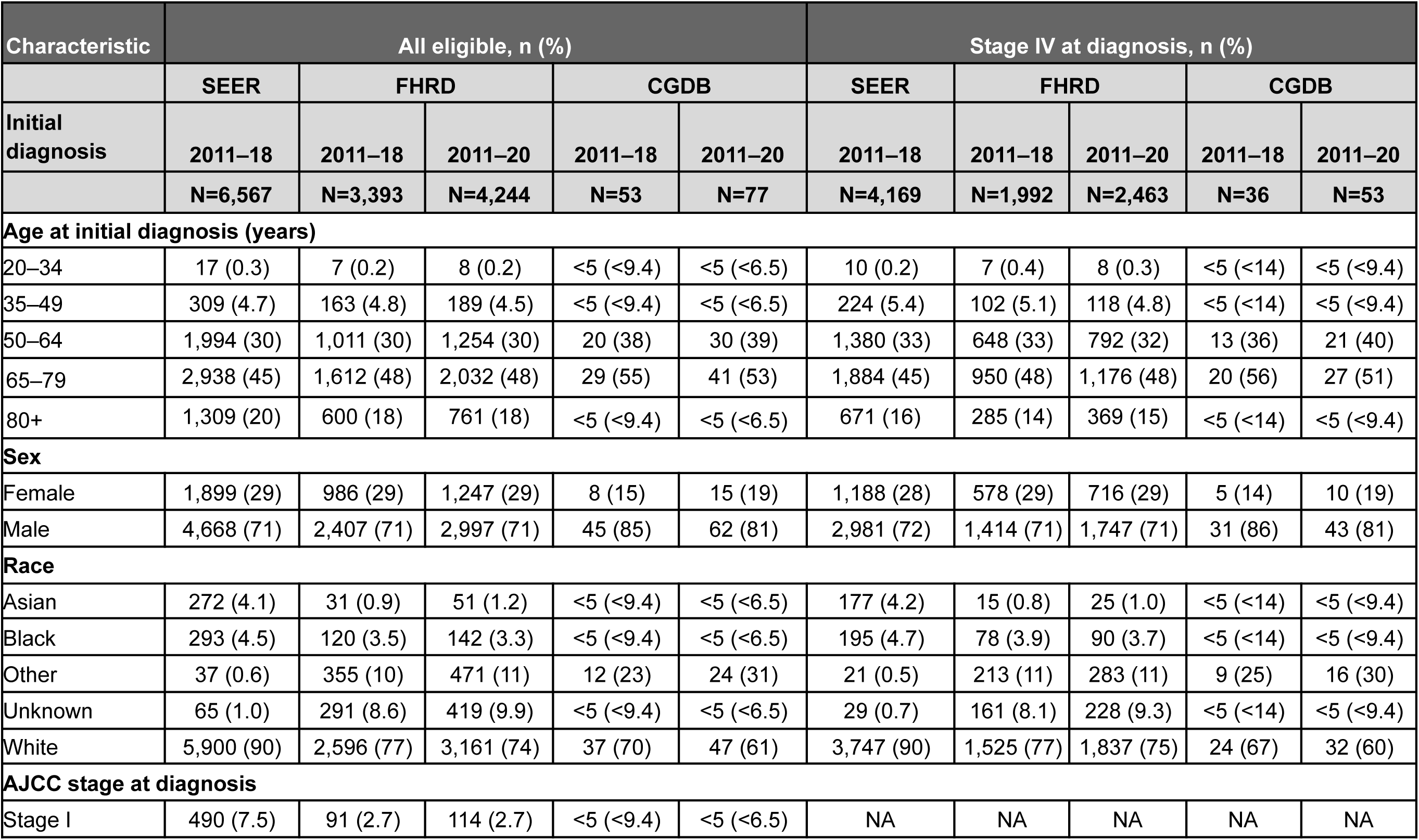
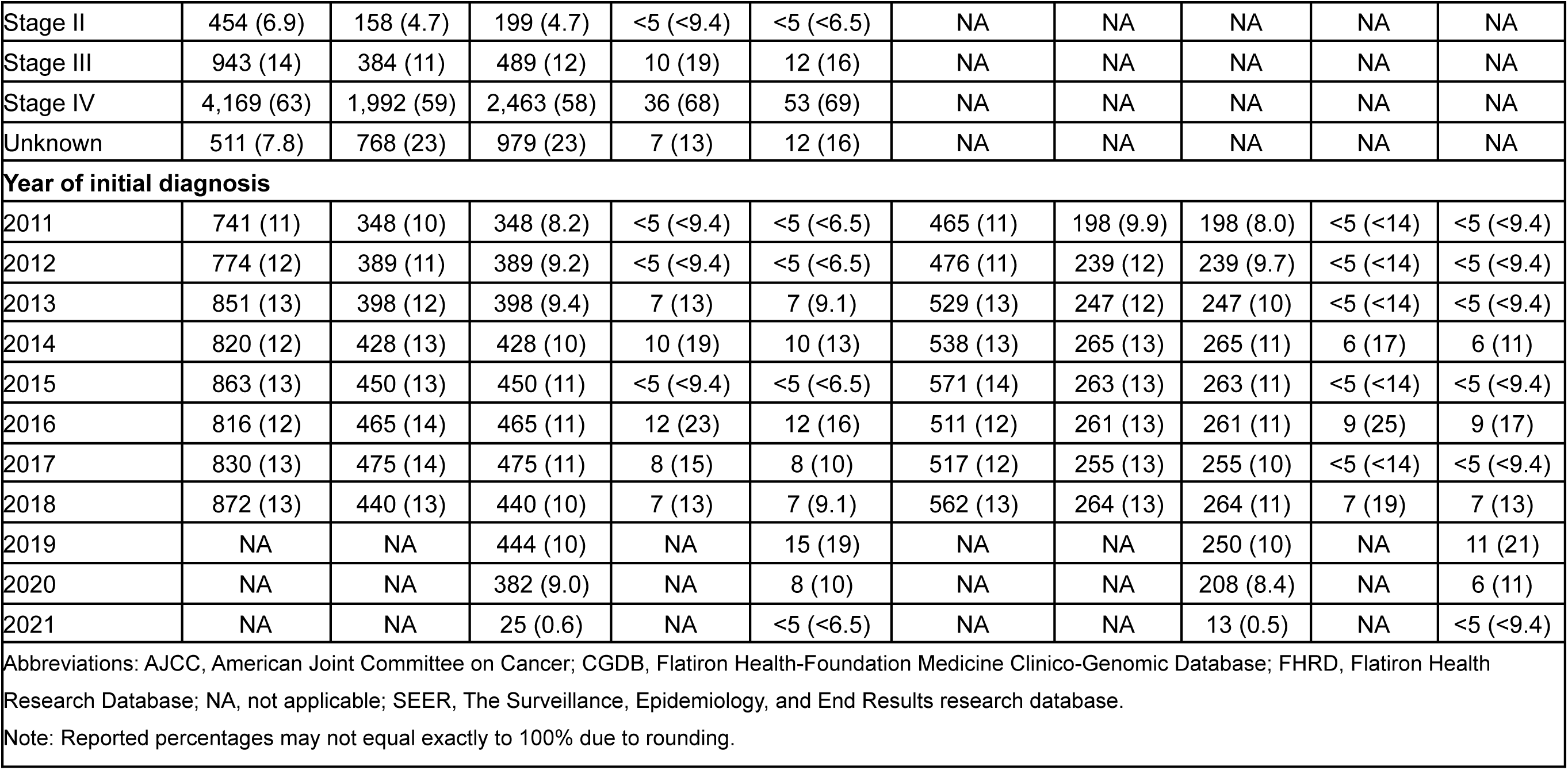
Mantle Cell Lymphoma – Comparison of Demographic and Clinical Characteristics Between SEER vs FHRD vs CGDB.

**Table 16.** Melanoma – Comparison of Demographic and Clinical Characteristics Between SEER vs FHRD (advanced-only) vs CGDB^a^.

| Characteristic | All eligible, n (%) |  |  |  |  | Stage IV at diagnosis, n (%) |  |  |  |  |
| --- | --- | --- | --- | --- | --- | --- | --- | --- | --- | --- |
|  | SEER | FHRD |  | CGDB |  | SEER | FHRD |  | CGDB |  |
| Initial diagnosis | 2011–18 | 2011–18 | 2011–20 | 2011–18 | 2011–20 | 2011–18 | 2011–18 | 2011–20 | 2011–18 | 2011–20 |
|  | N=179,698 | N=7,911 | N=9,427 | N=991 | N=1,306 | N=6,834 | N=1,724 | N=2,290 | N=218 | N=361 |
| **Age at initial diagnosis (years)** |  |  |  |  |  |  |  |  |  |  |
| 0–19 | 713 (0.4) | 24 (0.3) | 27 (0.3) | <5 (<0.5) | <5 (<0.4) | 18 (0.3) | <5 (<0.3) | <5 (<0.2) | <5 (<2.3) | <5 (<1.4) |
| 20–34 | 9,255 (5.2) | 378 (4.8) | 432 (4.6) | 34 (3.4) | 49 (3.8) | 185 (2.7) | 47 (2.7) | 61 (2.7) | 11 (5.0) | 17 (4.7) |
| 35–49 | 23,805 (13) | 938 (12) | 1,099 (12) | 120 (12) | 157 (12) | 660 (9.7) | 162 (9.4) | 219 (9.6) | 22 (10) | 40 (11) |
| 50–64 | 55,516 (31) | 2,410 (30) | 2,806 (30) | 346 (35) | 432 (33) | 2,154 (32) | 532 (31) | 676 (30) | 67 (31) | 103 (29) |
| 65–79 | 61,407 (34) | 2,964 (37) | 3,565 (38) | 382 (39) | 513 (39) | 2,538 (37) | 671 (39) | 901 (39) | 89 (41) | 154 (43) |
| 80+ | 29,002 (16) | 1,197 (15) | 1,498 (16) | 107 (11) | 152 (12) | 1,279 (19) | 312 (18) | 433 (19) | 29 (13) | 47 (13) |
| **Sex** |  |  |  |  |  |  |  |  |  |  |
| Female | 73,251 (41) | 2,693 (34) | 3,204 (34) | 330 (33) | 435 (33) | 2,087 (31) | 535 (31) | 703 (31) | 70 (32) | 116 (32) |
| Male | 106,447 (59) | 5,217 (66) | 6,222 (66) | 661 (67) | 871 (67) | 4,747 (69) | 1,189 (69) | 1,587 (69) | 148 (68) | 245 (68) |
| Unknown | 0 (0) | <5 (<0.1) | <5 (<0.1) | 0 (0) | 0 (0) | 0 (0) | 0 (0) | 0 (0) | 0 (0) | 0 (0) |
| **Race** |  |  |  |  |  |  |  |  |  |  |
| Asian | 1,098 (0.6) | 18 (0.2) | 21 (0.2) | <5 (<0.5) | <5 (<0.4) | 100 (1.5) | 6 (0.3) | 7 (0.3) | <5 (<2.3) | <5 (<1.4) |
| Black | 754 (0.4) | 33 (0.4) | 44 (0.5) | <5 (<0.5) | <5 (<0.4) | 95 (1.4) | 16 (0.9) | 22 (1.0) | <5 (<2.3) | <5 (<1.4) |
| Other | 445 (0.2) | 514 (6.5) | 659 (7.0) | 106 (11) | 155 (12) | 27 (0.4) | 141 (8.2) | 211 (9.2) | 29 (13) | 53 (15) |
| Unknown | 8,606 (4.8) | 631 (8.0) | 851 (9.0) | 55 (5.5) | 100 (7.7) | 27 (0.4) | 166 (9.6) | 257 (11) | 11 (5.0) | 29 (8.0) |
| White | 168,795 (94) | 6,715 (85) | 7,852 (83) | 827 (83) | 1,047 (80) | 6,585 (96) | 1,395 (81) | 1,793 (78) | 177 (81) | 277 (77) |

| AJCC stage at diagnosis |  |  |  |  |  |  |  |  |  |  |
| --- | --- | --- | --- | --- | --- | --- | --- | --- | --- | --- |
| Stage 0 | 79 (<0.1) | 21 (0.3) | 22 (0.2) | <5 (<0.5) | <5 (<0.4) | NA | NA | NA | NA | NA |
| Stage I | 116,777 (65) | 604 (7.6) | 624 (6.6) | 104 (10) | 113 (8.7) | NA | NA | NA | NA | NA |
| Stage II | 20,654 (11) | 1,187 (15) | 1,271 (13) | 202 (20) | 241 (18) | NA | NA | NA | NA | NA |
| Stage III | 11,578 (6.4) | 3,458 (44) | 4,271 (45) | 312 (31) | 422 (32) | NA | NA | NA | NA | NA |
| Stage IV | 6,834 (3.8) | 1,724 (22) | 2,290 (24) | 218 (22) | 361 (28) | NA | NA | NA | NA | NA |
| Unknown | 23,776 (13) | 917 (12) | 949 (10) | 153 (15) | 167 (13) | NA | NA | NA | NA | NA |
| Year of initial diagnosis |  |  |  |  |  |  |  |  |  |  |
| 2011 | 19,368 (11) | 788 (10.0) | 788 (8.4) | 52 (5.2) | 52 (4.0) | 722 (11) | 104 (6.0) | 104 (4.5) | <5 (<2.3) | <5 (<1.4) |
| 2012 | 20,517 (11) | 894 (11) | 894 (9.5) | 81 (8.2) | 81 (6.2) | 746 (11) | 153 (8.9) | 153 (6.7) | 9 (4.1) | 9 (2.5) |
| 2013 | 21,277 (12) | 978 (12) | 978 (10) | 94 (9.5) | 94 (7.2) | 806 (12) | 195 (11) | 195 (8.5) | 10 (4.6) | 10 (2.8) |
| 2014 | 23,024 (13) | 1,061 (13) | 1,061 (11) | 105 (11) | 105 (8.0) | 914 (13) | 219 (13) | 219 (9.6) | 14 (6.4) | 14 (3.9) |
| 2015 | 23,673 (13) | 1,089 (14) | 1,089 (12) | 136 (14) | 136 (10) | 867 (13) | 226 (13) | 226 (9.9) | 31 (14) | 31 (8.6) |
| 2016 | 23,574 (13) | 1,068 (14) | 1,068 (11) | 135 (14) | 135 (10) | 902 (13) | 263 (15) | 263 (11) | 34 (16) | 34 (9.4) |
| 2017 | 24,097 (13) | 1,047 (13) | 1,047 (11) | 176 (18) | 176 (13) | 918 (13) | 278 (16) | 278 (12) | 53 (24) | 53 (15) |
| 2018 | 24,168 (13) | 986 (12) | 986 (10) | 212 (21) | 212 (16) | 959 (14) | 286 (17) | 286 (12) | 64 (29) | 64 (18) |
| 2019 | NA | NA | 888 (9.4) | NA | 177 (14) | NA | NA | 297 (13) | NA | 70 (19) |
| 2020 | NA | NA | 594 (6.3) | NA | 120 (9.2) | NA | NA | 248 (11) | NA | 61 (17) |
| 2021 | NA | NA | 34 (0.4) | NA | 18 (1.4) | NA | NA | 21 (0.9) | NA | 12 (3.3) |
| Abbreviations: AJCC, American Joint Committee on Cancer; CGDB, Flatiron Health-Foundation Medicine Clinico-Genomic Database; FHRD, Flatiron Health Research Database; NA, not applicable; SEER, The Surveillance, Epidemiology, and End Results research database. |  |  |  |  |  |  |  |  |  |  |
| *Proportions of patients reported in the FHRD cohort are not directly comparable to the SEER and CGDB cohorts given that patients with advanced disease (at initial diagnosis or through the subsequent development of recurrence or metastasis) are included in the Melanoma FHRD whereas SEER and the Melanoma CGDB include patients of all stages regardless of advanced disease status. |  |  |  |  |  |  |  |  |  |  |
| Note: Reported percentages may not equal exactly to 100% due to rounding. |  |  |  |  |  |  |  |  |  |  |

**Table 17.** Multiple Myeloma – Comparison of Demographic and Clinical Characteristics Between SEER vs FHRD vs CGDB.

| Characteristic | All eligible, n (%) |  |  |  |  |
| --- | --- | --- | --- | --- | --- |
|  | SEER | FHRD |  | CGDB |  |
| Initial diagnosis | 2011–18 | 2011–18 | 2011–20 | 2011–18 | 2011–20 |
|  | N=50,504 | N=9,676 | N=12,093 | N=267 | N=292 |
| **Age at initial diagnosis (years)** |  |  |  |  |  |
| 0–19 | <5 (<0.1) | <5 (<0.1) | <5 (<0.1) | <5 (<1.9) | <5 (<1.7) |
| 20–34 | 202 (0.4) | 34 (0.4) | 41 (0.3) | <5 (<1.9) | <5 (<1.7) |
| 35–49 | 3,174 (6.3) | 525 (5.4) | 639 (5.3) | 28 (10) | 30 (10) |
| 50–64 | 14,882 (29) | 2,926 (30) | 3,561 (29) | 146 (55) | 161 (55) |
| 65–79 | 22,272 (44) | 4,463 (46) | 5,686 (47) | 84 (31) | 91 (31) |
| 80+ | 9,970 (20) | 1,728 (18) | 2,166 (18) | 7 (2.6) | 8 (2.7) |
| **Sex** |  |  |  |  |  |
| Female | 22,249 (44) | 4,458 (46) | 5,545 (46) | 111 (42) | 125 (43) |
| Male | 28,255 (56) | 5,218 (54) | 6,548 (54) | 156 (58) | 167 (57) |
| **Race** |  |  |  |  |  |
| Asian | 2,871 (5.7) | 173 (1.8) | 217 (1.8) | <5 (<1.9) | <5 (<1.7) |
| Black | 10,442 (21) | 1,608 (17) | 2,019 (17) | 24 (9.0) | 29 (9.9) |
| Other | 348 (0.7) | 1,038 (11) | 1,331 (11) | 28 (10) | 32 (11) |
| Unknown | 488 (1.0) | 952 (9.8) | 1,309 (11) | 22 (8.2) | 24 (8.2) |
| White | 36,355 (72) | 5,905 (61) | 7,217 (60) | 189 (71) | 203 (70) |
| **ISS stage at initial diagnosis** |  |  |  |  |  |
| Unknown | NA | 4,909 (51) | 5,836 (48) | 139 (52) | 150 (51) |
| Stage I | NA | 1,634 (17) | 2,135 (18) | 56 (21) | 63 (22) |
| Stage II | NA | 1,564 (16) | 2,057 (17) | 28 (10) | 34 (12) |
| Stage III | NA | 1,569 (16) | 2,065 (17) | 44 (16) | 45 (15) |
| **Year of initial diagnosis** |  |  |  |  |  |
| 2011 | 5,671 (11) | 900 (9.3) | 900 (7.4) | 20 (7.5) | 20 (6.8) |
| 2012 | 5,932 (12) | 1,051 (11) | 1,051 (8.7) | 35 (13) | 35 (12) |
| 2013 | 6,076 (12) | 1,121 (12) | 1,121 (9.3) | 40 (15) | 40 (14) |
| 2014 | 6,191 (12) | 1,208 (12) | 1,208 (10.0) | 44 (16) | 44 (15) |
| 2015 | 6,531 (13) | 1,344 (14) | 1,344 (11) | 46 (17) | 46 (16) |
| 2016 | 6,793 (13) | 1,417 (15) | 1,417 (12) | 45 (17) | 45 (15) |
| 2017 | 6,739 (13) | 1,292 (13) | 1,292 (11) | 19 (7.1) | 19 (6.5) |
| 2018 | 6,571 (13) | 1,343 (14) | 1,343 (11) | 18 (6.7) | 18 (6.2) |
| 2019 | NA | NA | 1,309 (11) | NA | 14 (4.8) |
| 2020 | NA | NA | 995 (8.2) | NA | 11 (3.8) |
| 2021 | NA | NA | 113 (0.9) | NA | <5 (<1.7) |
| Abbreviations: CGDB, Flatiron Health-Foundation Medicine Clinico-Genomic Database; FHRD, Flatiron Health Research Database; ISS, International Staging System for Multiple Myeloma; NA, not applicable; SEER, The Surveillance, Epidemiology, and End Results research database. |  |  |  |  |  |
| Note: Reported percentages may not equal exactly to 100% due to rounding. |  |  |  |  |  |

**Table 18.** Non–Small Cell Lung Cancer – Comparison of Demographic and Clinical Characteristics Between SEER vs FHRD (advanced-only) vs CGDB^a^.

| Characteristic | All eligible, n (%) |  |  |  |  | Stage IV at diagnosis, n (%) |  |  |  |  |
| --- | --- | --- | --- | --- | --- | --- | --- | --- | --- | --- |
|  | SEER | FHRD |  | CGDB |  | SEER | FHRD |  | CGDB |  |
| Initial diagnosis | 2011–18 | 2011–18 | 2011–20 | 2011–18 | 2011–20 | 2011–18 | 2011–18 | 2011–20 | 2011–18 | 2011–20 |
|  | N=292,179 | N=54,222 | N=65,348 | N=9,517 | N=13,604 | N=122,539 | N=33,828 | N=42,008 | N=5,006 | N=7,581 |
| **Age at initial diagnosis (years)** |  |  |  |  |  |  |  |  |  |  |
| 0–19 | 18 (<0.1) | <5 (<0.1) | <5 (<0.1) | <5 (<0.1) | <5 (<0.1) | 7 (<0.1) | <5 (<0.1) | <5 (<0.1) | <5 (<0.1) | <5 (<0.1) |
| 20–34 | 475 (0.2) | 106 (0.2) | 127 (0.2) | 42 (0.4) | 49 (0.4) | 322 (0.3) | 83 (0.2) | 103 (0.2) | 30 (0.6) | 37 (0.5) |
| 35–49 | 8,490 (2.9) | 1,897 (3.5) | 2,233 (3.4) | 480 (5.0) | 607 (4.5) | 4,905 (4.0) | 1,339 (4.0) | 1,615 (3.8) | 318 (6.4) | 412 (5.4) |
| 50–64 | 79,831 (27) | 16,536 (30) | 19,669 (30) | 3,204 (34) | 4,332 (32) | 37,708 (31) | 10,716 (32) | 13,025 (31) | 1,805 (36) | 2,549 (34) |
| 65–79 | 148,440 (51) | 27,509 (51) | 33,256 (51) | 4,724 (50) | 6,863 (50) | 57,914 (47) | 16,419 (49) | 20,591 (49) | 2,297 (46) | 3,614 (48) |
| 80+ | 54,925 (19) | 8,172 (15) | 10,061 (15) | 1,067 (11) | 1,753 (13) | 21,683 (18) | 5,269 (16) | 6,672 (16) | 556 (11) | 969 (13) |
| **Sex** |  |  |  |  |  |  |  |  |  |  |
| Female | 139,010 (48) | 25,564 (47) | 30,918 (47) | 4,859 (51) | 6,913 (51) | 56,171 (46) | 15,750 (47) | 19,751 (47) | 2,496 (50) | 3,773 (50) |
| Male | 153,169 (52) | 28,654 (53) | 34,425 (53) | 4,658 (49) | 6,691 (49) | 66,368 (54) | 18,077 (53) | 22,255 (53) | 2,510 (50) | 3,808 (50) |
| Unknown | 0 (0) | <5 (<0.1) | 5 (<0.1) | 0 (0) | 0 (0) | 0 (0) | <5 (<0.1) | <5 (<0.1) | 0 (0) | 0 (0) |
| **Race** |  |  |  |  |  |  |  |  |  |  |
| Asian | 21,553 (7.4) | 1,376 (2.5) | 1,691 (2.6) | 284 (3.0) | 407 (3.0) | 10,523 (8.6) | 972 (2.9) | 1,214 (2.9) | 184 (3.7) | 255 (3.4) |
| Black | 34,511 (12) | 4,645 (8.6) | 5,616 (8.6) | 548 (5.8) | 834 (6.1) | 15,763 (13) | 2,832 (8.4) | 3,528 (8.4) | 296 (5.9) | 479 (6.3) |
| Other | 1,611 (0.6) | 5,037 (9.3) | 6,335 (9.7) | 1,323 (14) | 1,997 (15) | 679 (0.6) | 3,203 (9.5) | 4,175 (9.9) | 662 (13) | 1,087 (14) |
| Unknown | 765 (0.3) | 5,705 (11) | 7,253 (11) | 767 (8.1) | 1,135 (8.3) | 235 (0.2) | 3,904 (12) | 5,052 (12) | 427 (8.5) | 659 (8.7) |
| White | 233,739 (80) | 37,459 (69) | 44,453 (68) | 6,595 (69) | 9,231 (68) | 95,339 (78) | 22,917 (68) | 28,039 (67) | 3,437 (69) | 5,101 (67) |

| AJCC stage at diagnosis |  |  |  |  |  |  |  |  |  |  |
| --- | --- | --- | --- | --- | --- | --- | --- | --- | --- | --- |
| Stage 0 | 9 (<0.1) | 5 (<0.1) | 5 (<0.1) | <5 (<0.1) | <5 (<0.1) | NA | NA | NA | NA | NA |
| Stage I | 71,448 (24) | 4,688 (8.6) | 4,995 (7.6) | 1,155 (12) | 1,419 (10) | NA | NA | NA | NA | NA |
| Stage II | 19,762 (6.8) | 2,853 (5.3) | 3,072 (4.7) | 800 (8.4) | 1,020 (7.5) | NA | NA | NA | NA | NA |
| Stage III | 64,342 (22) | 11,309 (21) | 13,606 (21) | 2,199 (23) | 3,077 (23) | NA | NA | NA | NA | NA |
| Stage IV | 122,539 (42) | 33,828 (62) | 42,008 (64) | 5,006 (53) | 7,581 (56) | NA | NA | NA | NA | NA |
| Unknown | 14,079 (4.8) | 1,539 (2.8) | 1,662 (2.5) | 356 (3.7) | 506 (3.7) | NA | NA | NA | NA | NA |
| Year of initial diagnosis |  |  |  |  |  |  |  |  |  |  |
| 2011 | 36,193 (12) | 4,910 (9.1) | 4,910 (7.5) | 244 (2.6) | 244 (1.8) | 14,871 (12) | 2,707 (8.0) | 2,707 (6.4) | 82 (1.6) | 82 (1.1) |
| 2012 | 36,396 (12) | 5,942 (11) | 5,942 (9.1) | 381 (4.0) | 381 (2.8) | 15,249 (12) | 3,460 (10) | 3,460 (8.2) | 112 (2.2) | 112 (1.5) |
| 2013 | 36,526 (13) | 6,749 (12) | 6,749 (10) | 620 (6.5) | 620 (4.6) | 15,303 (12) | 4,140 (12) | 4,140 (9.9) | 280 (5.6) | 280 (3.7) |
| 2014 | 36,776 (13) | 7,211 (13) | 7,211 (11) | 984 (10) | 984 (7.2) | 15,460 (13) | 4,507 (13) | 4,507 (11) | 479 (9.6) | 479 (6.3) |
| 2015 | 37,119 (13) | 7,625 (14) | 7,625 (12) | 1,440 (15) | 1,440 (11) | 15,312 (12) | 4,796 (14) | 4,796 (11) | 744 (15) | 744 (9.8) |
| 2016 | 37,214 (13) | 7,484 (14) | 7,484 (11) | 1,663 (17) | 1,663 (12) | 16,222 (13) | 4,733 (14) | 4,733 (11) | 882 (18) | 882 (12) |
| 2017 | 37,708 (13) | 7,373 (14) | 7,373 (11) | 1,939 (20) | 1,939 (14) | 16,020 (13) | 4,862 (14) | 4,862 (12) | 1,092 (22) | 1,092 (14) |
| 2018 | 34,247 (12) | 6,928 (13) | 6,928 (11) | 2,246 (24) | 2,246 (17) | 14,102 (12) | 4,623 (14) | 4,623 (11) | 1,335 (27) | 1,335 (18) |
| 2019 | NA | NA | 6,187 (9.5) | NA | 2,077 (15) | NA | NA | 4,273 (10) | NA | 1,258 (17) |
| 2020 | NA | NA | 4,558 (7.0) | NA | 1,758 (13) | NA | NA | 3,597 (8.6) | NA | 1,135 (15) |
| 2021 | NA | NA | 381 (0.6) | NA | 252 (1.9) | NA | NA | 310 (0.7) | NA | 182 (2.4) |
| Abbreviations: AJCC, American Joint Committee on Cancer; CGDB, Flatiron Health-Foundation Medicine Clinico-Genomic Database; FHRD, Flatiron Health Research Database; NA, not applicable; NSCLC, non-small cell lung cancer; SEER, The Surveillance, Epidemiology, and End Results research database. |  |  |  |  |  |  |  |  |  |  |
| *Proportions of patients reported in the FHRD cohort are not directly comparable to the SEER and CGDB cohorts given that patients with advanced disease (at initial diagnosis or through the subsequent development of recurrence or metastasis) are included in the NSCLC FHRD whereas SEER and the NSCLC CGDB include patients of all stages regardless of advanced disease status. |  |  |  |  |  |  |  |  |  |  |
| Note: Reported percentages may not equal exactly to 100% due to rounding. |  |  |  |  |  |  |  |  |  |  |

**Table 19.**
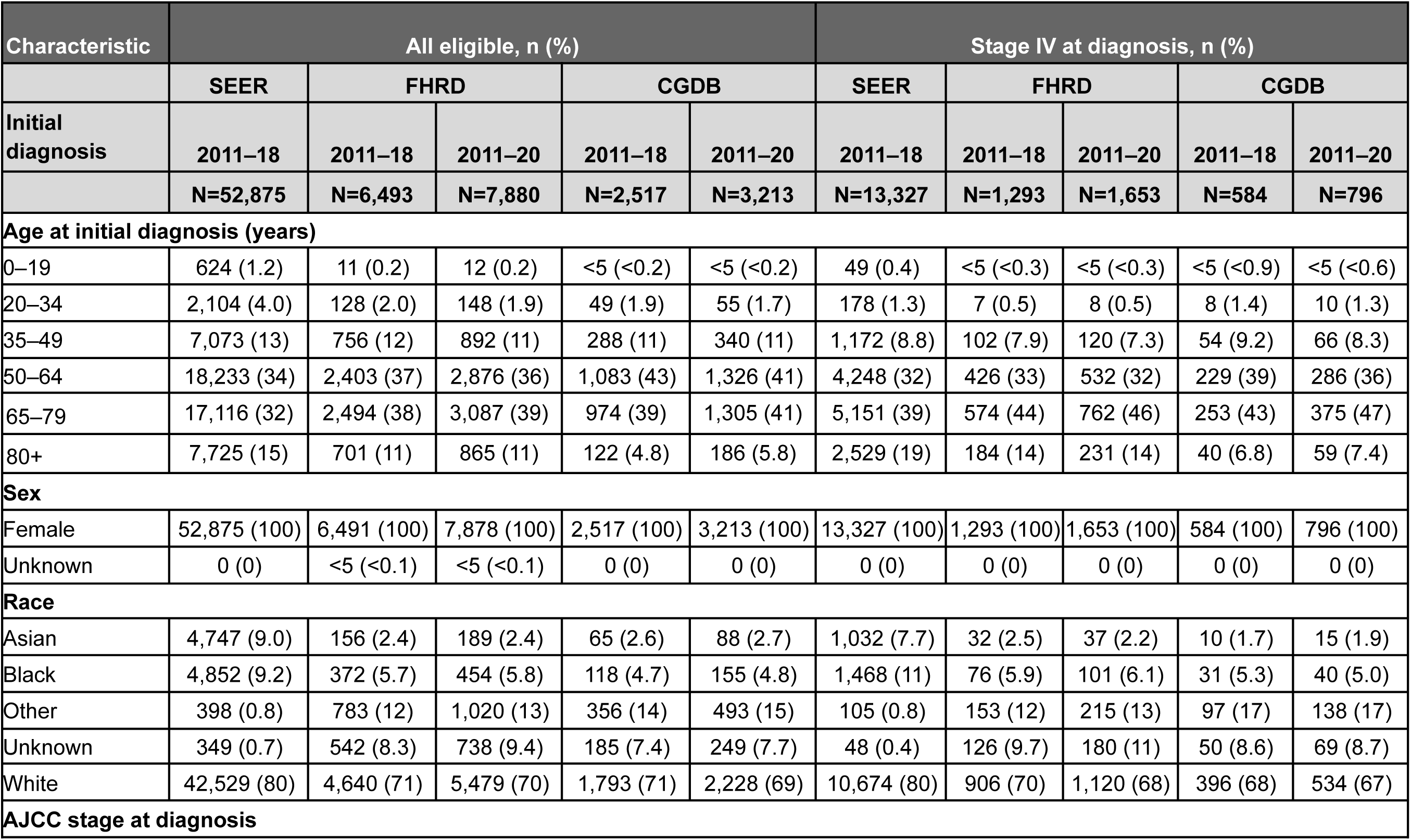
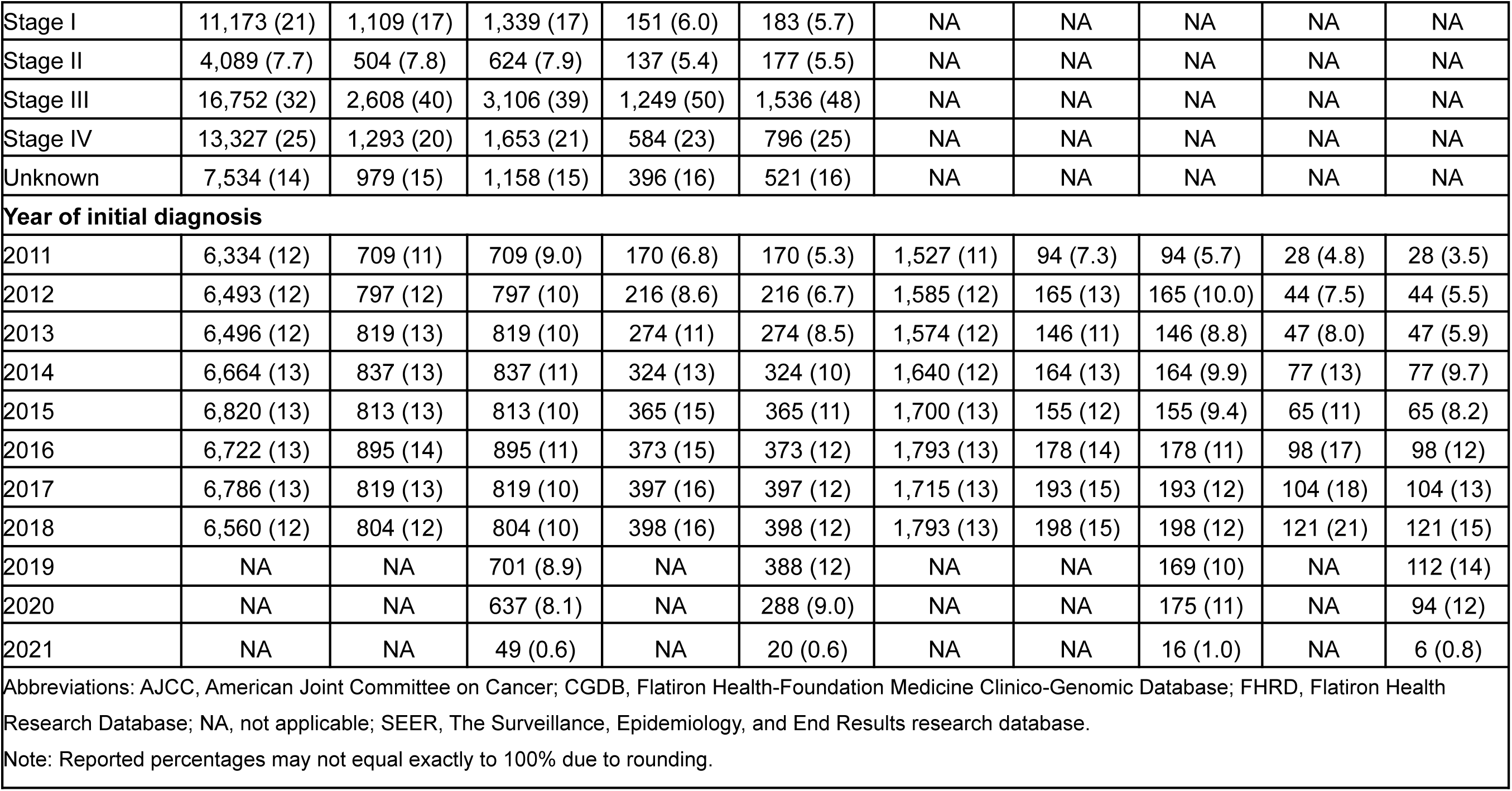
Ovarian Carcinoma – Comparison of Demographic and Clinical Characteristics Between SEER vs FHRD vs CGDB.

**Table 20.**
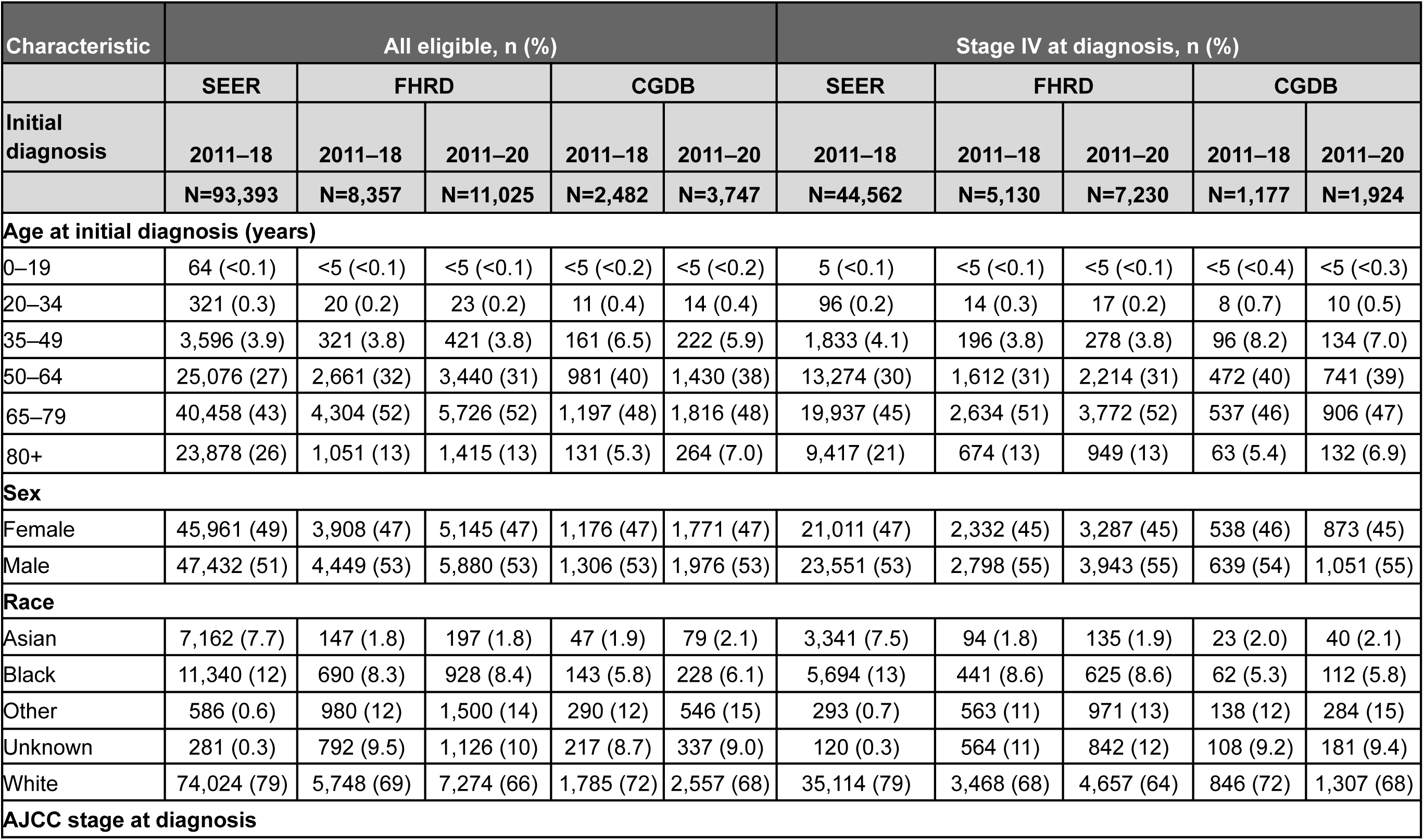
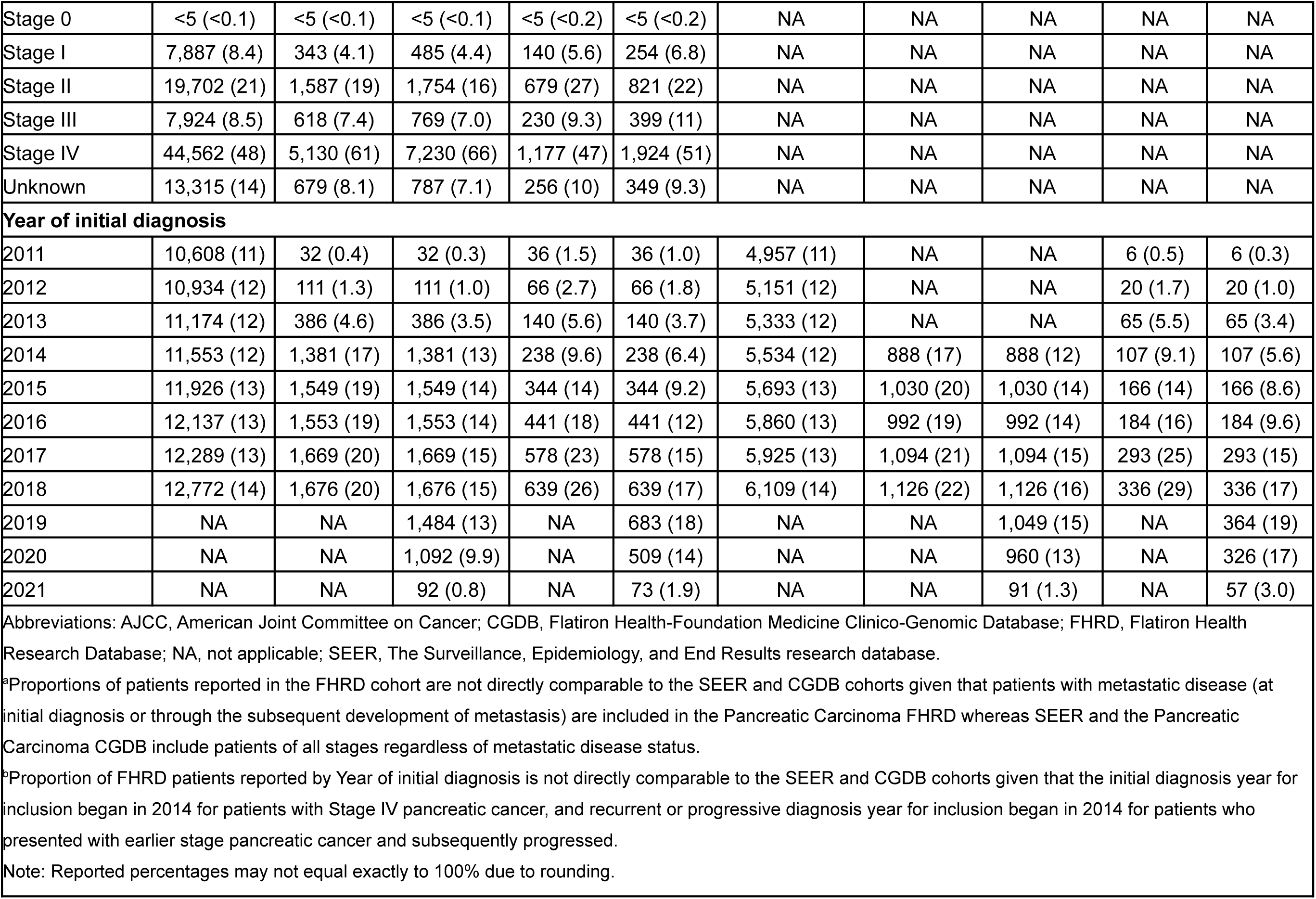
Pancreatic Adenocarcinoma – Comparison of Demographic and Clinical Characteristic Between SEER vs FHRD (metastatic at any time) vs CGDB^a,b^.

**Table 21.**
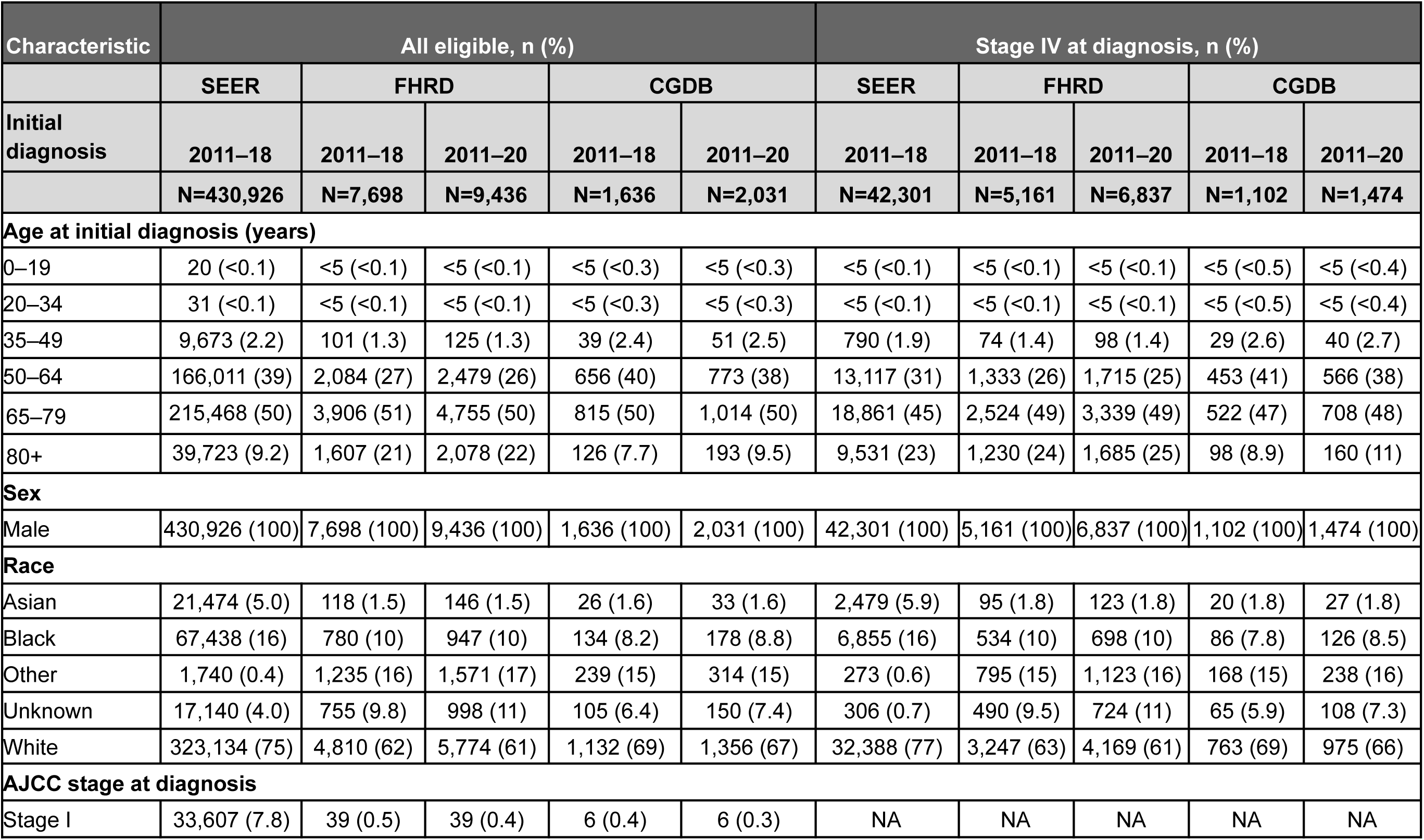
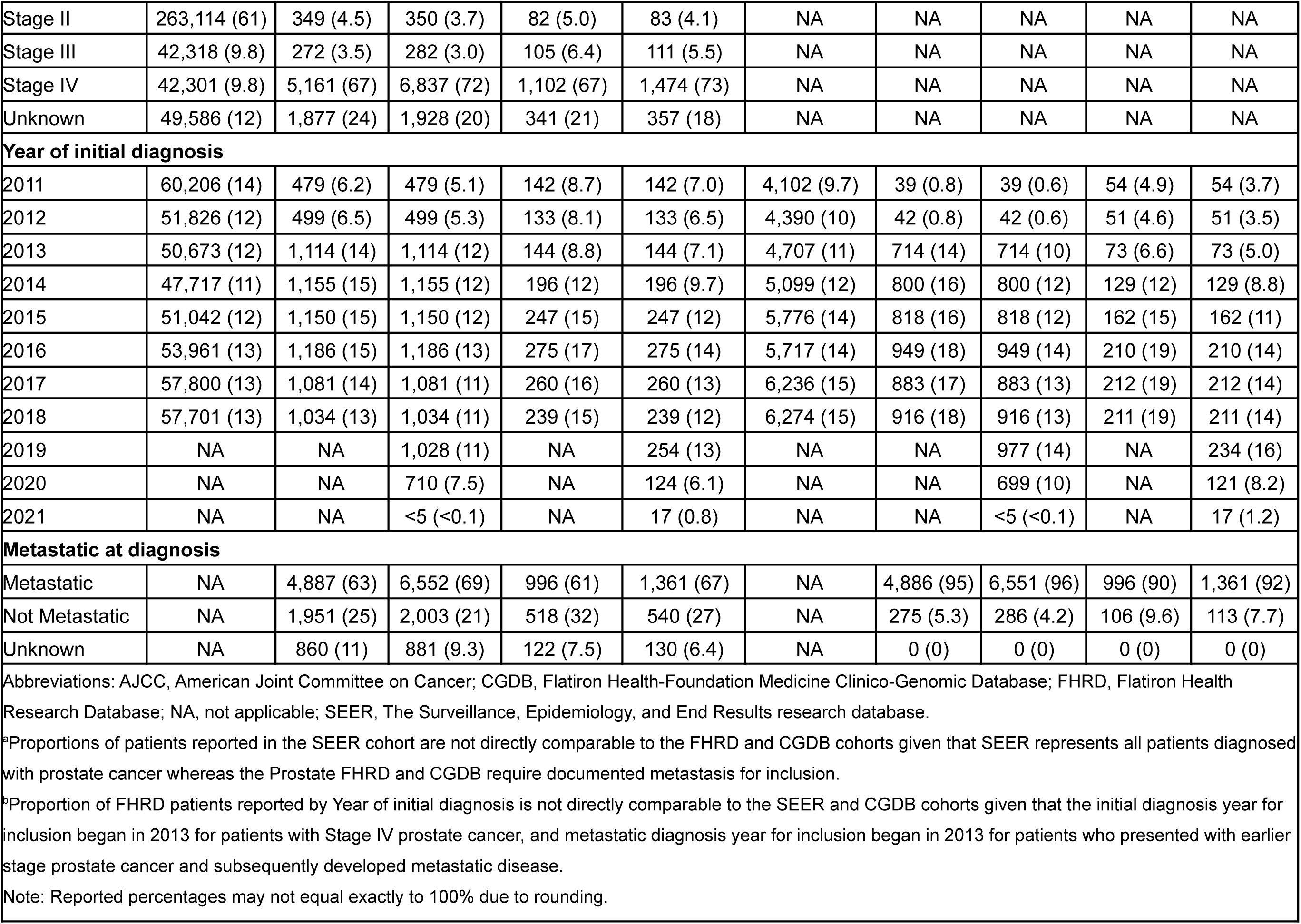
Prostate Cancer – Comparison of Demographic and Clinical Characteristics Between SEER (metastatic and non-metastatic at diagnosis) vs FHRD (metastatic at any time) vs CGDB (metastatic at any time)^a,b^.

**Table 22.**
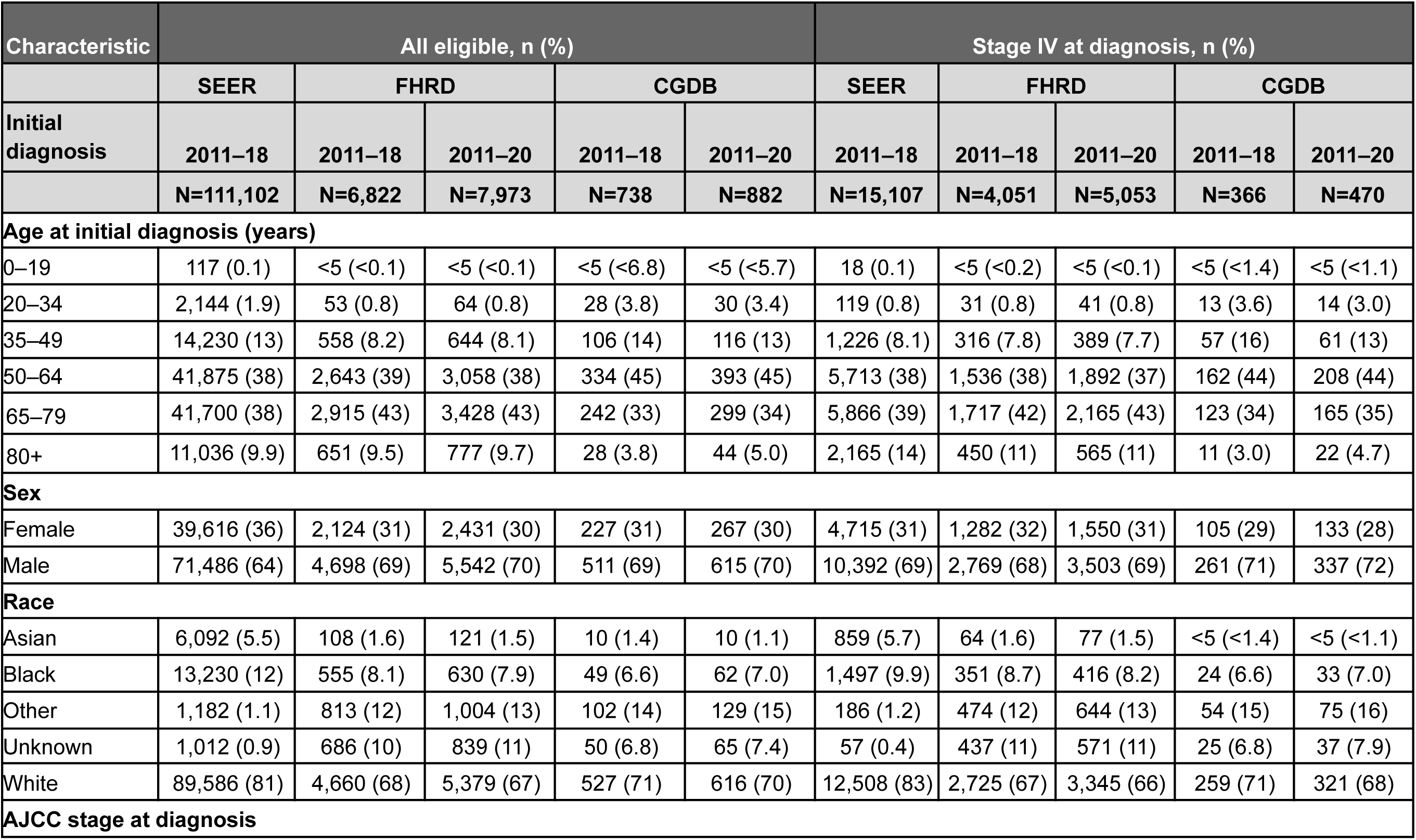
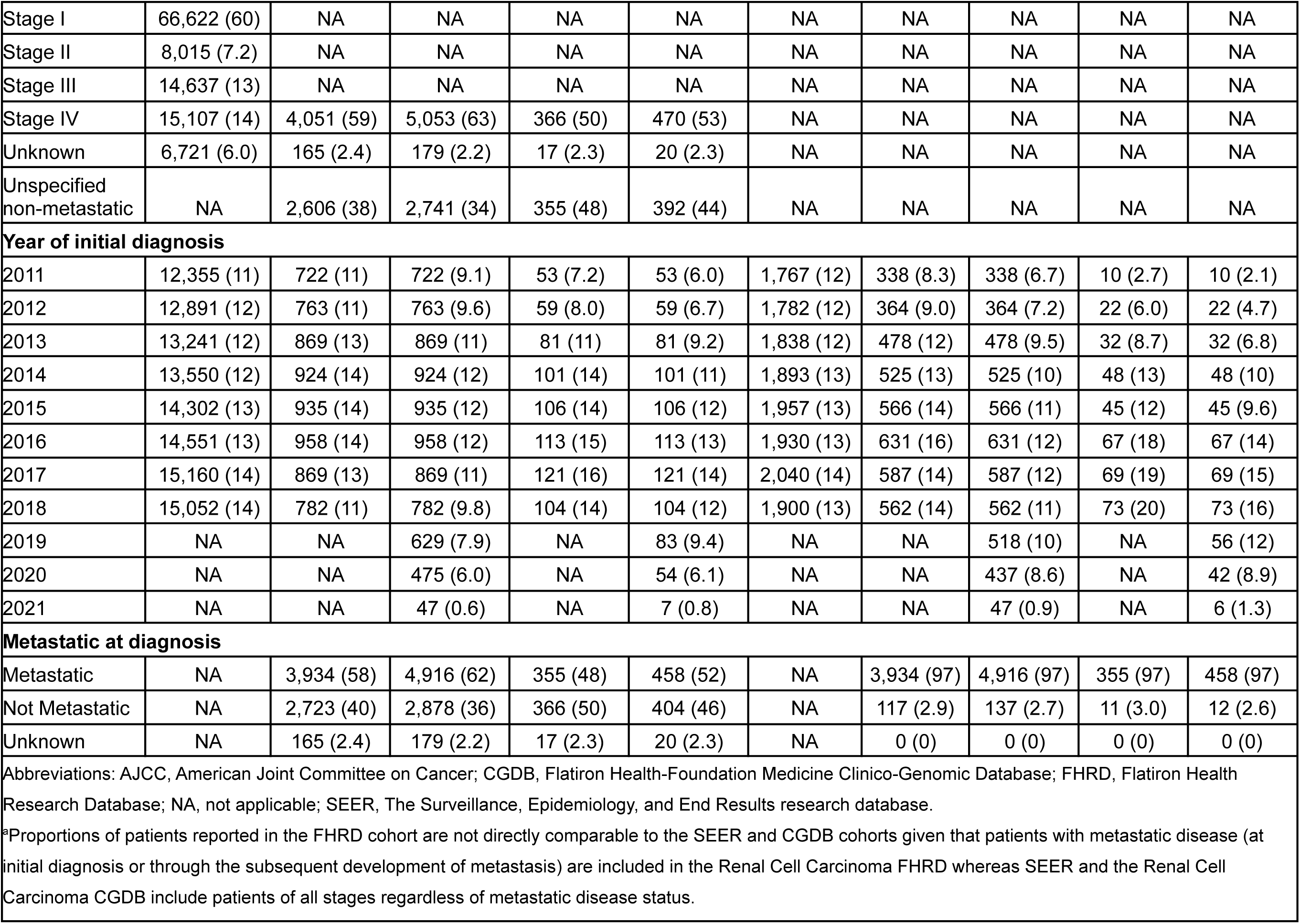
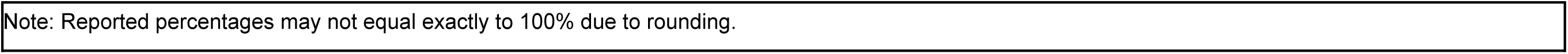
Renal Cell Carcinoma – Comparison of Demographic and Clinical Characteristics Between SEER vs FHRD (metastatic at any time) vs CGDB^a^.

| Characteristic | All eligible, n (%) |  |  |  |  | Stage IV at diagnosis, n (%) |  |  |  |  |
| --- | --- | --- | --- | --- | --- | --- | --- | --- | --- | --- |
|  | SEER | FHRD |  | CGDB |  | SEER | FHRD |  | CGDB |  |
| Initial diagnosis | 2011–18 | 2011–18 | 2011–20 | 2011–18 | 2011–20 | 2011–18 | 2011–18 | 2011–20 | 2011–18 | 2011–20 |
|  | N=111,102 | N=6,822 | N=7,973 | N=738 | N=882 | N=15,107 | N=4,051 | N=5,053 | N=366 | N=470 |
| Age at initial diagnosis (years) |  |  |  |  |  |  |  |  |  |  |
| 0–19 | 117 (0.1) | <5 (<0.1) | <5 (<0.1) | <5 (<6.8) | <5 (<5.7) | 18 (0.1) | <5 (<0.2) | <5 (<0.1) | <5 (<1.4) | <5 (<1.1) |
| 20–34 | 2,144 (1.9) | 53 (0.8) | 64 (0.8) | 28 (3.8) | 30 (3.4) | 119 (0.8) | 31 (0.8) | 41 (0.8) | 13 (3.6) | 14 (3.0) |
| 35–49 | 14,230 (13) | 558 (8.2) | 644 (8.1) | 106 (14) | 116 (13) | 1,226 (8.1) | 316 (7.8) | 389 (7.7) | 57 (16) | 61 (13) |
| 50–64 | 41,875 (38) | 2,643 (39) | 3,058 (38) | 334 (45) | 393 (45) | 5,713 (38) | 1,536 (38) | 1,892 (37) | 162 (44) | 208 (44) |
| 65–79 | 41,700 (38) | 2,915 (43) | 3,428 (43) | 242 (33) | 299 (34) | 5,866 (39) | 1,717 (42) | 2,165 (43) | 123 (34) | 165 (35) |
| 80+ | 11,036 (9.9) | 651 (9.5) | 777 (9.7) | 28 (3.8) | 44 (5.0) | 2,165 (14) | 450 (11) | 565 (11) | 11 (3.0) | 22 (4.7) |
| Sex |  |  |  |  |  |  |  |  |  |  |
| Female | 39,616 (36) | 2,124 (31) | 2,431 (30) | 227 (31) | 267 (30) | 4,715 (31) | 1,282 (32) | 1,550 (31) | 105 (29) | 133 (28) |
| Male | 71,486 (64) | 4,698 (69) | 5,542 (70) | 511 (69) | 615 (70) | 10,392 (69) | 2,769 (68) | 3,503 (69) | 261 (71) | 337 (72) |
| Race |  |  |  |  |  |  |  |  |  |  |
| Asian | 6,092 (5.5) | 108 (1.6) | 121 (1.5) | 10 (1.4) | 10 (1.1) | 859 (5.7) | 64 (1.6) | 77 (1.5) | <5 (<1.4) | <5 (<1.1) |
| Black | 13,230 (12) | 555 (8.1) | 630 (7.9) | 49 (6.6) | 62 (7.0) | 1,497 (9.9) | 351 (8.7) | 416 (8.2) | 24 (6.6) | 33 (7.0) |
| Other | 1,182 (1.1) | 813 (12) | 1,004 (13) | 102 (14) | 129 (15) | 186 (1.2) | 474 (12) | 644 (13) | 54 (15) | 75 (16) |
| Unknown | 1,012 (0.9) | 686 (10) | 839 (11) | 50 (6.8) | 65 (7.4) | 57 (0.4) | 437 (11) | 571 (11) | 25 (6.8) | 37 (7.9) |
| White | 89,586 (81) | 4,660 (68) | 5,379 (67) | 527 (71) | 616 (70) | 12,508 (83) | 2,725 (67) | 3,345 (66) | 259 (71) | 321 (68) |
| AJCC stage at diagnosis |  |  |  |  |  |  |  |  |  |  |
| Stage I | 66,622 (60) | NA | NA | NA | NA | NA | NA | NA | NA | NA |
| Stage II | 8,015 (7.2) | NA | NA | NA | NA | NA | NA | NA | NA | NA |
| Stage III | 14,637 (13) | NA | NA | NA | NA | NA | NA | NA | NA | NA |
| Stage IV | 15,107 (14) | 4,051 (59) | 5,053 (63) | 366 (50) | 470 (53) | NA | NA | NA | NA | NA |
| Unknown | 6,721 (6.0) | 165 (2.4) | 179 (2.2) | 17 (2.3) | 20 (2.3) | NA | NA | NA | NA | NA |
| Unspecified non-metastatic | NA | 2,606 (38) | 2,741 (34) | 355 (48) | 392 (44) | NA | NA | NA | NA | NA |
| **Year of initial diagnosis** |  |  |  |  |  |  |  |  |  |  |
| 2011 | 12,355 (11) | 722 (11) | 722 (9.1) | 53 (7.2) | 53 (6.0) | 1,767 (12) | 338 (8.3) | 338 (6.7) | 10 (2.7) | 10 (2.1) |
| 2012 | 12,891 (12) | 763 (11) | 763 (9.6) | 59 (8.0) | 59 (6.7) | 1,782 (12) | 364 (9.0) | 364 (7.2) | 22 (6.0) | 22 (4.7) |
| 2013 | 13,241 (12) | 869 (13) | 869 (11) | 81 (11) | 81 (9.2) | 1,838 (12) | 478 (12) | 478 (9.5) | 32 (8.7) | 32 (6.8) |
| 2014 | 13,550 (12) | 924 (14) | 924 (12) | 101 (14) | 101 (11) | 1,893 (13) | 525 (13) | 525 (10) | 48 (13) | 48 (10) |
| 2015 | 14,302 (13) | 935 (14) | 935 (12) | 106 (14) | 106 (12) | 1,957 (13) | 566 (14) | 566 (11) | 45 (12) | 45 (9.6) |
| 2016 | 14,551 (13) | 958 (14) | 958 (12) | 113 (15) | 113 (13) | 1,930 (13) | 631 (16) | 631 (12) | 67 (18) | 67 (14) |
| 2017 | 15,160 (14) | 869 (13) | 869 (11) | 121 (16) | 121 (14) | 2,040 (14) | 587 (14) | 587 (12) | 69 (19) | 69 (15) |
| 2018 | 15,052 (14) | 782 (11) | 782 (9.8) | 104 (14) | 104 (12) | 1,900 (13) | 562 (14) | 562 (11) | 73 (20) | 73 (16) |
| 2019 | NA | NA | 629 (7.9) | NA | 83 (9.4) | NA | NA | 518 (10) | NA | 56 (12) |
| 2020 | NA | NA | 475 (6.0) | NA | 54 (6.1) | NA | NA | 437 (8.6) | NA | 42 (8.9) |
| 2021 | NA | NA | 47 (0.6) | NA | 7 (0.8) | NA | NA | 47 (0.9) | NA | 6 (1.3) |
| **Metastatic at diagnosis** |  |  |  |  |  |  |  |  |  |  |
| Metastatic | NA | 3,934 (58) | 4,916 (62) | 355 (48) | 458 (52) | NA | 3,934 (97) | 4,916 (97) | 355 (97) | 458 (97) |
| Not Metastatic | NA | 2,723 (40) | 2,878 (36) | 366 (50) | 404 (46) | NA | 117 (2.9) | 137 (2.7) | 11 (3.0) | 12 (2.6) |
| Unknown | NA | 165 (2.4) | 179 (2.2) | 17 (2.3) | 20 (2.3) | NA | 0 (0) | 0 (0) | 0 (0) | 0 (0) |

Abbreviations: AJCC, American Joint Committee on Cancer; CGDB, Flatiron Health-Foundation Medicine Clinico-Genomic Database; FHRD, Flatiron Health Research Database; NA, not applicable; SEER, The Surveillance, Epidemiology, and End Results research database.
\*Proportions of patients reported in the FHRD cohort are not directly comparable to the SEER and CGDB cohorts given that patients with metastatic disease (at initial diagnosis or through the subsequent development of metastasis) are included in the Renal Cell Carcinoma FHRD whereas SEER and the Renal Cell Carcinoma CGDB include patients of all stages regardless of metastatic disease status.
Note: Reported percentages may not equal exactly to 100% due to rounding.

**Table 23.**
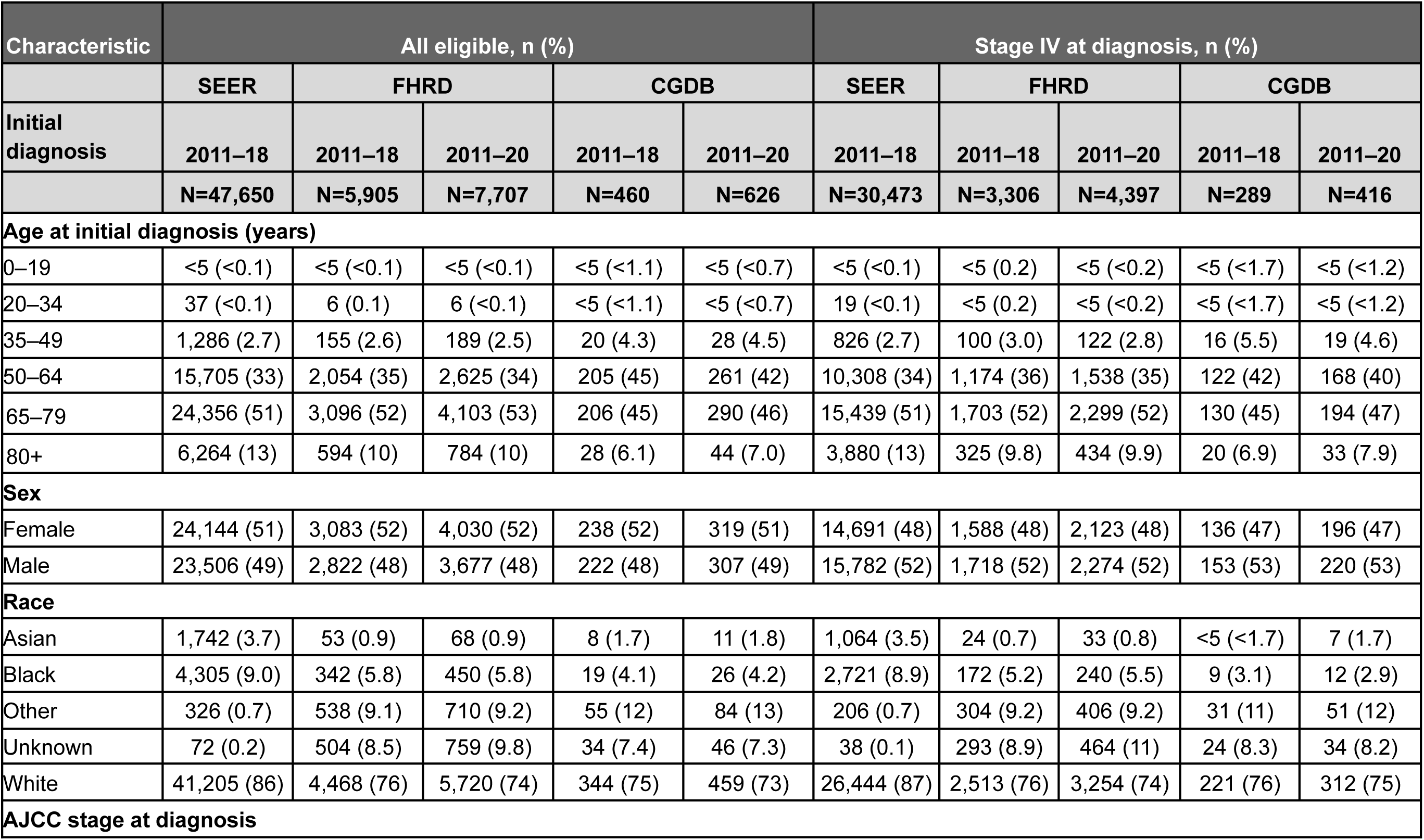
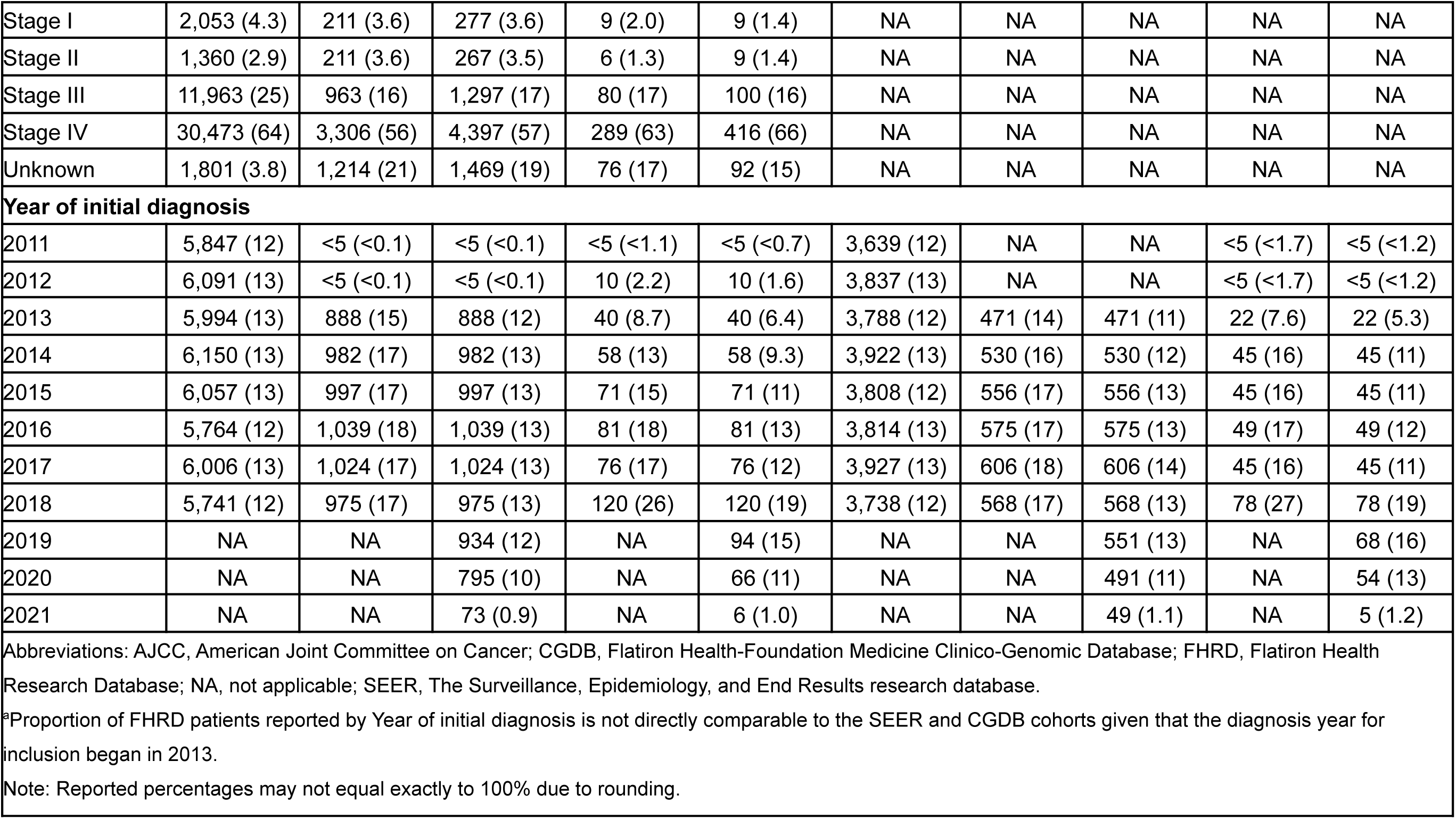
Small Cell Lung Cancer – Comparison of Demographic and Clinical Characteristics Between SEER vs FHRD vs CGDB^a^.

| Characteristic | All eligible, n (%) |  |  |  |  | Stage IV at diagnosis, n (%) |  |  |  |  |
| --- | --- | --- | --- | --- | --- | --- | --- | --- | --- | --- |
|  | SEER | FHRD |  | CGDB |  | SEER | FHRD |  | CGDB |  |
| Initial diagnosis | 2011–18 | 2011–18 | 2011–20 | 2011–18 | 2011–20 | 2011–18 | 2011–18 | 2011–20 | 2011–18 | 2011–20 |
|  | N=47,650 | N=5,905 | N=7,707 | N=460 | N=626 | N=30,473 | N=3,306 | N=4,397 | N=289 | N=416 |
| Age at initial diagnosis (years) |  |  |  |  |  |  |  |  |  |  |
| 0–19 | <5 (<0.1) | <5 (<0.1) | <5 (<0.1) | <5 (<1.1) | <5 (<0.7) | <5 (<0.1) | <5 (0.2) | <5 (<0.2) | <5 (<1.7) | <5 (<1.2) |
| 20–34 | 37 (<0.1) | 6 (0.1) | 6 (<0.1) | <5 (<1.1) | <5 (<0.7) | 19 (<0.1) | <5 (0.2) | <5 (<0.2) | <5 (<1.7) | <5 (<1.2) |
| 35–49 | 1,286 (2.7) | 155 (2.6) | 189 (2.5) | 20 (4.3) | 28 (4.5) | 826 (2.7) | 100 (3.0) | 122 (2.8) | 16 (5.5) | 19 (4.6) |
| 50–64 | 15,705 (33) | 2,054 (35) | 2,625 (34) | 205 (45) | 261 (42) | 10,308 (34) | 1,174 (36) | 1,538 (35) | 122 (42) | 168 (40) |
| 65–79 | 24,356 (51) | 3,096 (52) | 4,103 (53) | 206 (45) | 290 (46) | 15,439 (51) | 1,703 (52) | 2,299 (52) | 130 (45) | 194 (47) |
| 80+ | 6,264 (13) | 594 (10) | 784 (10) | 28 (6.1) | 44 (7.0) | 3,880 (13) | 325 (9.8) | 434 (9.9) | 20 (6.9) | 33 (7.9) |
| Sex |  |  |  |  |  |  |  |  |  |  |
| Female | 24,144 (51) | 3,083 (52) | 4,030 (52) | 238 (52) | 319 (51) | 14,691 (48) | 1,588 (48) | 2,123 (48) | 136 (47) | 196 (47) |
| Male | 23,506 (49) | 2,822 (48) | 3,677 (48) | 222 (48) | 307 (49) | 15,782 (52) | 1,718 (52) | 2,274 (52) | 153 (53) | 220 (53) |
| Race |  |  |  |  |  |  |  |  |  |  |
| Asian | 1,742 (3.7) | 53 (0.9) | 68 (0.9) | 8 (1.7) | 11 (1.8) | 1,064 (3.5) | 24 (0.7) | 33 (0.8) | <5 (<1.7) | 7 (1.7) |
| Black | 4,305 (9.0) | 342 (5.8) | 450 (5.8) | 19 (4.1) | 26 (4.2) | 2,721 (8.9) | 172 (5.2) | 240 (5.5) | 9 (3.1) | 12 (2.9) |
| Other | 326 (0.7) | 538 (9.1) | 710 (9.2) | 55 (12) | 84 (13) | 206 (0.7) | 304 (9.2) | 406 (9.2) | 31 (11) | 51 (12) |
| Unknown | 72 (0.2) | 504 (8.5) | 759 (9.8) | 34 (7.4) | 46 (7.3) | 38 (0.1) | 293 (8.9) | 464 (11) | 24 (8.3) | 34 (8.2) |
| White | 41,205 (86) | 4,468 (76) | 5,720 (74) | 344 (75) | 459 (73) | 26,444 (87) | 2,513 (76) | 3,254 (74) | 221 (76) | 312 (75) |
| AJCC stage at diagnosis |  |  |  |  |  |  |  |  |  |  |
| Stage I | 2,053 (4.3) | 211 (3.6) | 277 (3.6) | 9 (2.0) | 9 (1.4) | NA | NA | NA | NA | NA |
| Stage II | 1,360 (2.9) | 211 (3.6) | 267 (3.5) | 6 (1.3) | 9 (1.4) | NA | NA | NA | NA | NA |
| Stage III | 11,963 (25) | 963 (16) | 1,297 (17) | 80 (17) | 100 (16) | NA | NA | NA | NA | NA |
| Stage IV | 30,473 (64) | 3,306 (56) | 4,397 (57) | 289 (63) | 416 (66) | NA | NA | NA | NA | NA |
| Unknown | 1,801 (3.8) | 1,214 (21) | 1,469 (19) | 76 (17) | 92 (15) | NA | NA | NA | NA | NA |
| Year of initial diagnosis |  |  |  |  |  |  |  |  |  |  |
| 2011 | 5,847 (12) | <5 (<0.1) | <5 (<0.1) | <5 (<1.1) | <5 (<0.7) | 3,639 (12) | NA | NA | <5 (<1.7) | <5 (<1.2) |
| 2012 | 6,091 (13) | <5 (<0.1) | <5 (<0.1) | 10 (2.2) | 10 (1.6) | 3,837 (13) | NA | NA | <5 (<1.7) | <5 (<1.2) |
| 2013 | 5,994 (13) | 888 (15) | 888 (12) | 40 (8.7) | 40 (6.4) | 3,788 (12) | 471 (14) | 471 (11) | 22 (7.6) | 22 (5.3) |
| 2014 | 6,150 (13) | 982 (17) | 982 (13) | 58 (13) | 58 (9.3) | 3,922 (13) | 530 (16) | 530 (12) | 45 (16) | 45 (11) |
| 2015 | 6,057 (13) | 997 (17) | 997 (13) | 71 (15) | 71 (11) | 3,808 (12) | 556 (17) | 556 (13) | 45 (16) | 45 (11) |
| 2016 | 5,764 (12) | 1,039 (18) | 1,039 (13) | 81 (18) | 81 (13) | 3,814 (13) | 575 (17) | 575 (13) | 49 (17) | 49 (12) |
| 2017 | 6,006 (13) | 1,024 (17) | 1,024 (13) | 76 (17) | 76 (12) | 3,927 (13) | 606 (18) | 606 (14) | 45 (16) | 45 (11) |
| 2018 | 5,741 (12) | 975 (17) | 975 (13) | 120 (26) | 120 (19) | 3,738 (12) | 568 (17) | 568 (13) | 78 (27) | 78 (19) |
| 2019 | NA | NA | 934 (12) | NA | 94 (15) | NA | NA | 551 (13) | NA | 68 (16) |
| 2020 | NA | NA | 795 (10) | NA | 66 (11) | NA | NA | 491 (11) | NA | 54 (13) |
| 2021 | NA | NA | 73 (0.9) | NA | 6 (1.0) | NA | NA | 49 (1.1) | NA | 5 (1.2) |

Abbreviations: AJCC, American Joint Committee on Cancer; CGDB, Flatiron Health-Foundation Medicine Clinico-Genomic Database; FHRD, Flatiron Health Research Database; NA, not applicable; SEER, The Surveillance, Epidemiology, and End Results research database.
<sup>a</sup>Proportion of FHRD patients reported by Year of initial diagnosis is not directly comparable to the SEER and CGDB cohorts given that the diagnosis year for inclusion began in 2013.
Note: Reported percentages may not equal exactly to 100% due to rounding.

**Tables 24–30** present CGDB vs SEER comparisons for the following cancer types: breast cancer (all patients) (**Table 24**), cholangiocarcinoma (**Table 25**), glioma (**Table 26**), malignant pleural mesothelioma (**Table 27**), all solid tumors (**Table 28**), all hematological malignancies (**Table 29**), and all patients in each database (**Table 30**).

**Table 24.**
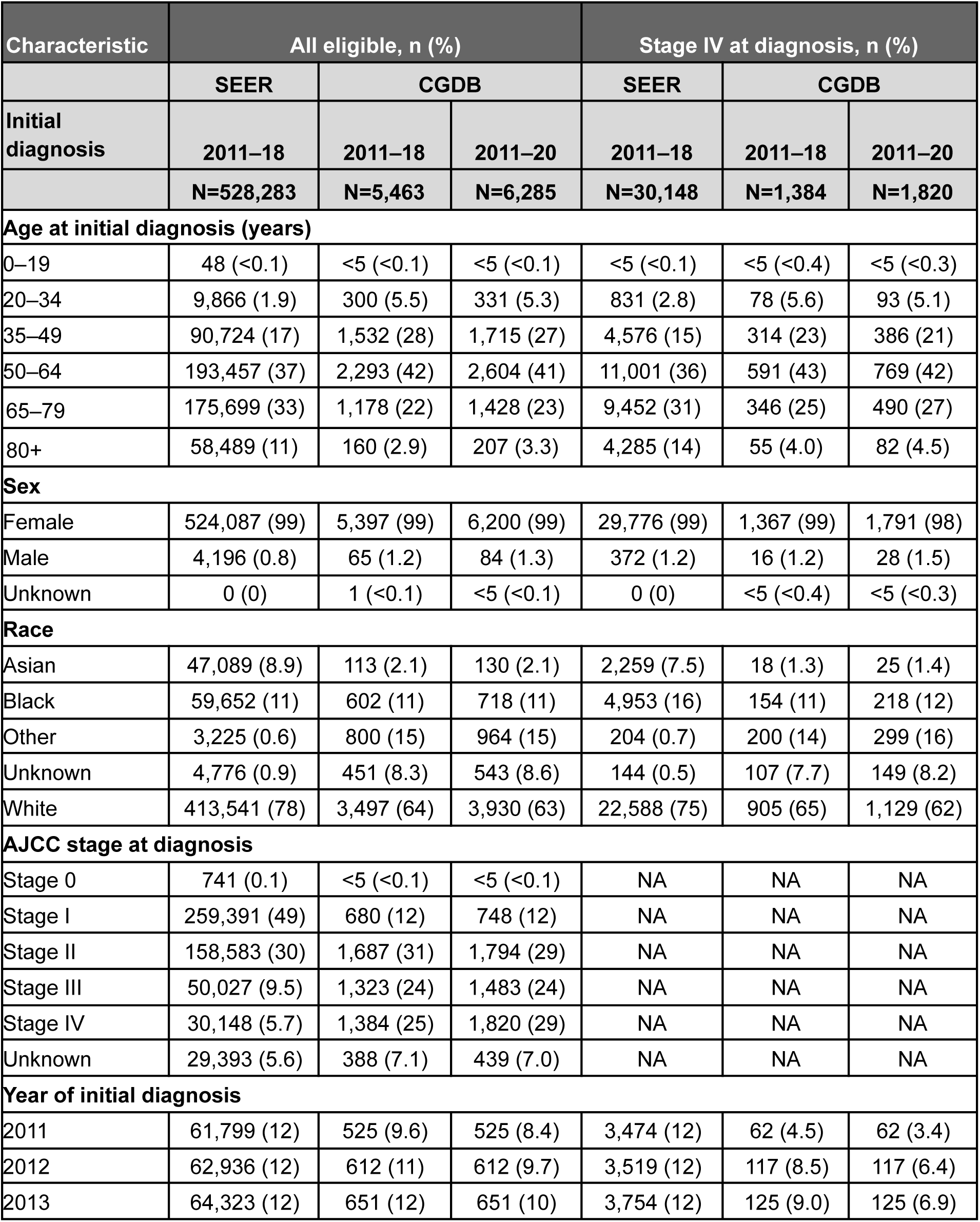
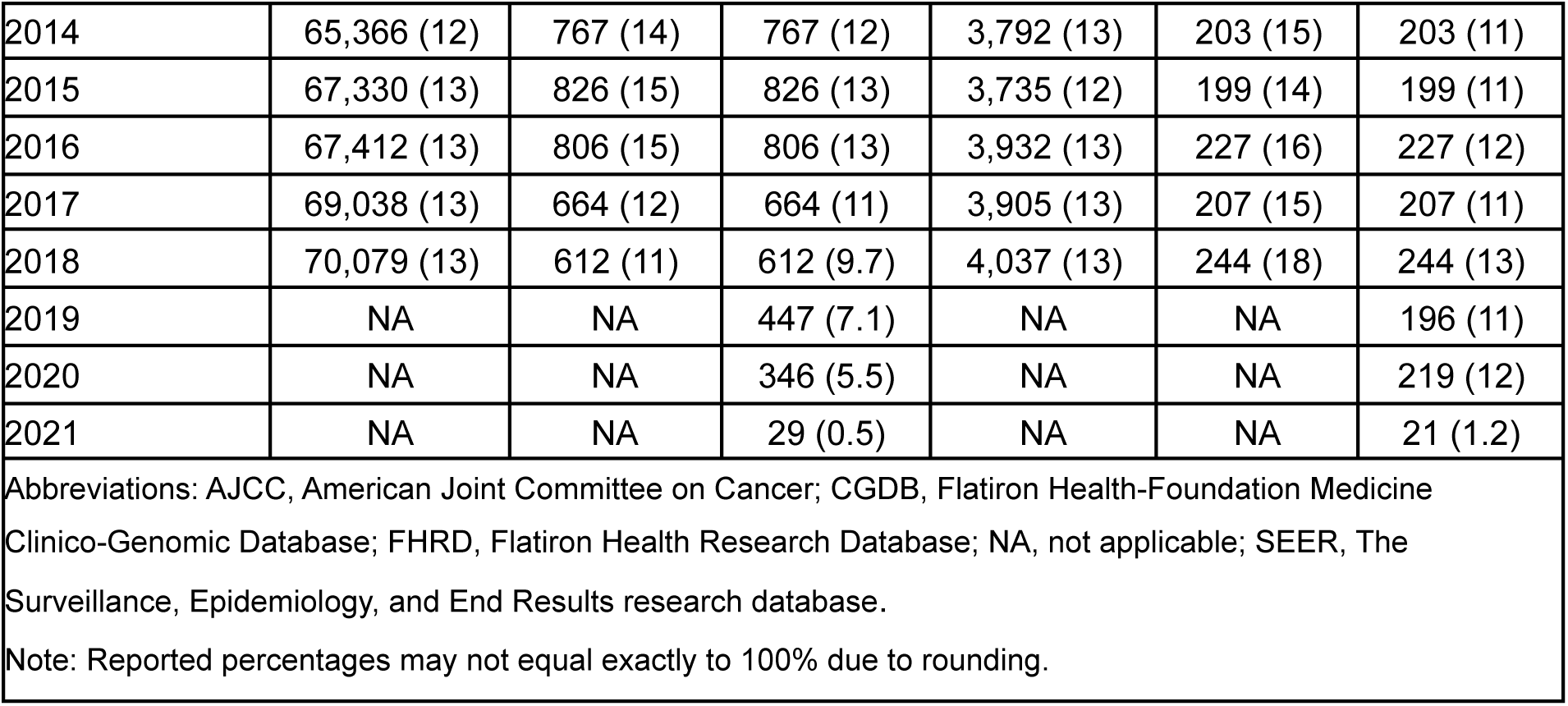
Breast Cancer (All) – Comparison of Demographic and Clinical Characteristics Between SEER vs CGDB.

| Characteristic | All eligible, n (%) |  |  | Stage IV at diagnosis, n (%) |  |  |
| --- | --- | --- | --- | --- | --- | --- |
|  | SEER | CGDB |  | SEER | CGDB |  |
| Initial diagnosis | 2011–18 | 2011–18 | 2011–20 | 2011–18 | 2011–18 | 2011–20 |
|  | N=528,283 | N=5,463 | N=6,285 | N=30,148 | N=1,384 | N=1,820 |
| **Age at initial diagnosis (years)** |  |  |  |  |  |  |
| 0–19 | 48 (<0.1) | <5 (<0.1) | <5 (<0.1) | <5 (<0.1) | <5 (<0.4) | <5 (<0.3) |
| 20–34 | 9,866 (1.9) | 300 (5.5) | 331 (5.3) | 831 (2.8) | 78 (5.6) | 93 (5.1) |
| 35–49 | 90,724 (17) | 1,532 (28) | 1,715 (27) | 4,576 (15) | 314 (23) | 386 (21) |
| 50–64 | 193,457 (37) | 2,293 (42) | 2,604 (41) | 11,001 (36) | 591 (43) | 769 (42) |
| 65–79 | 175,699 (33) | 1,178 (22) | 1,428 (23) | 9,452 (31) | 346 (25) | 490 (27) |
| 80+ | 58,489 (11) | 160 (2.9) | 207 (3.3) | 4,285 (14) | 55 (4.0) | 82 (4.5) |
| **Sex** |  |  |  |  |  |  |
| Female | 524,087 (99) | 5,397 (99) | 6,200 (99) | 29,776 (99) | 1,367 (99) | 1,791 (98) |
| Male | 4,196 (0.8) | 65 (1.2) | 84 (1.3) | 372 (1.2) | 16 (1.2) | 28 (1.5) |
| Unknown | 0 (0) | 1 (<0.1) | <5 (<0.1) | 0 (0) | <5 (<0.4) | <5 (<0.3) |
| **Race** |  |  |  |  |  |  |
| Asian | 47,089 (8.9) | 113 (2.1) | 130 (2.1) | 2,259 (7.5) | 18 (1.3) | 25 (1.4) |
| Black | 59,652 (11) | 602 (11) | 718 (11) | 4,953 (16) | 154 (11) | 218 (12) |
| Other | 3,225 (0.6) | 800 (15) | 964 (15) | 204 (0.7) | 200 (14) | 299 (16) |
| Unknown | 4,776 (0.9) | 451 (8.3) | 543 (8.6) | 144 (0.5) | 107 (7.7) | 149 (8.2) |
| White | 413,541 (78) | 3,497 (64) | 3,930 (63) | 22,588 (75) | 905 (65) | 1,129 (62) |
| **AJCC stage at diagnosis** |  |  |  |  |  |  |
| Stage 0 | 741 (0.1) | <5 (<0.1) | <5 (<0.1) | NA | NA | NA |
| Stage I | 259,391 (49) | 680 (12) | 748 (12) | NA | NA | NA |
| Stage II | 158,583 (30) | 1,687 (31) | 1,794 (29) | NA | NA | NA |
| Stage III | 50,027 (9.5) | 1,323 (24) | 1,483 (24) | NA | NA | NA |
| Stage IV | 30,148 (5.7) | 1,384 (25) | 1,820 (29) | NA | NA | NA |
| Unknown | 29,393 (5.6) | 388 (7.1) | 439 (7.0) | NA | NA | NA |
| **Year of initial diagnosis** |  |  |  |  |  |  |
| 2011 | 61,799 (12) | 525 (9.6) | 525 (8.4) | 3,474 (12) | 62 (4.5) | 62 (3.4) |
| 2012 | 62,936 (12) | 612 (11) | 612 (9.7) | 3,519 (12) | 117 (8.5) | 117 (6.4) |
| 2013 | 64,323 (12) | 651 (12) | 651 (10) | 3,754 (12) | 125 (9.0) | 125 (6.9) |
| 2014 | 65,366 (12) | 767 (14) | 767 (12) | 3,792 (13) | 203 (15) | 203 (11) |
| 2015 | 67,330 (13) | 826 (15) | 826 (13) | 3,735 (12) | 199 (14) | 199 (11) |
| 2016 | 67,412 (13) | 806 (15) | 806 (13) | 3,932 (13) | 227 (16) | 227 (12) |
| 2017 | 69,038 (13) | 664 (12) | 664 (11) | 3,905 (13) | 207 (15) | 207 (11) |
| 2018 | 70,079 (13) | 612 (11) | 612 (9.7) | 4,037 (13) | 244 (18) | 244 (13) |
| 2019 | NA | NA | 447 (7.1) | NA | NA | 196 (11) |
| 2020 | NA | NA | 346 (5.5) | NA | NA | 219 (12) |
| 2021 | NA | NA | 29 (0.5) | NA | NA | 21 (1.2) |

Abbreviations: AJCC, American Joint Committee on Cancer; CGDB, Flatiron Health-Foundation Medicine Clinico-Genomic Database; FHRD, Flatiron Health Research Database; NA, not applicable; SEER, The Surveillance, Epidemiology, and End Results research database.
Note: Reported percentages may not equal exactly to 100% due to rounding.

**Table 25.** Cholangiocarcinoma – Comparison of Demographic and Clinical Characteristics Between SEER vs CGDB.

| Characteristic | All eligible, n (%) |  |  | Stage IV at diagnosis, n (%) |  |  |
| --- | --- | --- | --- | --- | --- | --- |
|  | SEER | CGDB |  | SEER | CGDB |  |
| Initial diagnosis | 2011–18 | 2011–18 | 2011–20 | 2011–18 | 2011–18 | 2011–20 |
|  | N=17,620 | N=1,160 | N=1,723 | N=6,235 | N=534 | N=848 |
| **Age at initial diagnosis (years)** |  |  |  |  |  |  |
| 0–19 | 11 (<0.1) | <5 (<0.4) | <5 (<0.3) | <5 (<0.1) | <5 (<0.9) | <5 (<0.6) |
| 20–34 | 147 (0.8) | 15 (1.3) | 22 (1.3) | 72 (1.2) | 7 (1.3) | 11 (1.3) |
| 35–49 | 928 (5.3) | 127 (11) | 155 (9.0) | 434 (7.0) | 61 (11) | 80 (9.4) |
| 50–64 | 4,938 (28) | 452 (39) | 636 (37) | 2,082 (33) | 208 (39) | 316 (37) |
| 65–79 | 7,556 (43) | 510 (44) | 794 (46) | 2,748 (44) | 233 (44) | 385 (45) |
| 80+ | 4,040 (23) | 56 (4.8) | 116 (6.7) | 898 (14) | 25 (4.7) | 56 (6.6) |
| **Sex** |  |  |  |  |  |  |
| Female | 8,208 (47) | 603 (52) | 883 (51) | 3,006 (48) | 273 (51) | 438 (52) |
| Male | 9,412 (53) | 557 (48) | 840 (49) | 3,229 (52) | 261 (49) | 410 (48) |
| **Race** |  |  |  |  |  |  |
| Asian | 2,168 (12) | 46 (4.0) | 57 (3.3) | 751 (12) | 18 (3.4) | 22 (2.6) |
| Black | 1,480 (8.4) | 66 (5.7) | 105 (6.1) | 535 (8.6) | 28 (5.2) | 52 (6.1) |
| Other | 144 (0.8) | 128 (11) | 263 (15) | 45 (0.7) | 55 (10) | 117 (14) |
| Unknown | 50 (0.3) | 97 (8.4) | 155 (9.0) | 11 (0.2) | 44 (8.2) | 80 (9.4) |
| White | 13,778 (78) | 823 (71) | 1,143 (66) | 4,893 (78) | 389 (73) | 577 (68) |
| **AJCC stage at diagnosis** |  |  |  |  |  |  |
| Stage I | 2,757 (16) | 47 (4.1) | 68 (3.9) | NA | NA | NA |
| Stage II | 2,458 (14) | 126 (11) | 170 (9.9) | NA | NA | NA |
| Stage III | 2,013 (11) | 90 (7.8) | 148 (8.6) | NA | NA | NA |
| Stage IV | 6,235 (35) | 534 (46) | 848 (49) | NA | NA | NA |
| Unknown | 4,157 (24) | 363 (31) | 489 (28) | NA | NA | NA |
| **Year of initial diagnosis** |  |  |  |  |  |  |
| 2011 | 1,734 (9.8) | 25 (2.2) | 25 (1.5) | 525 (8.4) | 7 (1.3) | 7 (0.8) |
| 2012 | 1,830 (10) | 41 (3.5) | 41 (2.4) | 569 (9.1) | 9 (1.7) | 9 (1.1) |
| 2013 | 1,989 (11) | 66 (5.7) | 66 (3.8) | 671 (11) | 24 (4.5) | 24 (2.8) |
| 2014 | 2,148 (12) | 116 (10) | 116 (6.7) | 770 (12) | 49 (9.2) | 49 (5.8) |
| 2015 | 2,275 (13) | 161 (14) | 161 (9.3) | 829 (13) | 70 (13) | 70 (8.3) |
| 2016 | 2,402 (14) | 200 (17) | 200 (12) | 960 (15) | 93 (17) | 93 (11) |
| 2017 | 2,632 (15) | 247 (21) | 247 (14) | 1,156 (19) | 129 (24) | 129 (15) |
| 2018 | 2,610 (15) | 304 (26) | 304 (18) | 755 (12) | 153 (29) | 153 (18) |
| 2019 | NA | NA | 295 (17) | NA | NA | 159 (19) |
| 2020 | NA | NA | 237 (14) | NA | NA | 137 (16) |
| 2021 | NA | NA | 31 (1.8) | NA | NA | 18 (2.1) |

Abbreviations: AJCC, American Joint Committee on Cancer; CGDB, Flatiron Health-Foundation Medicine Clinico-Genomic Database; FHRD, Flatiron Health Research Database; NA, not applicable; SEER, The Surveillance, Epidemiology, and End Results research database.
Note: Reported percentages may not equal exactly to 100% due to rounding.

**Table 26.** Glioma – Comparison of Demographic and Clinical Characteristics Between SEER vs CGDB.

| Characteristic | All eligible, n (%) |  |  |
| --- | --- | --- | --- |
|  | SEER | CGDB |  |
| Initial diagnosis | 2011–18 | 2011–18 | 2011–20 |
|  | N=42,277 | N=1,087 | N=1,311 |
| **Age at initial diagnosis (years)** |  |  |  |
| 0–19 | 4,291 (10) | 33 (3.0) | 33 (2.5) |
| 20–34 | 3,732 (8.8) | 187 (17) | 204 (16) |
| 35–49 | 5,845 (14) | 246 (23) | 288 (22) |
| 50–64 | 12,631 (30) | 388 (36) | 479 (37) |
| 65–79 | 11,984 (28) | 219 (20) | 287 (22) |
| 80+ | 3,794 (9.0) | 14 (1.3) | 20 (1.5) |
| **Sex** |  |  |  |
| Female | 18,340 (43) | 446 (41) | 543 (41) |
| Male | 23,937 (57) | 640 (59) | 767 (59) |
| Unknown | 0 (0) | <5 (<0.5) | <5 (<0.4) |
| **Race** |  |  |  |
| Asian | 2,431 (5.8) | 21 (1.9) | 30 (2.3) |
| Black | 2,915 (6.9) | 23 (2.1) | 28 (2.1) |
| Other | 242 (0.6) | 102 (9.4) | 153 (12) |
| Unknown | 325 (0.8) | 73 (6.7) | 95 (7.2) |
| White | 36,364 (86) | 868 (80) | 1,005 (77) |
| **Year of initial diagnosis** |  |  |  |
| 2011 | 5,066 (12) | 32 (2.9) | 32 (2.4) |
| 2012 | 5,267 (12) | 54 (5.0) | 54 (4.1) |
| 2013 | 5,323 (13) | 97 (8.9) | 97 (7.4) |
| 2014 | 5,228 (12) | 142 (13) | 142 (11) |
| 2015 | 5,425 (13) | 196 (18) | 196 (15) |
| 2016 | 5,267 (12) | 177 (16) | 177 (14) |
| 2017 | 5,367 (13) | 198 (18) | 198 (15) |
| 2018 | 5,334 (13) | 191 (18) | 191 (15) |
| 2019 | NA | NA | 135 (10) |
| 2020 | NA | NA | 84 (6.4) |
| 2021 | NA | NA | 5 (0.4) |
| Abbreviations: AJCC, American Joint Committee on Cancer; CGDB, Flatiron Health-Foundation Medicine Clinico-Genomic Database; FHRD, Flatiron Health Research Database; NA, not applicable; SEER, The Surveillance, Epidemiology, and End Results research database.<br>Note: Reported percentages may not equal exactly to 100% due to rounding. |  |  |  |

**Table 27.** Malignant Pleural Mesothelioma – Comparison of Demographic and Clinical Characteristics Between SEER vs CGDB.

| Characteristic | All eligible, n (%) |  |  | Stage IV at diagnosis, n (%) |  |  |
| --- | --- | --- | --- | --- | --- | --- |
|  | SEER | CGDB |  | SEER | CGDB |  |
| Initial diagnosis | 2011–18 | 2011–18 | 2011–20 | 2011–18 | 2011–18 | 2011–20 |
|  | N=5,439 | N=149 | N=184 | N=2,047 | N=38 | N=50 |
| **Age at initial diagnosis (years)** |  |  |  |  |  |  |
| 0–19 | <5 (<0.1) | <5 (<3.4) | <5 (<2.7) | <5 (<0.3) | <5 (<13) | <5 (<10) |
| 20–34 | 15 (0.3) | <5 (<3.4) | <5 (<2.7) | 8 (0.4) | <5 (<13) | <5 (<10) |
| 35–49 | 132 (2.4) | <5 (<3.4) | <5 (<2.7) | 66 (3.2) | <5 (<13) | <5 (<10) |
| 50–64 | 850 (16) | 31 (21) | 35 (19) | 385 (19) | 7 (18) | 7 (14) |
| 65–79 | 2,652 (49) | 94 (63) | 112 (61) | 1,009 (49) | 26 (68) | 33 (66) |
| 80+ | 1,789 (33) | 21 (14) | 32 (17) | 578 (28) | <5 (<13) | 8 (16) |
| **Sex** |  |  |  |  |  |  |
| Female | 1,259 (23) | 44 (30) | 56 (30) | 475 (23) | 12 (32) | 18 (36) |
| Male | 4,180 (77) | 105 (70) | 128 (70) | 1,572 (77) | 26 (68) | 32 (64) |
| **Race** |  |  |  |  |  |  |
| Asian | 212 (3.9) | <5 (<3.4) | <5 (<2.7) | 91 (4.4) | <5 (<13) | <5 (<10) |
| Black | 272 (5.0) | 7 (4.7) | 9 (4.9) | 111 (5.4) | <5 (<13) | <5 (<10) |
| Other | 27 (0.5) | 20 (13) | 28 (15) | 11 (0.5) | <5 (<13) | 6 (12) |
| Unknown | 17 (0.3) | 16 (11) | 18 (9.8) | 6 (0.3) | 7 (18) | 7 (14) |
| White | 4,911 (90) | 104 (70) | 127 (69) | 1,828 (89) | 26 (68) | 35 (70) |
| **AJCC stage at diagnosis** |  |  |  |  |  |  |
| Stage I | 1,009 (19) | 9 (6.0) | 11 (6.0) | NA | NA | NA |
| Stage II | 475 (8.7) | 11 (7.4) | 11 (6.0) | NA | NA | NA |
| Stage III | 977 (18) | 41 (28) | 49 (27) | NA | NA | NA |
| Stage IV | 2,047 (38) | 38 (26) | 50 (27) | NA | NA | NA |
| Unknown | 931 (17) | 50 (34) | 63 (34) | NA | NA | NA |
| **Year of initial diagnosis** |  |  |  |  |  |  |
| 2011 | 710 (13) | 7 (4.7) | 7 (3.8) | 254 (12) | <5 (<13) | <5 (<10) |
| 2012 | 685 (13) | 6 (4.0) | 6 (3.3) | 246 (12) | 0 (0) | 0 (0) |
| 2013 | 702 (13) | 9 (6.0) | 9 (4.9) | 248 (12) | <5 (<13) | <5 (<10) |
| 2014 | 686 (13) | 21 (14) | 21 (11) | 253 (12) | <5 (<13) | <5 (<10) |
| 2015 | 719 (13) | 17 (11) | 17 (9.2) | 283 (14) | 5 (13) | 5 (10) |
| 2016 | 670 (12) | 32 (21) | 32 (17) | 281 (14) | 10 (26) | 10 (20) |
| 2017 | 675 (12) | 25 (17) | 25 (14) | 269 (13) | 6 (16) | 6 (12) |
| 2018 | 592 (11) | 32 (21) | 32 (17) | 213 (10) | 8 (21) | 8 (16) |
| 2019 | NA | NA | 20 (11) | NA | NA | 8 (16) |
| 2020 | NA | NA | 14 (7.6) | NA | NA | <5 (<10) |
| 2021 | NA | NA | <5 (<2.7) | NA | NA | <5 (<10) |

Abbreviations: AJCC, American Joint Committee on Cancer; CGDB, Flatiron Health-Foundation Medicine Clinico-Genomic Database; FHRD, Flatiron Health Research Database; NA, not applicable; SEER, The Surveillance, Epidemiology, and End Results research database.
Note: Reported percentages may not equal exactly to 100% due to rounding.

**Table 28.**
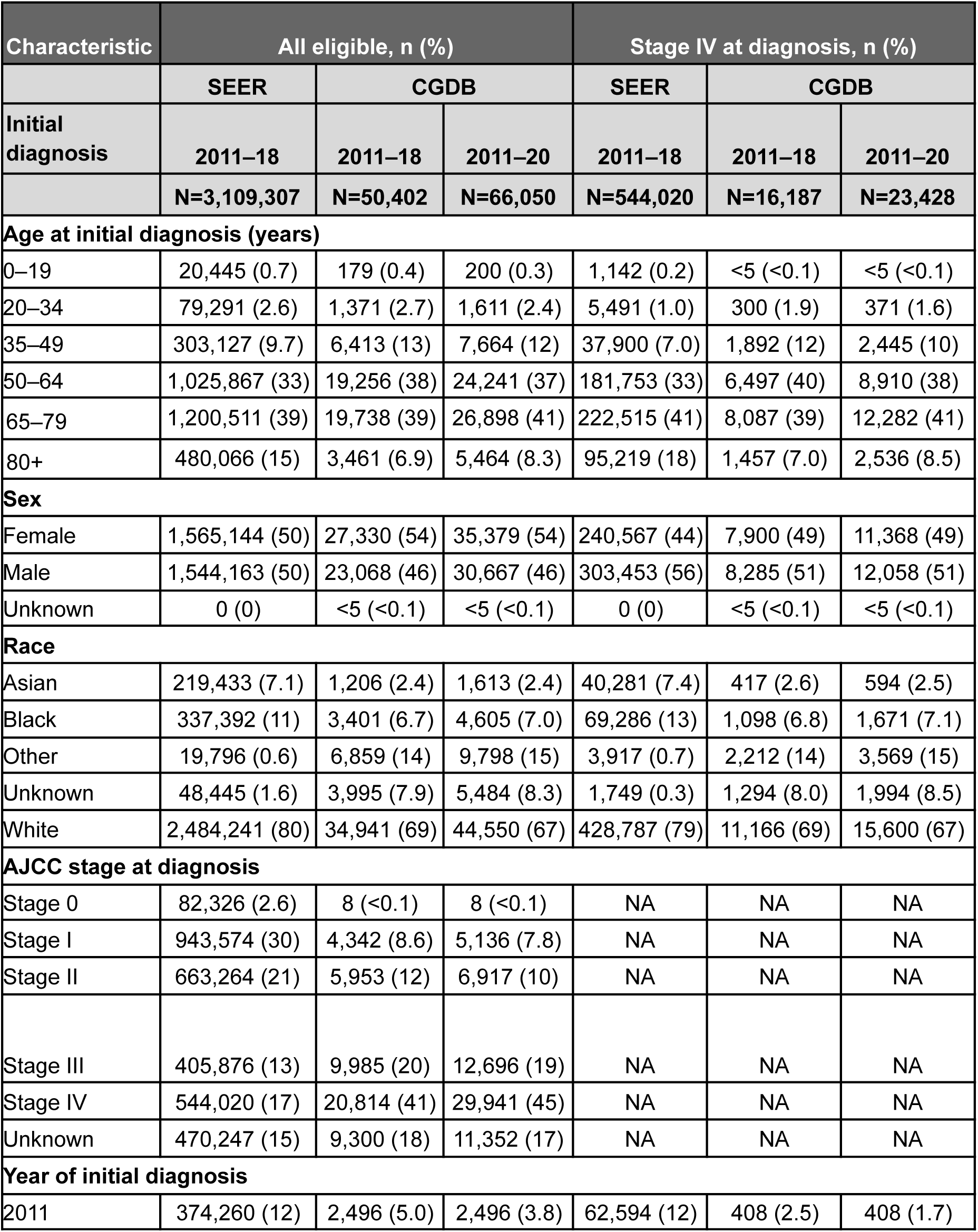
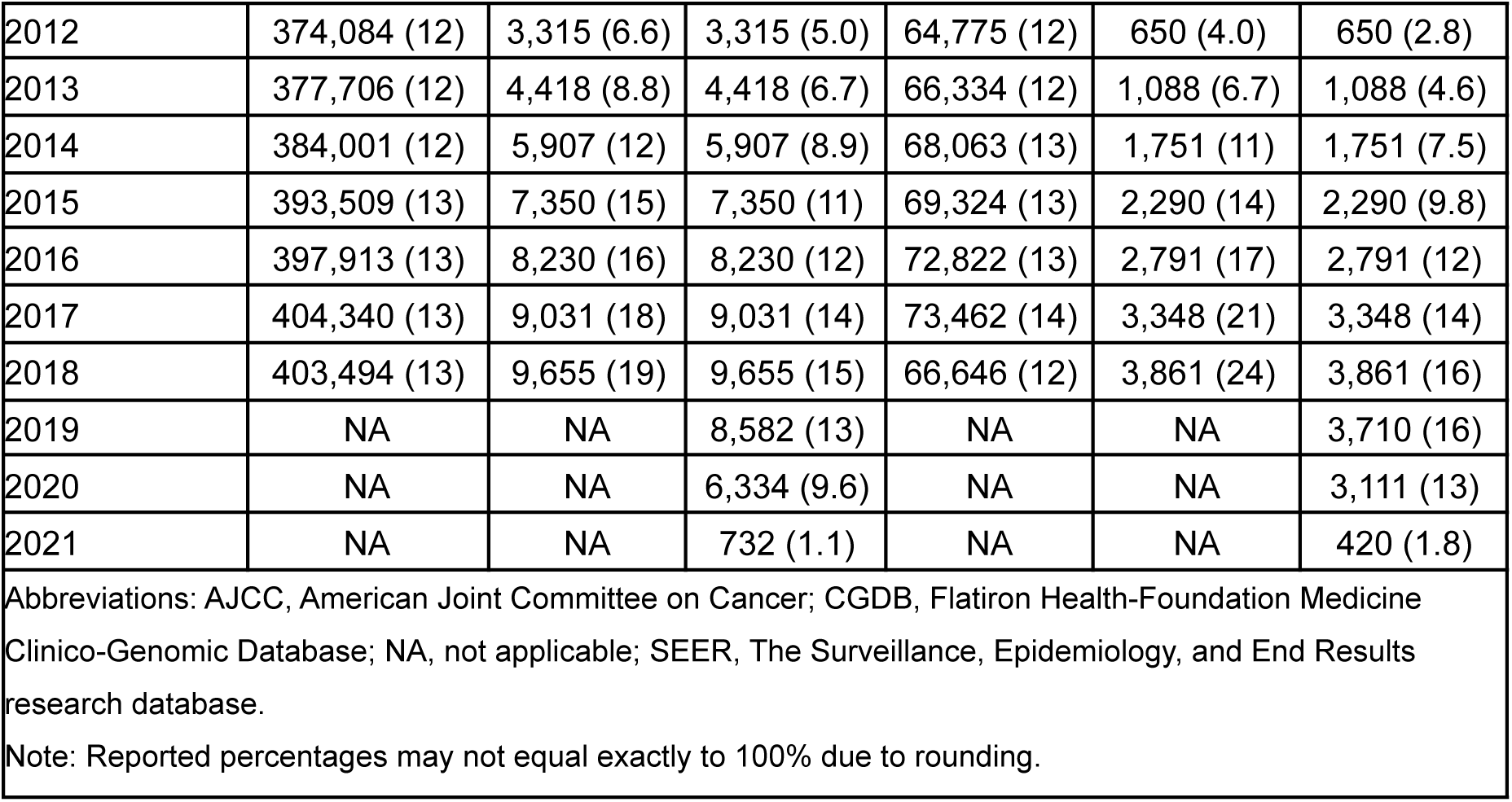
Solid Tumors – Comparison of Demographic and Clinical Characteristics Between SEER vs CGDB.

| Characteristic | All eligible, n (%) |  |  | Stage IV at diagnosis, n (%) |  |  |
| --- | --- | --- | --- | --- | --- | --- |
|  | SEER | CGDB |  | SEER | CGDB |  |
| Initial diagnosis | 2011–18 | 2011–18 | 2011–20 | 2011–18 | 2011–18 | 2011–20 |
|  | N=3,109,307 | N=50,402 | N=66,050 | N=544,020 | N=16,187 | N=23,428 |
| **Age at initial diagnosis (years)** |  |  |  |  |  |  |
| 0–19 | 20,445 (0.7) | 179 (0.4) | 200 (0.3) | 1,142 (0.2) | <5 (<0.1) | <5 (<0.1) |
| 20–34 | 79,291 (2.6) | 1,371 (2.7) | 1,611 (2.4) | 5,491 (1.0) | 300 (1.9) | 371 (1.6) |
| 35–49 | 303,127 (9.7) | 6,413 (13) | 7,664 (12) | 37,900 (7.0) | 1,892 (12) | 2,445 (10) |
| 50–64 | 1,025,867 (33) | 19,256 (38) | 24,241 (37) | 181,753 (33) | 6,497 (40) | 8,910 (38) |
| 65–79 | 1,200,511 (39) | 19,738 (39) | 26,898 (41) | 222,515 (41) | 8,087 (39) | 12,282 (41) |
| 80+ | 480,066 (15) | 3,461 (6.9) | 5,464 (8.3) | 95,219 (18) | 1,457 (7.0) | 2,536 (8.5) |
| **Sex** |  |  |  |  |  |  |
| Female | 1,565,144 (50) | 27,330 (54) | 35,379 (54) | 240,567 (44) | 7,900 (49) | 11,368 (49) |
| Male | 1,544,163 (50) | 23,068 (46) | 30,667 (46) | 303,453 (56) | 8,285 (51) | 12,058 (51) |
| Unknown | 0 (0) | <5 (<0.1) | <5 (<0.1) | 0 (0) | <5 (<0.1) | <5 (<0.1) |
| **Race** |  |  |  |  |  |  |
| Asian | 219,433 (7.1) | 1,206 (2.4) | 1,613 (2.4) | 40,281 (7.4) | 417 (2.6) | 594 (2.5) |
| Black | 337,392 (11) | 3,401 (6.7) | 4,605 (7.0) | 69,286 (13) | 1,098 (6.8) | 1,671 (7.1) |
| Other | 19,796 (0.6) | 6,859 (14) | 9,798 (15) | 3,917 (0.7) | 2,212 (14) | 3,569 (15) |
| Unknown | 48,445 (1.6) | 3,995 (7.9) | 5,484 (8.3) | 1,749 (0.3) | 1,294 (8.0) | 1,994 (8.5) |
| White | 2,484,241 (80) | 34,941 (69) | 44,550 (67) | 428,787 (79) | 11,166 (69) | 15,600 (67) |
| **AJCC stage at diagnosis** |  |  |  |  |  |  |
| Stage 0 | 82,326 (2.6) | 8 (<0.1) | 8 (<0.1) | NA | NA | NA |
| Stage I | 943,574 (30) | 4,342 (8.6) | 5,136 (7.8) | NA | NA | NA |
| Stage II | 663,264 (21) | 5,953 (12) | 6,917 (10) | NA | NA | NA |
| Stage III | 405,876 (13) | 9,985 (20) | 12,696 (19) | NA | NA | NA |
| Stage IV | 544,020 (17) | 20,814 (41) | 29,941 (45) | NA | NA | NA |
| Unknown | 470,247 (15) | 9,300 (18) | 11,352 (17) | NA | NA | NA |
| **Year of initial diagnosis** |  |  |  |  |  |  |
| 2011 | 374,260 (12) | 2,496 (5.0) | 2,496 (3.8) | 62,594 (12) | 408 (2.5) | 408 (1.7) |
| 2012 | 374,084 (12) | 3,315 (6.6) | 3,315 (5.0) | 64,775 (12) | 650 (4.0) | 650 (2.8) |
| 2013 | 377,706 (12) | 4,418 (8.8) | 4,418 (6.7) | 66,334 (12) | 1,088 (6.7) | 1,088 (4.6) |
| 2014 | 384,001 (12) | 5,907 (12) | 5,907 (8.9) | 68,063 (13) | 1,751 (11) | 1,751 (7.5) |
| 2015 | 393,509 (13) | 7,350 (15) | 7,350 (11) | 69,324 (13) | 2,290 (14) | 2,290 (9.8) |
| 2016 | 397,913 (13) | 8,230 (16) | 8,230 (12) | 72,822 (13) | 2,791 (17) | 2,791 (12) |
| 2017 | 404,340 (13) | 9,031 (18) | 9,031 (14) | 73,462 (14) | 3,348 (21) | 3,348 (14) |
| 2018 | 403,494 (13) | 9,655 (19) | 9,655 (15) | 66,646 (12) | 3,861 (24) | 3,861 (16) |
| 2019 | NA | NA | 8,582 (13) | NA | NA | 3,710 (16) |
| 2020 | NA | NA | 6,334 (9.6) | NA | NA | 3,111 (13) |
| 2021 | NA | NA | 732 (1.1) | NA | NA | 420 (1.8) |

Abbreviations: AJCC, American Joint Committee on Cancer; CGDB, Flatiron Health-Foundation Medicine Clinico-Genomic Database; NA, not applicable; SEER, The Surveillance, Epidemiology, and End Results research database.
Note: Reported percentages may not equal exactly to 100% due to rounding.

**Table 29.** Hematologic Malignancies – Comparison of Demographic and Clinical Characteristics Between SEER vs CGDB.

| Characteristic | All eligible, n (%) |  |  |
| --- | --- | --- | --- |
|  | SEER | CGDB |  |
| Initial diagnosis | 2011–18 | 2011–18 | 2011–20 |
|  | N=395,565 | N=2,271 | N=2,972 |
| **Age at initial diagnosis (years)** |  |  |  |
| 0–19 | 15,215 (3.8) | 24 (1.1) | 26 (0.9) |
| 20–34 | 17,527 (4.4) | 105 (4.6) | 128 (4.3) |
| 35–49 | 34,567 (8.7) | 217 (9.6) | 264 (8.9) |
| 50–64 | 99,853 (25) | 695 (31) | 857 (29) |
| 65–79 | 146,750 (37) | 998 (44) | 1,323 (44) |
| 80+ | 81,653 (21) | 234 (10) | 376 (13) |
| **Sex** |  |  |  |
| Female | 174,271 (44) | 905 (40) | 1,208 (41) |
| Male | 221,294 (56) | 1,366 (60) | 1,764 (59) |
| Unknown |  |  |  |
| **Race** |  |  |  |
| Asian | 25,650 (6.5) | 42 (1.8) | 57 (1.9) |
| Black | 40,181 (10) | 123 (5.4) | 169 (5.7) |
| Other | 2,274 (0.6) | 364 (16) | 523 (18) |
| Unknown | 6,190 (1.6) | 180 (7.9) | 231 (7.8) |
| White | 321,270 (81) | 1,562 (69) | 1,992 (67) |
| **Year of initial diagnosis** |  |  |  |
| 2011 | 47,040 (12) | 128 (5.6) | 128 (4.3) |
| 2012 | 48,180 (12) | 160 (7.0) | 160 (5.4) |
| 2013 | 48,779 (12) | 222 (9.8) | 222 (7.5) |
| 2014 | 50,124 (13) | 272 (12) | 272 (9.2) |
| 2015 | 50,608 (13) | 319 (14) | 319 (11) |
| 2016 | 50,916 (13) | 367 (16) | 367 (12) |
| 2017 | 50,585 (13) | 385 (17) | 385 (13) |
| 2018 | 49,333 (12) | 418 (18) | 418 (14) |
| 2019 | NA | NA | 374 (13) |
| 2020 | NA | NA | 297 (10.0) |
| 2021 | NA | NA | 30 (1.0) |
| Abbreviations: CGDB, Flatiron Health-Foundation Medicine Clinico-Genomic Database; NA, not applicable; SEER, The Surveillance, Epidemiology, and End Results research database.<br>Note: Reported percentages may not equal exactly to 100% due to rounding. |  |  |  |

**Table 30.** All Patients – Comparison of Demographic and Clinical Characteristics Between SEER vs CGDB.

| Characteristic | All eligible, n (%) |  |  | Stage IV at diagnosis, n (%) |  |  |
| --- | --- | --- | --- | --- | --- | --- |
|  | SEER | CGDB |  | SEER | CGDB |  |
| Initial diagnosis | 2011–18 | 2011–18 | 2011–20 | 2011–18 | 2011–18 | 2011–20 |
|  | N=3,504,872 | N=60,782 | N=78,084 | N=600,500 | N=22,450 | N=32,064 |
| **Age at initial diagnosis (years)** |  |  |  |  |  |  |
| 0–19 | 35,660 (1.0) | 204 (0.3) | 227 (0.3) | 2,331 (0.4) | 48 (0.2) | 57 (0.2) |
| 20–34 | 96,818 (2.8) | 1,934 (3.2) | 2,237 (2.9) | 8,080 (1.3) | 554 (2.5) | 685 (2.1) |
| 35–49 | 337,694 (9.6) | 8,979 (15) | 10,506 (13) | 43,196 (7.2) | 2,872 (13) | 3,668 (11) |
| 50–64 | 1,125,720 (32) | 23,361 (38) | 28,861 (37) | 197,587 (33) | 8,907 (40) | 12,121 (38) |
| 65–79 | 1,514,540 (38) | 23,564 (39) | 31,324 (40) | 253,647 (40) | 8,892 (40) | 13,248 (41) |
| 80+ | 613,271 (15) | 2,740 (4.5) | 4,929 (6.3) | 109,324 (17) | 1,177 (5.2) | 2,285 (7.1) |
| **Sex** |  |  |  |  |  |  |
| Female | 1,739,415 (50) | 36,215 (60) | 45,493 (58) | 264,330 (44) | 11,704 (52) | 16,627 (52) |
| Male | 1,765,457 (50) | 24,562 (40) | 32,586 (42) | 336,170 (56) | 10,743 (48) | 15,434 (48) |
| Unknown | 0 (0) | 5 (<0.1) | 5 (<0.1) | 0 (0) | <5 (<0.1) | <5 (<0.1) |
| **Race** |  |  |  |  |  |  |
| Asian | 245,083 (7.0) | 1,418 (2.3) | 1,858 (2.4) | 43,936 (7.3) | 540 (2.4) | 770 (2.4) |
| Black | 377,573 (11) | 4,398 (7.2) | 5,784 (7.4) | 74,352 (12) | 1,629 (7.3) | 2,450 (7.6) |
| Other | 22,070 (0.6) | 8,410 (14) | 11,687 (15) | 4,209 (0.7) | 3,040 (14) | 4,864 (15) |
| Unknown | 54,635 (1.6) | 4,782 (7.9) | 6,436 (8.2) | 2,367 (0.4) | 1,805 (8.0) | 2,732 (8.5) |
| White | 2,805,511 (80) | 41,774 (69) | 52,319 (67) | 475,636 (79) | 15,436 (69) | 21,248 (66) |
| **AJCC stage at diagnosis** |  |  |  |  |  |  |
| Stage 0 | 82,326 (2.3) | 45 (<0.1) | 50 (<0.1) | NA | NA | NA |
| Stage I | 984,375 (28) | 5,609 (9.2) | 6,511 (8.3) | NA | NA | NA |
| Stage II | 690,712 (20) | 9,000 (15) | 10,143 (13) | NA | NA | NA |
| Stage III | 433,395 (12) | 12,507 (21) | 15,490 (20) | NA | NA | NA |
| Stage IV | 600,500 (17) | 22,450 (37) | 32,064 (41) | NA | NA | NA |
| Unknown | 713,564 (20) | 11,171 (18) | 13,826 (17) | NA | NA | NA |
| **Year of initial diagnosis** |  |  |  |  |  |  |
| 2011 | 421,300 (12) | 3,481 (5.7) | 3,481 (4.5) | 68,891 (11) | 611 (2.7) | 611 (1.9) |
| 2012 | 422,264 (12) | 4,432 (7.3) | 4,432 (5.7) | 71,092 (12) | 972 (4.3) | 972 (3.0) |
| 2013 | 426,485 (12) | 5,660 (9.3) | 5,660 (7.2) | 72,864 (12) | 1,586 (7.1) | 1,586 (4.9) |
| 2014 | 434,125 (12) | 7,328 (12) | 7,328 (9.4) | 74,835 (12) | 2,495 (11) | 2,495 (7.8) |
| 2015 | 444,117 (13) | 8,888 (15) | 8,888 (11) | 76,176 (13) | 3,171 (14) | 3,171 (9.9) |
| 2016 | 448,829 (13) | 9,750 (16) | 9,750 (12) | 79,481 (13) | 3,823 (17) | 3,823 (12) |
| 2017 | 454,925 (13) | 10,363 (17) | 10,363 (13) | 79,986 (13) | 4,573 (20) | 4,573 (14) |
| 2018 | 452,827 (13) | 10,880 (18) | 10,880 (14) | 77,175 (13) | 5,219 (23) | 5,219 (16) |
| 2019 | NA | NA | 9,514 (12) | NA | NA | 4,952 (15) |
| 2020 | NA | NA | 7,001 (9.0) | NA | NA | 4,128 (13) |
| 2021 | NA | NA | 787 (1.0) | NA | NA | 534 (1.7) |

Abbreviations: AJCC, American Joint Committee on Cancer; CGDB, Flatiron Health-Foundation Medicine Clinico-Genomic Database; FHRD, Flatiron Health Research Database; NA, not applicable; SEER, The Surveillance, Epidemiology, and End Results research database.
Note: Reported percentages may not equal exactly to 100% due to rounding.

## Discussion

The objective of this study was to describe how the distribution of demographic and clinical characteristics may differ when comparing the CGDBs with other US oncology-specific RWD sources, specifically the FHRDs and SEER. Characteristics of patients in the CGDBs and FHRDs were largely similar across the disease-specific databases, which is expected since they are built from the same source EHR data and have overlapping patients. Differences in the distribution of patient characteristics between SEER and the CGDBs and FHRDs likely stem from the distinct data collection methods and resulting populations. We observed that these differences were less apparent in diseases for which targeted therapies are approved (e.g., NSCLC and Melanoma) and also in other diseases when analyses were limited to patients diagnosed with stage IV disease, which may be related to SEER including patients of all stages, while certain CGDB and FHRD datasets include only early stage patients who subsequently develop advanced or metastatic disease.

Overall, the tumor type distribution between the CGDBs and SEER was similar. A notable difference was in disease-specific CGDBs that had a larger relative proportion of cases for diseases where CGP testing is more widely used, such as thoracic malignancies and soft tissue sarcomas. A lower relative proportion of cases was observed in hematologic malignancies, which may be a result of the comparatively low use of CGP in these diseases, in which fewer molecularly targeted therapies have been approved by the US Food and Drug Administration (FDA).

With regards to age, the CGDB cohorts have a lower proportion of patients who were aged 80 years or older at diagnosis compared to FHRD and SEER cohorts. Moreover, a slight proportional increase in CGP testing in patients aged 80 and older is observed when 2011–20 is compared to 2011–2018 across the majority of diseases representing greater adoption of CGP testing across this age group. A difference in the proportion of patients age 80 years and older between FHRD and CGDB versus SEER in some diseases might be due to:

- Patient was advised and/or opted for no treatment so their care is managed by a non-hematology/oncology provider and/or hospice
- Patient may have a rapid clinical decline and do not live long enough to establish care with an outpatient hematology/oncology provider
- Patient may not have had an indication to receive systemic therapy (e.g., Stage I and majority of Stage II CRC) and the disease can be treated by a non-hematology/oncology provider, or
- Patient may have indolent, asymptomatic disease managed with observation alone by a non-hematology/oncology provider (e.g., CLL, FL).

Of note, the proportion of patients age 80 and older with metastatic prostate cancer was higher for the FHRD cohort than CGDB and SEER cohorts. This difference with the CGDB cohort may reflect the more indolent disease biology typically seen in older patients with metastatic prostate cancer for which NGS testing is not considered by providers taking a conservative management approach. Other factors such as performance status, comorbidities, and patient choice in this age group may also be potential limitations for providers who may otherwise utilize NGS testing to guide clinical trial or off-label drug use. The difference with SEER is likely due to the FHRD cohort representing a metastatic-only cohort (i.e., older pts in SEER with non-metastatic disease may be managed by non-oncologists).

The overall gender distributions were similar across databases. With regards to race, the proportion of incomplete records for race in the CGDBs and FHRDs was greater than in SEER, which is expected given the different data collection methods between databases. However, this limits our ability to draw meaningful comparisons across the different data sources. Despite its inclusion as part of the Meaningful Use criteria for EHRs in the US, collection of race data continues to be incomplete in many clinical settings. While recently established programs such as the Oncology Care Model (OCM) and Merit-based Incentive Payment System (MIPS) require specific documentation, thus aiding in increased completeness across several demographic fields including race, a limited number of practices and eligible patients are enrolled. As a result, we continue to see incomplete structured capture of race information in EHR data. In contrast, SEER mandates data quality standards that require registries to reach high levels of completeness for these demographic variables. However, despite a higher proportion of incomplete race data relative to SEER, race distributions between the FHRDs and CGDBs were relatively similar across disease-specific and disease-agnostic comparisons.

The approaches for capturing tumor stage are different in SEER compared to the CGDBs and FHRDs, which impacts the completeness of stage data. We observed lower completeness of information on stage at initial diagnosis in the CGDBs and FHRDs since stage data are EHR-derived, where information on stage at initial diagnosis is not always documented by the treating oncologist either explicitly or with a consistent source of truth to allow for calculation of stage with high reproducibility. Lower capture of stage at diagnosis may also be due to lower documentation of stage in diseases where non-oncologists are more involved in the initial diagnosis (e.g., early stage bladder/prostate tumors), as well as in CGDBs and FHRDs developed with stricter requirements for calculation and coding to address greater complexity in staging rules (i.e., disease-specific rules apply for which some databases include derived stage group from documentation of TNM Classification of Malignant Tumors or metastasis when explicitly documented by a clinician while for other databases, no derivation of stage group is performed). In comparison, SEER allows the abstractor to determine the stage group in situations where involvement is described with non-definitive (ambiguous) terminology and relies on multiple sites of care as sources and disease stage is intentionally entered into their databases via mandated calculation and coding by trained tumor registrars.^(20,23)^

Overall the stage distributions for solid tumors were similar across CGDBs and FHRDs. In comparisons that included CGDB and FHRD patients with advanced/metastatic disease only (endometrial, bladder, and prostate cancer), the proportion of patients initially diagnosed with earlier stage disease is lower than SEER, which is expected as early stage patients who are cured of their disease do not qualify for inclusion in these databases. Proportions of patients reported in the SEER metastatic breast cancer cohort are not directly comparable to the FHRD and CGDB cohorts, given that the SEER cohort represents all patients with distant breast cancer at initial diagnosis whereas the breast cancer FHRD and CGDB represent patients diagnosed with Stage IV disease at initial diagnosis and patients diagnosed with earlier stage disease who subsequently developed metastasis. We also observed that some hematologic disease-specific CGDBs (i.e., CLL, follicular lymphoma) had more patients with advanced stages when compared to the respective FHRDs and SEER cohorts, and generally had smaller cohorts than expected given the size of respective FHRDs. This could be because CGP is not typically recommended for hematologic malignancies in clinical practice guidelines^(24–27)^ thus clinicians may not order CGP until they have exhausted conventional treatment options. We also observed overall similar distributions between the disease-specific FHRDs and CGDBs for patients diagnosed with solid tumors where stage IV AJCC classification is not exclusive to distant metastatic disease at diagnosis, including gastric/esophageal/GEJ cancer, hepatocellular carcinoma, prostate cancer, head and neck cancer, RCC, and urothelial/bladder cancer. This demonstrates the adoption of CGP testing in patients with regional extension of disease, in addition to those patients whose disease has spread to distant organs and for whom—while the extent of disease differs—prognosis is similar.

Finally, we found that overall there were similar distributions of diagnosis year across databases. There were some diseases (e.g., cholangiocarcinoma, pancreatic adenocarcinoma) where a slightly higher proportion of patients was diagnosed more recently in the CGDBs than in earlier years than would be expected, which might be driven by adoption of CGP testing associated with more recently approved biomarker-driven therapies in these diseases.

This study is subject to limitations. First, the CGDBs and FHRDs have data that are more recent than SEER. As of March 2021, the most recent data available from SEER are through December 31, 2018, so any demographic changes and evolution of staging for the cancer type over time could cause differences in these distributions. We tried to address this limitation by performing comparisons on the subsets of the CGDBs and FHRDs for patients with a diagnosis on or prior to the data cutoff date for the most recent SEER data. Second, the scope of characteristics compared was dependent upon the data elements that were available across all databases. For example, lack of geographic region data in the CGDBs makes it infeasible to compare the geographic distribution of cases against FHRDs and SEER, which is an important characteristic when evaluating representativeness of a database because treatment patterns may differ by geographic region. Third, there were smaller cohort sizes in the CGDBs compared to both the FHRDs and SEER for hematologic malignancies, which is expected since CGP testing is less commonly incorporated into standard of care in those diseases. Fourth, unlike the disease-specific CGDBs, the disease-agnostic CGDB does not apply disease-specific histology filters based on the reported histology of the FMI specimens, such that the CGP test specimen tumor type may not be consistent with the disease-agnostic CGDB tumor type. Fifth, given that the FHRDs and CGDBs include active follow-up to capture the development of advanced/metastatic disease following initial diagnosis, while SEER does not, proportions of patients reported in the SEER cohort are not directly comparable to some of the FHRD and CGDB cohorts.

In conclusion, this comparative analysis of real-world, US-based clinical oncology databases provides crucial insights into the representativeness of the data across the three databases. Overall, the CGDB has a similar distribution of core patient characteristics as both the FHRDs and SEER. As clinico-genomic data are expected to change over time due to the increased adoption of CGP as part of routine clinical practice for an increasing number of tumor types, ongoing monitoring and evaluation of the representativeness of these data will be critical to help researchers and regulators to contextualize evidence from the CGDBs. Additionally, given the growing comprehension of the ways in which a cancer’s molecular profile can dictate therapeutic response and the incomplete understanding of biomarkers that emerges through traditional drug development,^(28)^ retrospective analyses in clinico-genomic data will play an increasingly important role in informing clinical decision making.

## Data Availability

The data that support the findings of this study have been originated by Flatiron Health, Inc. Requests for data sharing by license or by permission for the specific purpose of replicating results in this manuscript can be submitted to

## Acknowledgments

The authors would like to thank Cody Patton and Hannah Gilham (Flatiron Health, Inc) for medical writing and publication management support.

