## Supplementary material for "Comparison of Population Characteristics in Real-World Clinical Oncology Databases in the US: Flatiron Health-Foundation Medicine Clinico-Genomic Databases, Flatiron Health Research Databases, and the National Cancer Institute SEER Population-Based Cancer Registry": Snow_Comparison_CG_SEER_Whitepaper_FINAL_Suppl_Nov2022.pdf

### Supplementary Materials

| **Supplementary Table 1: Comparison of Key Cohort Selection Criteria for Disease-specific Builds** | | |
|---|---|---|
| | **CGDBs** | **FHRDs** |
| **Diagnosis Date Window** | No diagnosis date window, requires 2+ clinical visits on/after 1/1/2011 | In addition to requiring 2+ clinical visits on/after 1/1/2011, most FHRDs require a diagnosis on or after 1/1/2011.<br><br>The following FHRDs have different diagnosis date windows which do not apply in the corresponding CGDB:<br>● AML (January 1, 2014)<br>● Colorectal Cancer (January 1, 2013)<br>● Advanced Endometrial (January 1, 2013)<br>● Pancreatic Adenocarcinoma (January 1, 2014)<br>● Metastatic Prostate Cancer (January 1, 2013)<br>● SCLC (January 1, 2013) |
| **Stage** | CGDBs include patients of all stages with the exception of the following databases limited to advanced/metastatic cohorts:<br>● Advanced Endometrial Cancer<br>● Metastatic Prostate Cancer<br>● Advanced Bladder Cancer | Solid tumor FHRDs are advanced/metastatic cohorts with the exception of the following databases for which patients of all stages are included:<br>● HCC<br>● Ovarian Cancer<br>● SCLC |

| **Supplementary Table 2. Overview of Variable Definitions and Steps Taken to Align Variables Across the Different Databases** | | |
|---|---|---|
| **Variable** | **FHRDs and CGDBs** | **SEER** |
| **Age at initial diagnosis** | Age at diagnosis calculated by taking the difference between a patient's date of birth (rounded to birth year for de-identification reasons) and their year of diagnosis. Birth years beyond a predetermined year are capped; however, actual birth years were used for this internal research study. | Ordered categorical variable defined using SEER's age at diagnosis variable |
| **Sex/Gender** | Binary categorical variable with the following categories:<br>● Male<br>● Female<br>● Unknown<br><br>Notes: FHRD and CGDB data largely reflect biological gender information. The FHRD and CGDB gender variable was compared against SEER's sex variable. | |
| **Race** | As captured in the EHR<br>● White = "White"<br>● Black or African American = "Black or African American"<br>● Asian = "Asian"<br>● Other Race = "Other"<br>● Unknown = "Unknown" | ● White = "White"<br>● Black or African American = "Black or African American"<br>● Asian = "Asian" or "Pacific Islander"<br>● Other Race = "American Indian" or "Alaska Native"<br>● Unknown = "Unknown" |
| **Stage at diagnosis** | Cancer staging information was collected as entered into the EHR by the treating physician or otherwise as assessed by Flatiron Health abstractors; during the study time period, the applicable staging criteria for solid tumors were those of the AJCC 7th edition and 8th edition manuals, including the adoption of the FIGO staging system by the AJCC for endometrial cancer.<br>Rai staging for CLL, Ann Arbor Staging System for DLBCL, FL, and mantle cell lymphoma, the ISS for multiple myeloma, and the WHO Classification for AML were used. | For all relevant diseases except Early Breast Cancer:<br>● For diagnoses before 2016: Derived AJCC Stage Group v7<br>● For diagnoses on or after 2016: Derived SEER combined stage group<br><br>Early Breast Cancer:<br><br>Considered to be diagnosed with early stage disease if one of the following variables contained 1, 2, or 3 from: derived_ajcc_stage_group_6th_ed_2004_2015, derived_seer_cmb_stg_grp_2016_2017, derived_eod_2018_stage_group_201 |

| | | |
|---|---|---|
| | | 8) |
| **Metastatic at diagnosis** | This breakdown was only done for FHRD and CGDB solid tumor diseases where Stage IV AJCC classification is not exclusive to distant metastatic disease at diagnosis, including:<br><br>**Gastric/Esophageal/GEJ**<br>● Met: Stage IV, M1<br>● Non-Met: M0 or non-stage IV (including unknown AJCC group stage if M0 is known)<br><br>**HNC**<br>Because AJCC v8 significantly changed how HPV+ oropharynx site tumors are staged, we defined Met as followed:<br>● pre-July 2018: Stage IVC<br>● July 2018 and onward: Stage IVC or Stage IV if HPV/p16+ and site = Oropharynx or Unknown primary Head and Neck cancer<br>And Non-Met as followed:<br>● pre-July 2018: non-Stage IVC<br>● July 2018 and onward: non-Stage IVC or non-Stage IV if HPV/p16+ and site = Oropharynx or Unknown primary Head and Neck cancer<br><br>**HCC**<br>● Met: Stage IV (not otherwise specified) or Stage IVB, M1<br>● Non-Met: M0 or non-stage IV/IVB (including unknown AJCC group stage if M0 is known)<br><br>**RCC**<br>● Met: Stage IV (excluding T4NxM0), M1<br>● Non-Met: M0, or non-stage IV (including unknown AJCC group stage if M0 is known)<br><br>**Urothelial/Bladder**<br>● Met: Stage IV (excluding T4bNxM0), M1<br>● Non-Met: M0 or non-stage IV (including unknown AJCC | NA |

| | |
|---|---|
| | group stage if M0 is known)<br><br>**Prostate**<br>● Met: Stage IV, M1<br>● Non-Met: M0 or non-stage IV (including unknown AJCC group stage if M0 is known) |
| **Year of initial diagnosis** | Ordered categorical variable defined as abstracted year of initial diagnosis date in FHRDs and CGDBs and as diagnosis date in SEER |
| Abbreviations: AJCC, American Joint Committee on Cancer; AML, acute myeloid leukemia; CGDB, Flatiron Health-Foundation Medicine Clinico-Genomic Database; CLL, chronic lymphocytic leukemia; DLBCL, diffuse large B-cell lymphoma; EHR, electronic health record; FHRD, Flatiron Health Research Database; FIGO, International Federation of Gynecology and Obstetrics; FL, follicular lymphoma; GEJ, gastroesophageal junction; HCC< hepatocellular carcinoma; HNC, head and neck cancer; HPV, human papillomavirus; ISS, International Staging System; NA, not applicable; RCC, renal cell carcinoma; SEER, The Surveillance, Epidemiology, and End Results research database. | |
